# Drivers and prevalence of COVID-19 vaccine uptake among homeless and precariously housed people in France: a cross-sectional population-based study

**DOI:** 10.1101/2022.07.18.22276918

**Authors:** Thomas Roederer, Bastien Mollo, Charline Vincent, Ghislain Leduc, Jessica Sayyad, Marine Mosnier, Stéphanie Vandentorren

## Abstract

**Background:** Few global data exist regarding COVID-19 vaccine coverage in people experiencing homelessness (PEH) or precariously housed (PH) who are at high risk for COVID-19 infection, hospitalization, and death. Given the absence of documented French data, we aimed to determine COVID-19 vaccine coverage in PEH/PH in France, and its drivers.

**Methods:** We carried out a cross-sectional study following a two-stage cluster-sampling design in Ile-de-France and Marseille, France, in late 2021. Participants aged over 18 years were recruited where they slept the previous night, and then stratified for analysis into three housing groups (“Streets”, “Accommodated”, and “Housed”). Interviews were conducted face-to-face in the participant’s preferred language. Multilevel univariate and multivariable logistic regression models were built.

**Findings:** 3,690 individuals were surveyed: 855 in the “Housed” stratum, 2,321 in the “Accommodated” stratum and 514 in the “Streets” stratum. 76·2% (95%CI 74·3-78·1) reported receiving at least one COVID-19 vaccine dose. Vaccine uptake varied by stratum, with uptake highest (85.6%; reference) in “Housed”, followed by “Accommodated” (75·4%; AOR=0·79 ; 95%CI 0·51-1·09 vs Housed) and lowest in “Streets” (42·0%; AOR=0·38 ; 95%CI 0·25-0·57 vs Housed). Use for vaccine certificate, socioeconomic drivers and vaccine hesitancy explained vaccine coverage.

**Interpretation:** In France, PEH/PH are less likely than the general population likely to receive COVID-19 vaccines; with the most excluded being the least likely. The influence of both structural drivers and vaccine beliefs in PEH/PH reinforces the importance of targeted outreach, on-site vaccination and sensitisation activities to further vaccine uptake.

**Funding:** Santé Publique France, Agence Nationale de Recherches sur le Sida/Capnet, Agence Régionale de Santé – Ile de France, Médecins Sans Frontières, and Société de Pathologie Infectieuse de Langue Française.

## INTRODUCTION

Evidence from the early COVID-19 waves suggests that population subgroups, such as people experiencing homelessness (PEH) or precariously housed (PH), are disproportionately exposed to infection [1-3] and the severe forms of disease [4-6], as well as suffering from greater mental health and social impacts [7]. Transmission risk is worsened by factors specific to these groups, such as precarious living conditions, high population density, need to access food distribution services, poor access to sanitation and hygiene, and difficulties accessing care [1, 2, 3, 5, 7]. COVID-19 prevention measures such as social distancing and self-isolation are challenging to maintain for such groups [1, 3, 5].

In 2021, highly efficacious COVID-19 vaccines became available, providing strong protection against severe disease, hospitalization and death. It is already known that PEH/PH tend to uptake vaccination against diseases other than COVID-19 to a lower degree than the general population [8-11]. Obstacles to vaccination for PEH/PH include practical barriers and service limitations [8, 10, 11], suboptimal experiences with vaccines or health services [8, 9, 10], modern medicine/vaccine hesitancy [8, 9, 11]. Structural obstacles include poor housing [8, 11, 12], inadequate medical coverage and access to care [8, 9, 10, 11, 12, 13], and not considering disease prevention a priority [11, 13]. Migrants and refugees also encounter obstacles to vaccination within host countries, including language barriers and lack of access to information [8, 9, 11, 12, 13, 14], not considering health as a priority [14], high mobility/no fixed address [9, 10, 12-14], and lack of suitable healthcare providers [12-14]. Moreover, migrants and refugees may also be reluctant to take up vaccination, for fear of deportation while waiting for right to reside [13, 14].

France’s COVID-19 vaccination strategy was implemented in five stages from January 2021 onwards, with vaccination cost-free, regardless of medical coverage or administrative status (see appendix 1 for detailed timeline). Expansion to all adults aged over 18, including PEH/PH, was then implemented in June 2021. Other important developments were the introduction of the Pass Sanitaire (vaccine certificate) in July 2021 and the introduction of third (booster) doses for all adults, including PEH/PH, from November 2021 on. Implementation of vaccination in PEH/PH was reliant on non-governmental organizations [15].

Estimates of the PEH/PH population for France in 2021 suggest this group comprises around 300,000 people, with roughly 150,000 in the Ile-de-France region and Marseille. Of these, 25,000 are housed in workers’ hostels, 50,000 in centers for asylum seekers and emergency shelters, and 35,000 in social hostels [16]. 2,800 are estimated to be permanently living rough in Paris, and around 1,500 in Marseille [17]. No official French data for COVID-19 vaccine coverage in migrants, homeless, or roofless populations exist. Few global data are available on vaccine uptake among PEH/PH; we identified only three quantitative [18-20] studies and no reports on drivers of or barriers to COVID-19 vaccination.

Given higher exposure to COVID-19 and the lack of data, it is crucial to estimate vaccination coverage among PEH/PH and to better understand determinants of vaccination. We aimed to estimate vaccination coverage in PEH and those precariously housed, and to analyse factors associated with vaccination status.

## METHODS

### Survey design and sampling strategy

We performed a cross-sectional study between 15 November and 22 December 2021 in the Ile-de-France region and Marseille. Participants aged >18 were recruited at the place they last slept the night, using a two-stage cluster-sampling design.

Recruitment sites were stratified using ETHOS definitions (Appendix 2). Four strata in Ile-de-France included migrant worker hostels; emergency shelters and centres for asylum seekers; social hostels and similar facilities; and individuals permanently sleeping rough and those in camps or slums. Another stratum was a subsample of a cohort following homeless and migrant populations in Marseille.

We estimated sample size per stratum, based on assumptions reflecting vaccine hesitancy reported in the literature, and considering 80% power, design effects of 3 and 5% accuracy. Total estimated required sample size was 3,751 (Appendix 3).

In Ile-de-France, we built sampling frames for each stratum using data provided by various actors involved with these populations; these listed location and size of each site (Appendix 3). In each stratum, recruitment sites were randomly selected (first stage) proportionally to their size.

Sample size per site was calculated in proportion to expected sites population, with participant sampling in the second stage depending on site type. In shelters, migrant workers’ hostels, and centres for asylum seekers, individuals were selected using simple random sampling when resident lists or room lists existed, and systematic random sampling otherwise. To ensure the selected person was included, sites were visited repeatedly at different times, including weekends and evenings. If individuals were absent or declined to consent, selected individuals were replaced by another sharing that room or the one adjacent.

For people living in the street/camps/slums, we obtained an exhaustive census map recording all homeless and migrants living in subdivisions of Paris in March 2021 [17]. All individuals were systematically invited to participate (exhaustive sampling) until stratum sample size was reached. In cases of refusal, the next person apparent was interviewed.

In Marseille, a local partner supports an exhaustive cohort of PEH/PH. We drew a subsample from this cohort, using simple random sampling, with planned replacements for refusal/absence.

### Outcomes and definitions

Vaccine uptake was the main outcome, defined as uptake of at least one COVID-19 vaccine dose, irrespective of brand or type. Vaccine coverage was defined as a full schedule of COVID-19 vaccine, i.e. at least two doses of messenger RNA vaccine (usually Pfizer), one dose of Janssen vaccine, or one dose of any vaccine following prior COVID-19 infection.

### Data collection

After obtaining verbal informed consent, questionnaires were administered by trained interviewers in participant’s preferred language. Interviews were conducted in French, English, Arabic, Farsi, Spanish, Turkish, Wolof, and Pulaar or were translated by phone in any other language. Responses were recorded using tablets (interview form in Appendix 4). COVID-19 vaccine status was verified via the national “Pass sanitaire” phone app (TousAntiCovid).

Questionnaire topics covered sociodemographics (age, gender, administrative status, native language, duration in France), housing (type of residence in the past 3 months, mobility), participants’ views about vaccines (general and COVID-19-specific), vaccination (status, place, date, reasons for vaccination and non-vaccination), health-related information (history of COVID-19 infection and/or hospitalization, medical coverage), sources of COVID-19 vaccination information (internet, TV/radio, relatives etc), finances and related (work, source of income, source of meals), support and coping mechanisms (food distribution, support organizations), moral and material support from relatives or social workers, health literacy and discrimination. We also collected information on recruitment sites, covering distance to vaccination sites, including those for the general population as well as those dedicated to PEH/PH.

### Grouping for analysis

Sampled populations are mobile, with individuals often staying at a site for just a few days. For the purposes of analysis, we recombined strata into three categories, based on the most reported type of residence over the last three months, irrespective of recruitment location. The three groups comprised “Housed” (individuals renting own accommodation or housed in a migrant workers’ hostel); “Accommodated” (temporarily hosted in asylum seekers’ centres, emergency shelters, or social hostels); and “Streets” (individuals sleeping rough, in camps, in squats or in slums).

### Statistical Analysis

Missing values were imputed when feasible (Appendix 5). Summary measures for main outcomes (vaccine uptake and coverage) were calculated by location, stratum, and participant characteristics, and expressed as estimates with Clopper–Pearson 95% confidence intervals (CI). Descriptive analyses were performed taking into account sampling weights for inclusion probability, and clusters for variance estimation. For univariate and multivariable modelling, unweighted analyses were performed.

Sample vaccine uptake and coverage were compared to the French general population. Weighted direct standardization by age category was performed, using age cut-off: 18, 25, 40, 55, 65, >65; estimates and 95% CI were computed for the overall study population and for each stratum (Appendix 5).

We used univariate logistic regression to explore factors associated with vaccine uptake for all strata combined. We constructed a multilevel multivariable logistic regression model with random intercepts for specific recruitment sites to account for clustering and random effects on several variables after testing for validity. (Appendix 5).

Additional analyses included univariate/multivariate stratified analyses as described above and negative binomial regressions on the total number of vaccinated individuals by site as a count outcome, and site-related variables as covariables (Appendix 5). Data were analysed using Stata v.16 software (StataCorp. 2019. College Station, TX) and R (v3.6.2).

### Ethics

The study protocol was approved by the Comité de Protection des Personnes III, Ile de France, Paris on 13 August 2021 (ref. 2021-A01960-41).

### Roles of the Funding Sources

The study was funded by Santé Publique France, Agence Nationale de Recherches sur le Sida (ANRS-MiE/Capnet)and Agence Régionale de Santé – Ile de France with additional support provided by Médecins Sans Frontières and Société de Pathologie Infectieuse de Langue Française. External donors had no role in the study design, data collection, interpretation, analysis, report writing, or the decision to submit for publication.

## RESULTS

### Study Population

In Ile-de-France, 3,319 people were surveyed at 43 centres for asylum seekers, 45 social hostels, 32 migrant workers hostels and 62 recruitment locations in the streets, camps, or slums. In Marseille, 371 were surveyed in 27 emergency shelters, recruitment locations in the streets and squats (study flow chart in Appendix 6 and maps in Appendix 7). 9,923 individuals were initially selected, of whom 4,124 (41%) were absent and 2,068 (21%) refused to participate or were subsequently excluded.

### Characteristics of study participants

Of the 3,690 surveyed individuals, 855 comprised the “Housed” stratum, 2,321 the “Accommodated” stratum and 514 the “Streets” stratum.

53·7% (95%CI 50·2-57·2) of study participants were male with very few women included in the Housed stratum (4·6%; 95%CI 2·7-6·6). Weighted mean age was 41 years (95%CI: 39·9-41·8). Housed participants were older than those in other strata (22·5% >65y vs 4·4% in Accommodated and 3·5% in Streets respectively; p<0·001). In the Housed stratum, individuals commonly originated from West Africa (48·0%, 95%CI 41·2-54·8) or Central/Southern Africa (26·1%, 95%CI 20·7-31·5); 61·1% had been in France >10 years ((95%CI 55·6-66·5), and 62·5% had official documentation (95%CI 58·1-66·9). In Accommodated stratum, geographic origins were similar (West Africa 43·1%; 95%CI 40-46·2, Central/Southern Africa, 20·1%; 95%CI 17·8-22·4), but 62·3% had been in France <10 years (95%CI 58·8-65·8) and 38·0% were undocumented (95%CI 34·4-41·7), with 31·2% seeking asylum (95%CI 28·4-34·3). 48·2% of the Streets stratum were French or EU citizens (95%CI 39·1-57·1) and this stratum had the highest proportion of recently-arrived migrants (27·7%, 95%CI 20·0-35·5).

Almost all Housed participants (92·8%; 95%CI 90·6-94·9) and the majority of Accommodated participants (63·5%; 95%CI 59·6-67·5) were able to buy their own meals, while 52% of Streets participants could (95%CI 43·9-60·0), and 43% (95%CI 34·5-51·9) of them resorted to panhandling.

While most participants in the Housed stratum (67·4%; 95%CI 60·2-74·6) and half of Streets participants (46·5%; 95%CI 37·7-55·2) reported living alone, most Accommodated participants shared a room with others (78·9%; 95%CI 74·8-82·9). Other variables are described by strata in the Appendix 8 (Table S1).

### Vaccination

76·2% (95%CI 74·3-78·1) of surveyed individuals reported receiving at least one COVID-19 vaccine dose in 2021; 73·1% (95%CI 69·6-76·5) reported receiving a full vaccine schedule. Vaccination status was verified on national electronic certificate for the majority (82·2%; 95%CI 79·8-84·6).

Vaccine uptake varied significantly by stratum (chi^2^ p<0·001) with an overall design-effect (DE) of 1·85 (Figure 2). Uptake was highest (85·6%; 95%CI 83·0-88·2; DE: 2·78) among Housed individuals, followed by Accommodated (75·4%; 95%CI 73·0-77·8; DE: 2·51) and lowest in Streets participants (42·0%; 95%CI 34·3-49·7; DE: 1·08). Overall and for each stratum, vaccine uptake in PEH/PH was delayed by roughly two months as compared to the general population (Figure 1).

**Figure 1.**
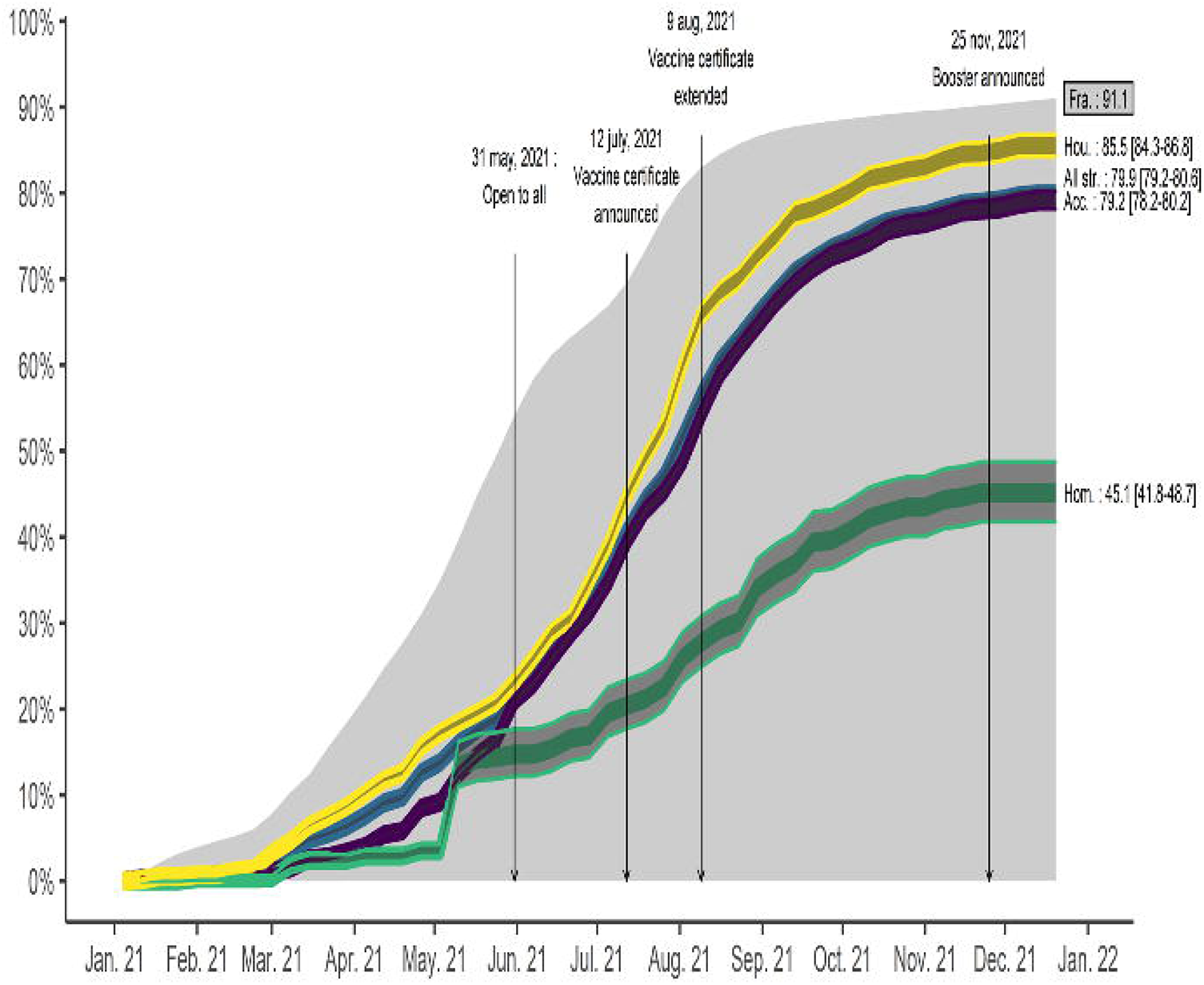
Forest plot for final model

**Figure 2.**
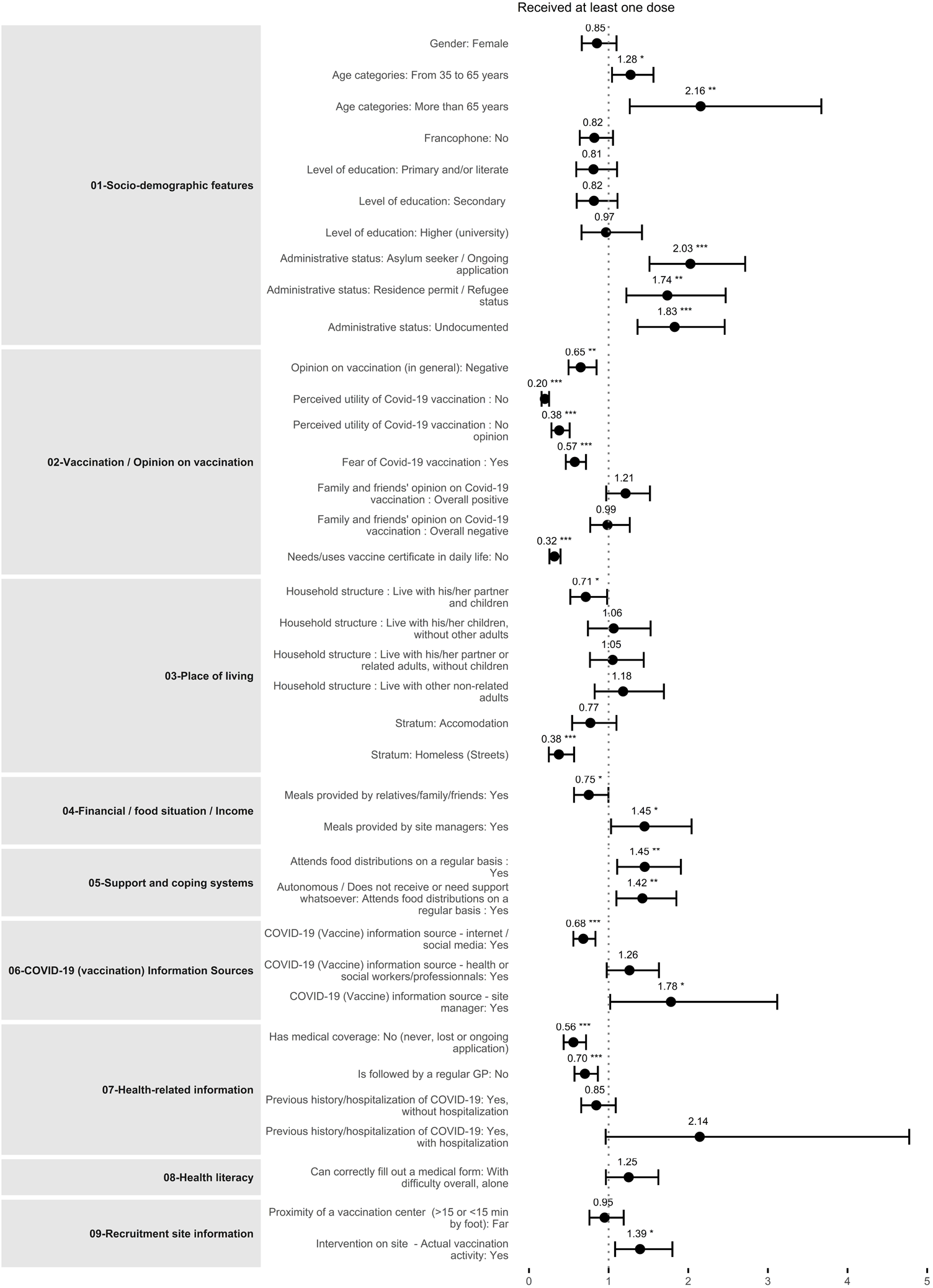
Standardized vaccination rates

Vaccination sites for surveyed individuals included vaccination centres open to all (55·1%; 95%CI 52·0-58·2), followed by healthcare services available to those with medical coverage (drugstores, general practitioners, hospitals;23·9%; 95%CI 20·3-27·5) and more rarely outreach/onsite vaccination activities targeting PEH (18·0%; 95%CI 15·2-20·9).

Reasons for vaccination did not differ across strata. Most participants reported accepting vaccination to protect themselves (63·0%; 95%CI 60·3-65·6), to protect vulnerable relatives (24·9%; 95%CI 21·6-28·2) or as their civic duty to protect everyone (32·7%; 95%CI 29·2-36·2). Many participants reported feeling compelled to accept vaccination (44·1%; 95%CI 41·4-46·8), either to keep their job, to travel abroad, or to obtain a vaccine certificate; 23·9% (95%CI 21·3-26·5) declared the vaccine certificate as the main reason for vaccination.

Half of non-vaccinated individuals reported having no intent to be vaccinated in the future (50·3%; 95%CI 54·7-55·0) but these comprised the majority in the Streets stratum (71·5%; 95%CI 62·3-80·7). Reasons for non-vaccination were generally linked with refusal and hesitancy (all strata: 75·6%; 95%CI 71·7-79·5), more than physical or practical barriers (all strata: 24·4, 95%CI 20·5-28·3). Housed and Accommodated strata did not differ in terms of specific reasons, with fear of immediate side effects being predominant (53·9%; 95%CI 42·5-65·3 and 59·7%; 95%CI 53·5-65·9, respectively) followed by fear of injection/fear of serious disease (43·6%; 95%CI 32·8-54·5 and 50·9%; 95%CI 44·2-57·6, respectively), and scepticism about vaccine effectiveness/utility (29·3%; 95%CI 18·5-40·1 and 25·5%; 95%CI 19·1-31·9, respectively). In the Streets stratum, participants were more subject to conspiracy theories/denial of the crisis (28·7%; 95%CI 18·5-38·9) and were more likely to be influenced by peers (31·9%; 95%CI 19·7-44·2]).

### Drivers of first dose intake

Factors associated with vaccine uptake in univariate analysis are summarized in Appendix 9 (table S2). Stratum is strongly associated with vaccination (p<0·0001). Sociodemographic characteristicss, opinions on vaccination, need for vaccine certification, food security, support and coping mechanisms, COVID-19 information sources, trust in authorities, health-related variables, and site-related variables were all candidates for multivariable analysis (p-values<0·001).

Table 3 and Figure 2 summarise results of the final multivariable model after backwards selection. Odds of vaccine uptake varies by stratum: compared to Housed, Accommodated did not differ, but Streets individuals were less likely to be vaccinated (AOR 0·38, 95%CI 0·25-0·57, p<0·001).

**Table 1.**
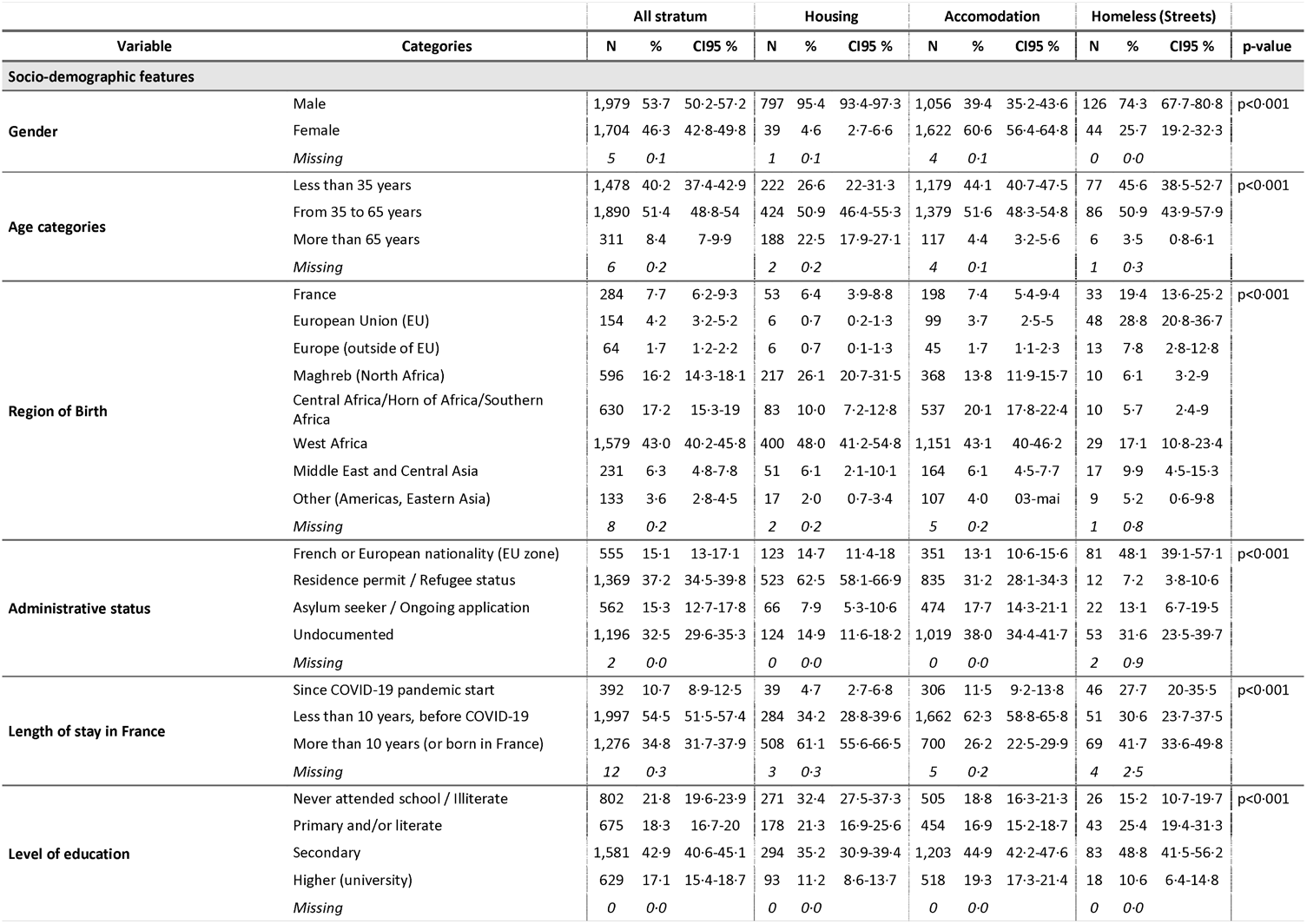
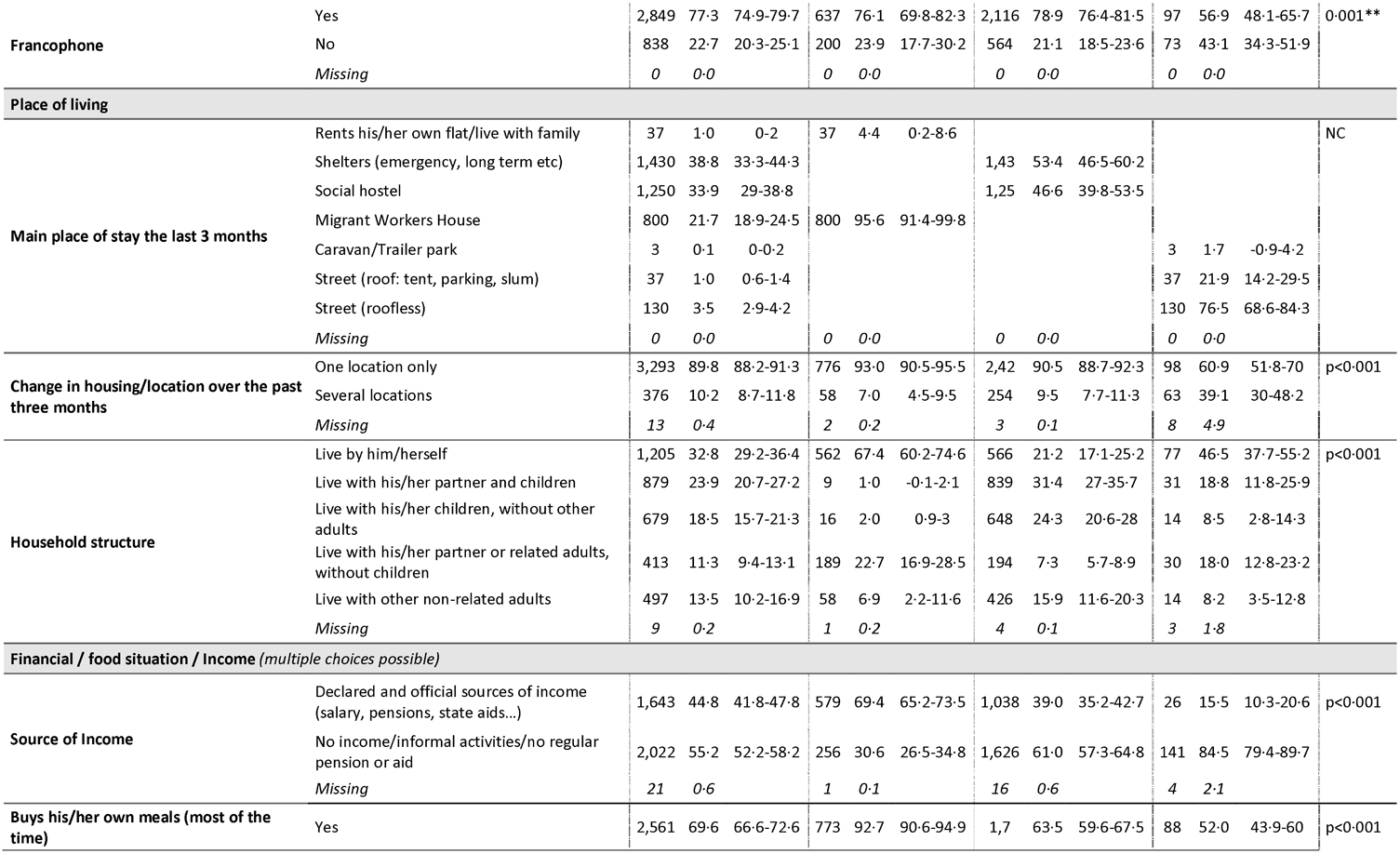
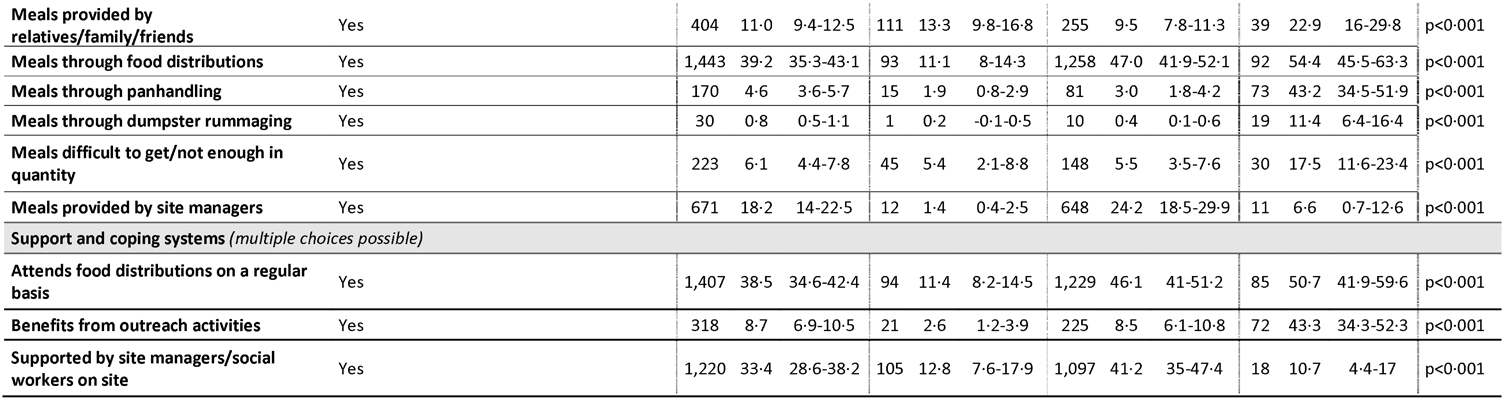
Participants characteristics (global and by stratum)

**Table 2.**
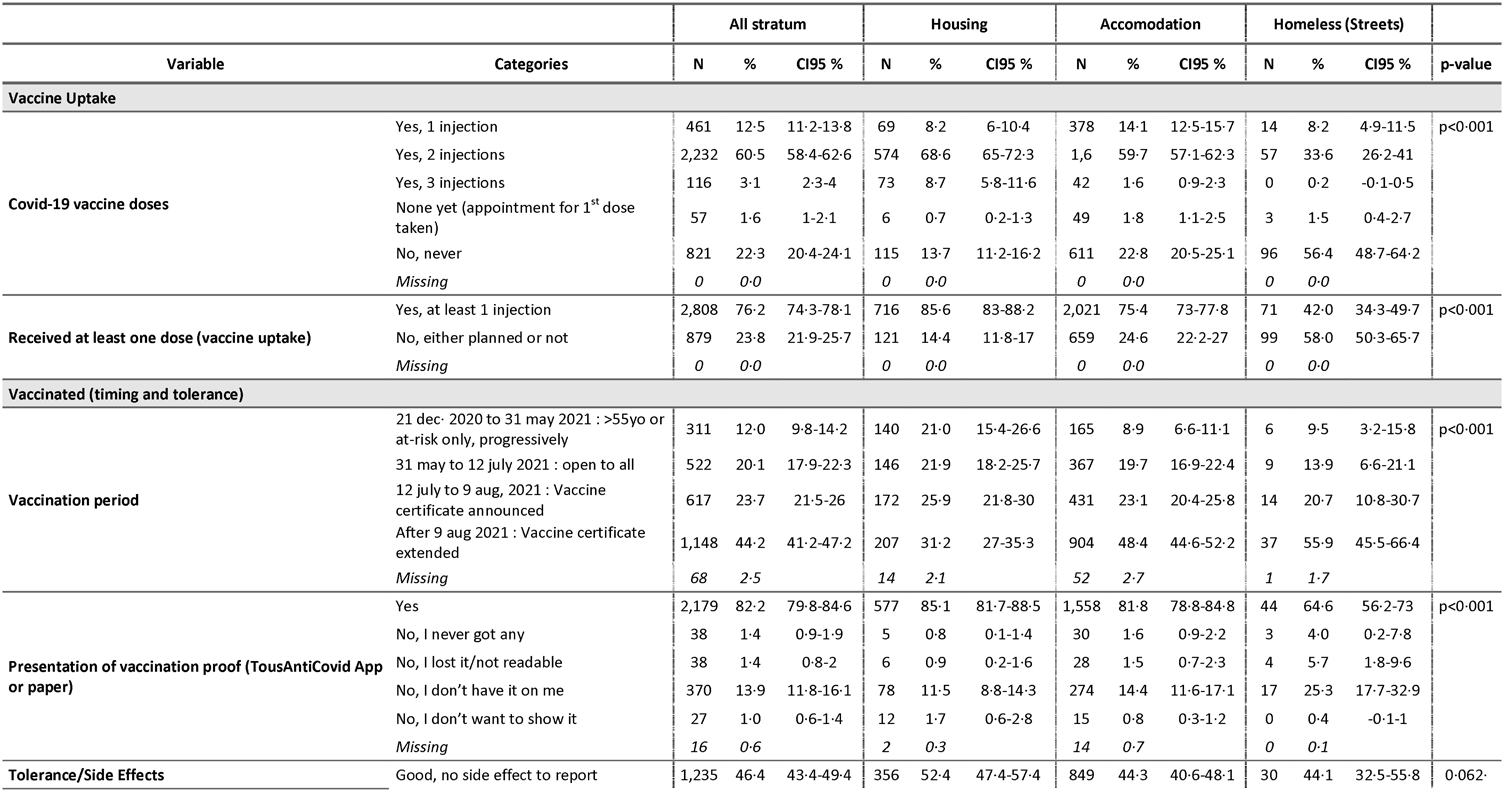
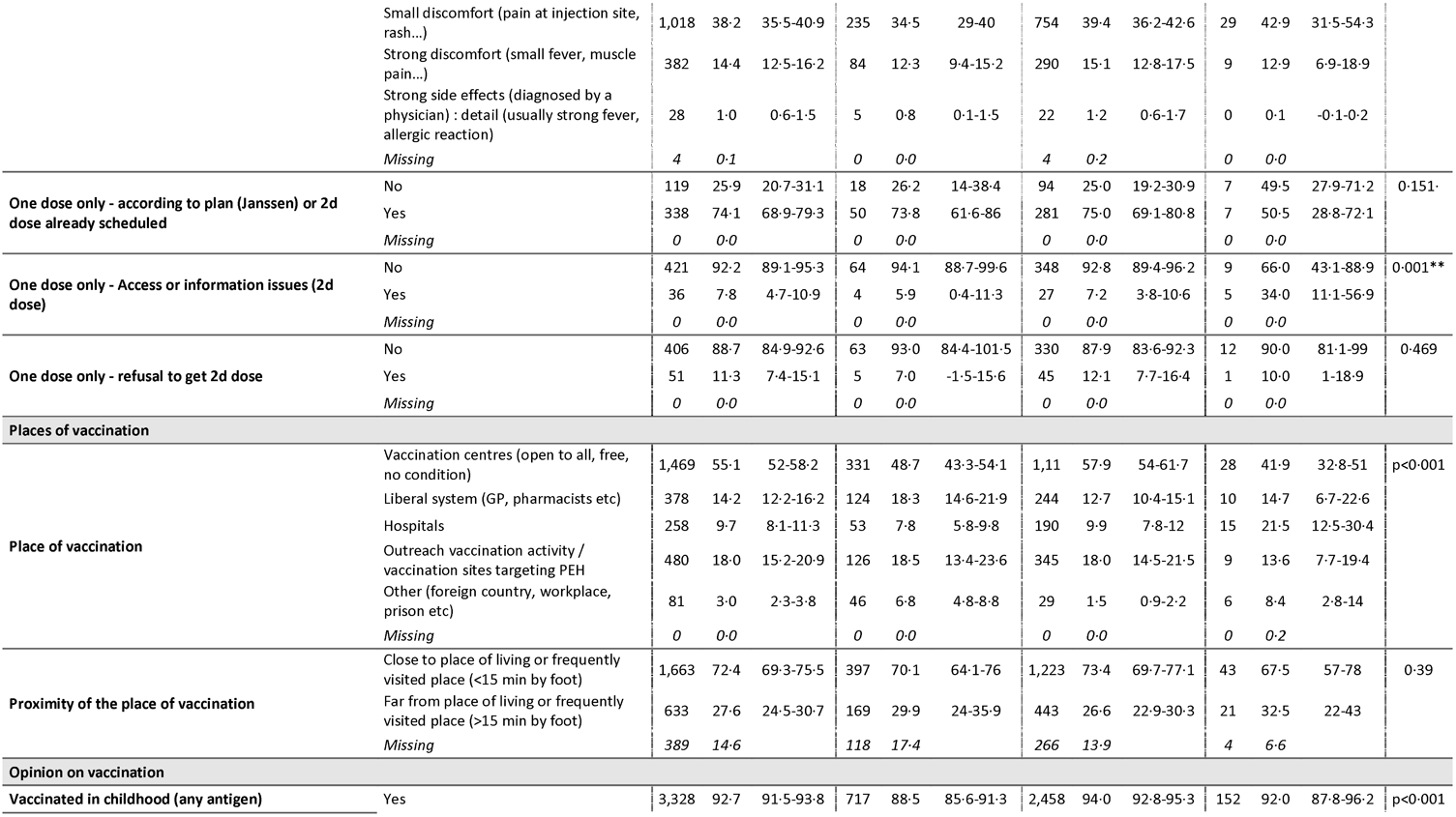
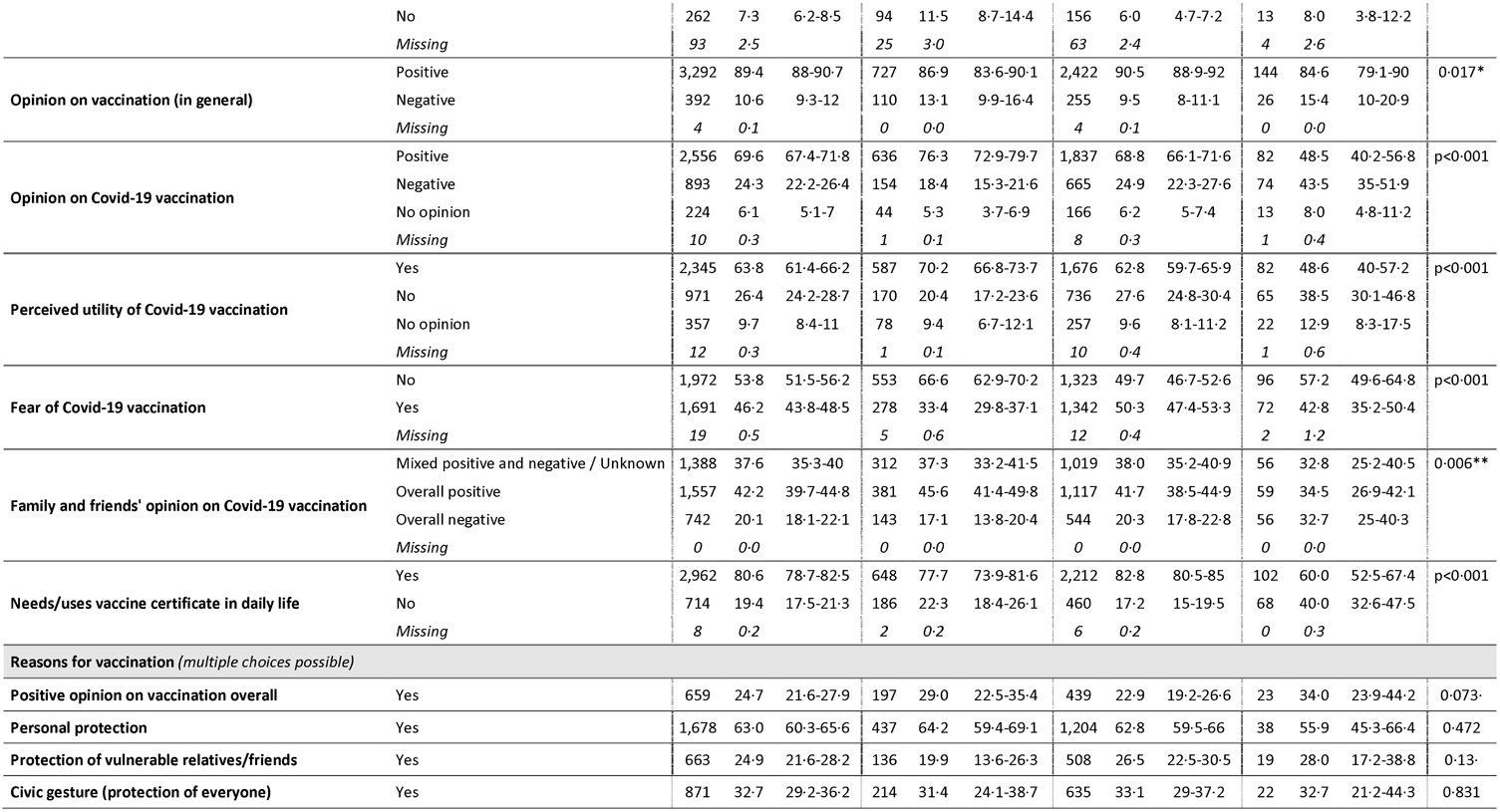
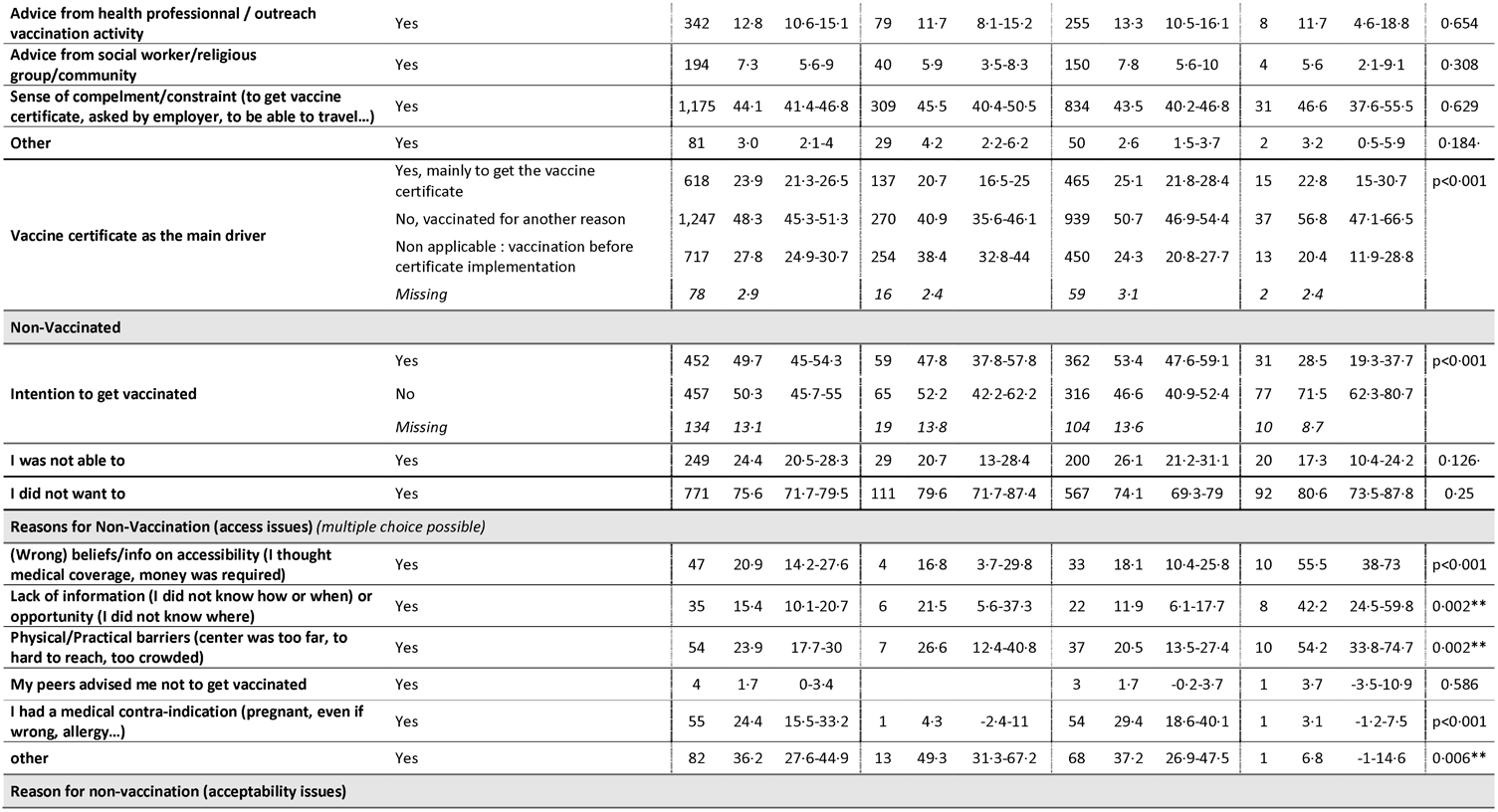
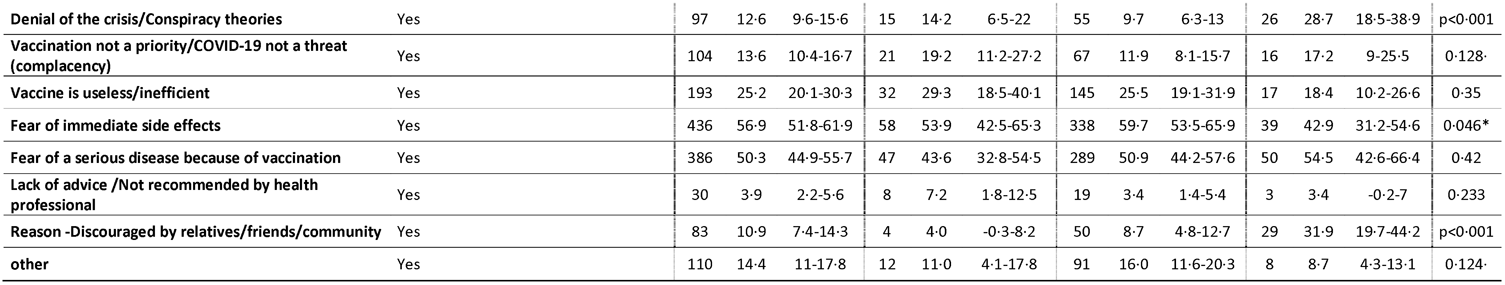
Vaccination characteristics (global and by stratum)

**Table 3.** Drivers of vaccine uptake: multivariable analysis. Final multivel mixed logistic model (imputed missing values)

| *Predictors* | At least one dose of Covid-19 vaccine |  |  |
| --- | --- | --- | --- |
|  | *Adjusted Odds Ratio* | *CI* | *p* |
| (Intercept) | 7.9 | 4.4 – 14.3 | **<0.001** |
| Gender: Male | *Reference* |  |  |
| Gender: Female | 0.9 | 0.7 – 1.1 | 0.218 |
| Age categories: Less than 35 years | *Reference* |  |  |
| Age categories: From 35 to 65 years | 1.3 | 1.0 – 1.6 | **0.018** |
| Age categories: More than 65 years | 2.2 | 1.3 – 3.7 | **0.005** |
| Francophone: Yes | *Reference* |  |  |
| Francophone: No | 0.8 | 0.6 – 1.1 | 0.122 |
| Level of education: Never attended school / Illiterate | *Reference* |  |  |
| Level of education: Primary and/or literate | 0.8 | 0.6 – 1.1 | 0.184 |
| Level of education: Secondary | 0.8 | 0.6 – 1.1 | 0.197 |
| Level of education: Higher (university) | 1.0 | 0.7 – 1.4 | 0.863 |
| Administrative status: French or European nationality (EU zone) | *Reference* |  |  |
| Administrative status: Asylum seeker / Ongoing application | 2.0 | 1.5 – 2.7 | **<0.001** |
| Administrative status: Residence permit / Refugee status | 1.7 | 1.2 – 2.5 | **0.002** |
| Administrative status: Undocumented | 1.8 | 1.4 – 2.5 | **<0.001** |
| Household structure : Live by him/herself | *Reference* |  |  |
| Household structure : Live with his/her partner and children | 0.7 | 0.5 – 1.0 | **0.037** |
| Household structure : Live with his/her children, without other adults | 1.1 | 0.7 – 1.5 | 0.743 |
| Household structure : Live with his/her partner or related adults, without children | 1.1 | 0.8 – 1.4 | 0.762 |
| Household structure : Live with other non-related adults | 1.2 | 0.8 – 1.7 | 0.364 |
| Stratum: Housing | *Reference* |  |  |
| Stratum: Accommodation | 0.8 | 0.5 – 1.1 | 0.150 |
| Stratum: Homeless (Streets) | 0.4 | 0.2 – 0.6 | **<0.001** |
| Meals provided by relatives/family/friends: No | *Reference* |  |  |
| Meals provided by relatives/family/friends: Yes | 0·8 | 0·6 – 1·0 | **0·048** |
| Meals provided by site managers: No | *Reference* |  |  |
| Meals provided by site managers: Yes | 1·5 | 1·0 – 2·0 | **0·033** |
| Attends food distributions on a regular basis : No | *Reference* |  |  |
| Attends food distributions on a regular basis : Yes | 1·5 | 1·1 – 1·9 | **0·007** |
| Autonomous / Does not receive or need support whatsoever: Attends food distributions on a regular basis : No | *Reference* |  |  |
| Autonomous / Does not receive or need support whatsoever: Attends food distributions on a regular basis : Yes | 1·4 | 1·1 – 1·9 | **0·008** |
| Opinion on vaccination (in general): Positive | *Reference* |  |  |
| Opinion on vaccination (in general): Negative | 0·6 | 0·5 – 0·8 | **0·002** |
| Perceived utility of Covid-19 vaccination : Yes | *Reference* |  |  |
| Perceived utility of Covid-19 vaccination : No | 0·2 | 0·2 – 0·3 | **<0·001** |
| Perceived utility of Covid-19 vaccination : No opinion | 0·4 | 0·3 – 0·5 | **<0·001** |
| Fear of Covid-19 vaccination : No | *Reference* |  |  |
| Fear of Covid-19 vaccination : Yes | 0·6 | 0·5 – 0·7 | **<0·001** |
| Family and friends' opinion on Covid-19 vaccination : Mixed positive and negative / Unknown | *Reference* |  |  |
| Family and friends' opinion on Covid-19 vaccination : Overall positive | 1·2 | 1·0 – 1·5 | 0·092 |
| Family and friends' opinion on Covid-19 vaccination : Overall negative | 1·0 | 0·8 – 1·3 | 0·910 |
| Needs/uses vaccine certificate in daily life: Yes | *Reference* |  |  |
| Needs/uses vaccine certificate in daily life: No | 0·3 | 0·3 – 0·4 | **<0·001** |
| Has medical coverage: Yes (national social security, state medical aid) | *Reference* |  |  |
| Has medical coverage: No (never, lost or ongoing application) | 0·6 | 0·4 – 0·7 | **<0·001** |
| Is followed by a regular GP: Yes | *Reference* |  |  |
| Is followed by a regular GP: No | 0·7 | 0·6 – 0·9 | **0·001** |
| Previous history/hospitalization of COVID-19: No | *Reference* |  |  |
| Previous history/hospitalization of COVID-19: Yes, without hospitalization | 0·8 | 0·7 – 1·1 | 0·193 |
| Previous history/hospitalization of COVID-19: Yes, with hospitalization | 2·1 | 1·0 – 4·8 | 0·062 |
| Can correctly fill out a medical form: With ease overall, alone | *Reference* |  |  |
| Can correctly fill out a medical form: With difficulty overall, alone | 1·3 | 1·0 – 1·6 | 0·091 |
| COVID-19 (Vaccine) information source - internet / social media: No | *Reference* |  |  |
| COVID-19 (Vaccine) information source - internet / social media: Yes | 0·7 | 0·6 – 0·8 | **<0·001** |
| COVID-19 (Vaccine) information source - health or social workers/professionals: No | *Reference* |  |  |
| COVID-19 (Vaccine) information source - health or social workers/professionals: Yes | 1·3 | 1·0 – 1·6 | 0·076 |
| COVID-19 (Vaccine) information source - site manager: No | *Reference* |  |  |
| COVID-19 (Vaccine) information source - site manager: Yes | 1·8 | 1·0 – 3·1 | **0·043** |
| Proximity of a vaccination center (>15 or <15 min by foot): Close | *Reference* |  |  |
| Proximity of a vaccination center (>15 or <15 min by foot): Far | 0·9 | 0·8 – 1·2 | 0·654 |
| Intervention on site - Actual vaccination activity: No | *Reference* |  |  |
| Intervention on site - Actual vaccination activity: Yes | 1·4 | 1·1 – 1·8 | **0·011** |

### Sociodemographic drivers

Odds of vaccine uptake increased with age (AOR for age 35-65 vs 18-35; 1·3, 95%CI 1·0-1·5, AOR for >65 vs 18-35, 2·1; 95%CI 1·3-3·7). Undocumented participants were more likely to be vaccinated than those with French or EU citizenship (AOR=1·8, 95%CI 1·3-2·4), as were those with residence permits or refugee status (AOR=1·7, 95%CI 1·2-2·4) and asylum seekers (AOR=2·0, 95%CI 1·5-2·7). Vaccine uptake odds were lower for participants living with family (AOR=0·7, 95%CI 0·5-1·0).

### Economic drivers

Odds of vaccination were higher where participants described provision of meals by a site manager (AOR 1·5, 95%CI 1·0-2·0), for participants dependent on food distribution (AOR 1·5, 95%CI 1·1-1·9), and conversely, where participants felt independent in terms of money and food (AOR 1·4, 95%CI 1·1-1·9). Odds were lower if meals were provided by family/friends (AOR 0·8, 95%CI 0·6-1·0).

### Source of COVID-19 information

Vaccine uptake odds were significantly higher where COVID-19 information was obtained from site managers (AOR 1·8, 95%CI 1·0-3·1). Conversely, uptake was lower were where the main source of information was the internet or social media (AOR 0·7, 95%CI 0·6-0·8).

### Vaccine Certificate and Health

Vaccine uptake odds were lower for those who never needed or used a vaccine certificate (AOR 0·3, 95%CI 0·2-0·4) and in participants with no medical coverage (AOR 0·6, 95%CI 0·4-0·7). Individuals with no regular general practitioner were less likely to be vaccinated (AOR 0·7, 95%CI 0·6-0·9).

### Opinions on vaccination

Participants with negative opinions about vaccines in general were less likely to be vaccinated (AOR 0·6, 95%CI 0·5-0·8), as were participants with negative perceptions of vaccine utility (AOR=0·2, 95%CI 0·1-0·3) or participants afraid of the vaccine or its effects (AOR=0·6, 95%CI 0·4-0·7).

### On-site activities

Participants living in settings with on-site vaccination activities were more likely to be vaccinated (AOR=1·4, 95%CI 1·1-1·8).

For results of negative binomial analysis at site level and stratified multivariable analyses, see Appendices 10 to 12 (Tables S3, S4 and S5).

## DISCUSSION

To our knowledge, few studies exist reporting COVID-19 vaccine uptake in PEH. One study examines uptake among US military veterans attending homelessness centres; this was conducted much earlier in the vaccination campaign, and reports that 46% of homeless veterans were vaccinated in August 2021, as compared to 51% of all veterans [18]. Two recent studies in Ontario, Canada, and Denmark used database mining to derive coverage estimates for homeless individuals, which were similar to those in our Streets stratum [19, 20].

Our study highlights that PEH/PH are less likely to receive COVID-19 vaccination than the general population (79% vs 91%). This aligns with evidence for other vaccines: 24% of immigrants to London were vaccinated against influenza vs 53% of UK-born participants [10]; 17% of Italian immigrants were vaccinated against influenza vs 40% of Italy-born citizens [11].

Our survey underlines that vaccination receipt varies according to precariousness and social integration: those living on the street, in camps or squats are much less likely to be vaccinated (42% having at least one dose), than Accommodated (75%) and Housed participants (85%).

We also aimed to describe and understand determinants of COVID-19 vaccination in PEH/PH. Existing evidence has assessed coverage in the general population in relation to income and/or ethnicity [21] or has focused on vaccine intention or hesitancy in PEH [22, 23]. However, little evidence reports on drivers of uptake in the type of population we have studied.

We report multiple factors linked with vaccine uptake, both individual-specific, and externally-determined. Some have been detailed for other vaccine types in PEH [8, 10, 12] and migrants [9, 11, 13, 14].

Factors we identify as associated with vaccination include age, with differences in vaccine uptake between older and younger adults potentially explained by greater fears around disease risks for the elderly [24, 25]. The introduction of vaccine certificates in July 2021 was intended to mitigate such effects and encourage younger age groups to uptake vaccination. Recent studies have since questioned the impact of such policies, which might explain the plateau observed in all strata and in the general population [26]. Thus, differences in vaccine uptake between strata cannot be solely explained by the introduction of certificates.

Practical or physical obstacles to vaccine uptake are rare, as compared to personal motivations. Vaccine hesitancy or negative views on vaccination were the main factors associated with lower coverage. Individuals opposed to vaccination comprised a minority in our sample (12% of non-vaccinated), as compared to hesitant people, with 54% afraid of vaccine effects.

Our data illustrates that vaccinated participants hold a variety of beliefs and behaviours; a majority were convinced the vaccine is useful (64%) and protects (66% vaccinated for this reason), but a non-negligible proportion hold sceptical views or felt compelled to accept vaccination; these findings confirm other work on vaccine intentions in PEH [22, 23, 27, 28].

The effects of peer pressure, reflecting influence of friends, relatives, and others on vaccination intentions is well-described in the literature [24, 25, 27, 28]. Our findings are consistent with others on general vaccine intention among migrants [13, 28], including some data for COVID-19 [22, 23].

These data underline the importance of awareness-raising and sensitization by trusted third parties such as social workers, mediators, and site managers, in line with qualitative studies [22, 23]. Our data also show the positive impact on vaccine uptake by support mechanisms and follow-up by medical professionals and/or by social organizations. Such factors are rarely reported in the literature [27, 28], although some data documents the effects of lack of support from health personnel or even their negative influence on vaccination [28, 29].

We also report on structural barriers to vaccine uptake, including distance/time to vaccination centres, lack of information on locations/dates, and problems making online appointments. The more minor role played by these factors may reflect national policies ensuring free vaccination, regardless of medical coverage or administrative status. Moreover, deployment of mobile teams and specialised site-based vaccination activities may have helped to reach those with lower access to care.

Participants in our sample reported lacking awareness of such strategies or misjudged their rights, with many mistakenly believing they lacked entitlement. Notably, we found that those with medical cover, registered with a GP, or who had recourse to the healthcare system, were more likely to access vaccination, in line with prior work on COVID-19 [27, 29] and other conditions [28]. Misinformation due to rumours, and defiance towards authorities reinforce these barriers, and are well-known in PEH [22, 23, 24, 27, 28, 29, 30].

### Strengths and Limitations

Study strengths include efforts to ensure rigorous methodology, including use of face-to-face survey (rare in PEH studies), conducted in eight of the most commonly used languages in this population. Sampling approaches involved every effort to ensure representativeness, with a two-stage cluster survey, large sample size, and simple random sampling to ensure exhaustive coverage of subpopulations, as well as stratification based on the ETHOS classification to fully represent housing types.

One limitation is the high replacement rate, either because of absence or refusal. In the Accommodated stratum, the proportion of women was higher than expected and vaccine uptake therefore may be underestimated, since women are less vaccinated overall (72·4% vs 79·4%, p<0·001; Table S1, Appendix 6). In the Housed stratum, elderly and/or retired people were also over-represented compared to people of working age, despite efforts to visit sites outside work hours. This would likely lead to overestimation of vaccine uptake, given that elderly people are generally better vaccinated overall. Language barriers may also have contributed to increased refusals.

Finally, limitations characterizing cross-sectional studies may also apply, specifically the potential for social desirability bias in responses (relating to support received and reasons for vaccination or non-vaccination), and survival or healthy worker bias; only those present and in good health could be interviewed.

## Conclusion

We found that COVID-19 vaccine uptake and coverage are lower for this PEH/PH population. Coverage was higher amongst those with access to the common law system and/or accompanied or supported by associations. The national strategy relating to PEH/PH seemed to be reaching those housed or accommodated, but substantial effort is still needed to reach the most excluded, street-sleeping individuals.

Our findings have implications for policy in relation to these vulnerable groups. Outreach activities and on-site vaccination programs should be extended and tailored to targeted subgroups. Sensitization activities involving field actors who work closely with PEH/PH populations should take place early in such vaccination campaigns to address barriers like vaccine hesitancy and complacency. Our study reveals that high levels of vaccination can be obtained even in these vulnerable groups; higher uptake amongst the “Housed” and “Accommodated” strata imply that policies ensuring free, universal access to vaccination and the support of field actors can ensure coverage.

This work highlights the role of determinants relating to social integration and housing in relation to vaccination. The overriding influence of housing insecurity, especially, suggests that policies prioritizing secure housing first, in order to address health and social needs, may have value. These insights may be applicable to activities and planning for future pandemics but remain valid well beyond the COVID-19 pandemic.

## Data Availability

All data produced in the present study are available upon reasonable request to the authors.

## Contributors

TR, BM and SV conceived the study (literature search, study design, etc). TR, BM, JS, MM and SV developed the study protocol. TR, BM, JS, MM and CV performed field data collection (and data questionnaires) and supervised the field study. TR and GL performed data management and statistical data analysis. TR, BM and SV performed literature search for the manuscript. GL and TR verified the underlying data and performed additional analyses. All authors interpreted the results, contributed to writing the manuscript, and approved the final version for submission.

## DATA SHARING

Anonymized data collected for the study and a data dictionary will be made available to other researchers following approval of a study proposal by TR for 5 years from publication. The study protocol, statistical analysis plan and electronic forms are also available from TR.

## ACKNOWLEDGEMENTS

Authors want to thank all the participants to the survey, as well as the managers of the hostels and emergency shelters who made the survey possible.

Authors would also like to thank partner organizations for sharing their data and allowing us to constitute sampling frames: RATP, Ville de Paris (UASA), DIHAL, SAMU Social de Paris, France Terre d’Asile, Emmaüs Solidarité and Prospective & Coopération.

External donors had no role in the study design, data collection, interpretation, analysis, report writing or the decision to submit for publication.

We acknowledge the support of Emma Veitch, medical editor for MSF, in providing editorial assistance; her work was funded by MSF-USA.

