## Supplementary File for "Drivers and prevalence of COVID-19 vaccine uptake among homeless and precariously housed people in France: a cross-sectional population-based study"

### Supplementary Materials

Table of Contents[_Toc107069989](#_Toc107069989)

#### Appendix 1. National Vaccination Strategy

Source Ministry of Health (in French) : <https://solidarites-sante.gouv.fr/grands-dossiers/vaccin-covid-19/je-suis-un-particulier/article/foire-aux-questions-la-strategie-de-vaccination-et-le-calendrier> )

OBJECTIVES AND PRINCIPLES OF THE VACCINATION STRATEGY

The vaccine strategy put in place follows three public health objectives:

- To reduce mortality and severe forms of the disease
- To protect caregivers and the health care system
- To guarantee the safety of vaccines and vaccination

The vaccination should be non-mandatory, free of charge and without requirements (medical coverage, citizenship etc) and must be the safest possible.

Vaccination Announcements Timeline :

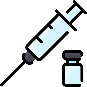

**27 Jan 2020**

**Launch of the campaign: all nursing homes residents, at-risk individuals, health prof. working in hospitals and nursing homes**

**18/01/2021**

Vaccination opened to **75y and older** and **vulnerable people**

**06/02/2021**

Vaccination opened to **all health and social professionals**

**and firefighters regardless of age**

**19/02/2021**

Vaccination opened to **50y and older with comorbidities**

**25/02/2021**

**General practitioners allowed to vaccinate**

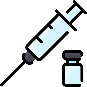

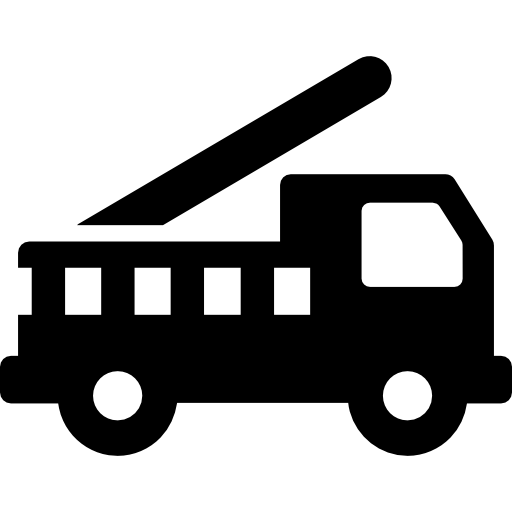

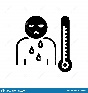

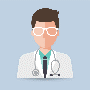

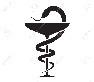

**15/03/2021**

**City drugstores and pharmacists allowed to vaccinate**

**08/04/2021**

Vaccination opened to **pregnant women (>= 2d trimester) regardless of age**

**12/04/2021**

Vaccination opened to **55y and older**

**01/05/2021**

Vaccination opened to **18y and older with comorbidities**

**12/05/2021**

Vaccination opened to **18y and older if doses are available in vaccination centers**

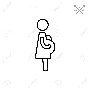

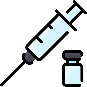

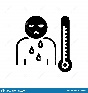

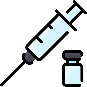

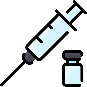

**31/05/2021**

Vaccination opened **to 18y and older**

**09/06/2021**

Introduction of **Pass Sanitaire for 18y and older to access hospitals and to travel**

**15/06/2021**

Vaccination opened to **12y and older**

**06/07/2021**

President Macron announcement:

**Mandatory Pass Sanitaire for 12y and older**

**Extension to leasure and cultural venues**

**Mandatory vaccination for health professionals**

**PCR and antigenic tests not free anymore for non-vaccinated**

**21/07/2021**

Pass Sanitaire **now mandatory to access bars, pubs, concert and sport venues with a capacity >=50 000**

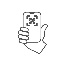

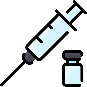

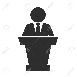

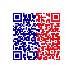

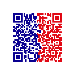

**09/08/2021**

Pass Sanitaire now mandatory to **access all leisure and cultural venues, to access large malls (>=20 000 m²) and to visit hospitals and nursing homes**

**01/09/2021**

**Booster dose (Pfizer/Moderna only) for vulnerable people**

**30/09/2021**

**Pass Sanitaire extended to 12y and older**

**05/10/2021**

**Booster dose for health professionals**

**27/11/2021**

**Booster** dose extended **to the entire population aged 18y and older**

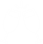

**15/12/2021**

Vaccination opened to **children aged 5 to 11 y with comorbidities or living with at-risk persons**

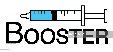

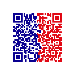

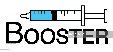

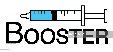

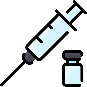

#### Appendix 2. Definitions

European Typology of Homelessness and Housing ExclusionTypology according to <https://www.feantsa.org/en/toolkit/2005/04/01/ethos-typology-on-homelessness-and-housing-exclusion>

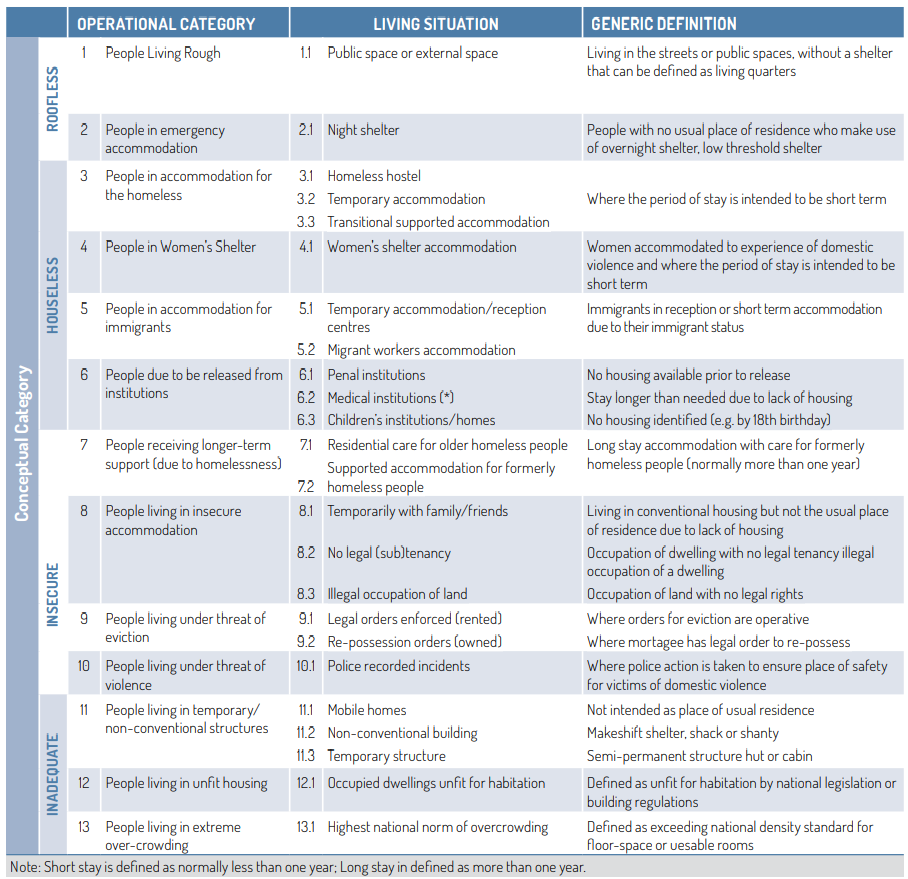

**French system : Types of facilities for roofless, homeless and migrant populations**

Extracted from : Fondation Abbe Pierre· l’État du mal-logement en France 2021 : <https://www.precarite-energie.org/wp-content/uploads/2022/02/reml2022-web.pdf>

Homelessness in France (**ETHOS Operational category**)

-People living in the streets, parking lots, under the bridges or in informal settlements/tents (roofless), can sometimes spend the night in overnight emergency shelters (Accueil de Nuit) and have a shower/meal in daily emergency centers (Accueil de Jour) (**ETHOS 1 and 2**)

-People living in make-shift housing / slums : people often grouped by community/country of origin (e·g· Roma community) (**ETHOS 11**)

-People living in squats and unofficial community dwellings (**ETHOS 8**)

- People hosted in collective/social/solidarity accommodations : managed by social or religious organizations (**ETHOS 3 and 4**)

- People hosted in social hotels : managed by the Samu Social organization. People who need a temporary stay have to call the 115 phone number and will be oriented to an accomodation, usually a real hotel dedicated to hosting precarious populations (**ETHOS 3** for emergency sheltering, **ETHOS 7** for transition accomodation)

- Migrants and asylum seekers hosted in accommodation for immigrants (Dispositif National d’Accueil) (**ETHOS 5**) :

(a) Centres for asylum seekers (CADA); (b) Emergency accommodation for asylum seekers (HUDA, AT-SA, PRAHDA, CAO); (c) Reception and administrative status examination centres (CAES)·

- Migrants and homeless hosted in Emergency Shelters for Homeless People (CHU) and Centers for Social Re-Integration (CHRS) (**ETHOS 3**)

- Undocumented migrants awaiting for papers upon arrival on french soil or under deportation from France orders can be detained in retention centers (usually in airports) ·(CRA) (**ETHOS 9**)

Homeless people can switch from any facility to any other (volatile and unstable situation) several times for a long time.

Migrant Workers Hostels (**ETHOS 12 and 13**): initiated by the French state; typically hosting migrant workers. Their original purpose (1950s and 1960s·) was two-fold: as a mean of monitoring a suspect foreign male population at a time of decolonization and workers’ struggles, and as a short-term housing solution for a supposedly temporary migrant labour force. MWH are often managed by state agencies, and sometimes private organizations.

#### Appendix 3. Sample Size Calculation and Sampling Frames

|  | CFA/ES  Ile-De-France | Social Hostels  Ile-De-France | MWH  Ile-De-France | Street/Camps/Squats  Paris | COVID Homeless Cohort  Marseille |
| --- | --- | --- | --- | --- | --- |
| Null Hypothesis (%) | 55 | 55 | 55 | 60 | 60 |
| Power (β, in %) | 80 | 80 | 80 | 80 | 80 |
| Type I error (α, in %) | 5 | 5 | 5 | 5 | 5 |
| Total population (sampling frame) | 45,000 | 32,000 | 23,000 | 5,000 | 1,200 |
| Accuracy (%) – Bilateral test | 5 | 5 | 5 | 5 | 5 |
| Design Effect* | 3 | 3 | 3 | 1 | 1 |
| **Total participants to include** | **1000** | **1000** | **1000** | **384** | **367** |
| Expected Non-Response/Refusals (%) | 20 | 20 | 20 | 33 | 33 |
| Number of clusters | 50 | 50 | 50 | 24 | 31 |

CFA/ES : Centres for Asylum Seekers/Emergency Shelters for homeless

MWH : Migrant Workers Hostels

COVIDHomeless Cohort : cohort of homeless and accomodated migrants followed up for 2 years in Marseille

Assumptions for null hypotheses were made according to published literature on vaccine hesitancy in general French population (*Schwarzinger M, Watson V, Arwidson P, Alla F, Luchini S. (2021). COVID-19 vaccine hesitancy in a representative working-age population in France: a survey experiment based on vaccine characteristics. Lancet Public Health. avr 2021;6(4):e210‑21* ) and in PEH/PH (*Longchamps, C., Ducarroz, S., Crouzet, L., Vignier L. et al. (2021). COVID-19 vaccine hesitancy among persons living in homeless shelters in France. Vaccine, xxxx.* [*https://doi.org/10.1016/j.vaccine.2021.05.012*](https://doi.org/10.1016/j.vaccine.2021.05.012).).

Sampling Frames

Sampling frames were usually obtained through partners and updated for each of the survey strata during the preparation phase between September and November 2021.

CFA/ES :

The complete list of facilities was constructed from the FINESS (Fichier national des établissements sanitaires et sociaux) national database of the Direction de la recherche, des études, de l'évaluation et des statistiques (DREES), which lists all health and social facilities, including Centres d'Hébergement d'Urgence (CHU), Centres d'Hébergement et de Réinsertion Sociale (CHRS), CPH, CADA/HUDA, and other CAO. This was corrected and amended by the APUR database, on which we relied in the framework of a partnership. We then contacted the main social organizations (France Terre d'Asile - FTDA, CRF, Aurore, etc.) and operators/landlords (Adoma/CDC-Habitat, Coallia and ADEF) to ensure that the list was up to date.

Social Hotels (Samu Social)

For this stratum, a partnership via a data transfer agreement was signed with the Samu Social de Paris. The SIAO 75 then sent us the complete and updated list of all the hotels, centers and residences managed by the Pôle 115 and the Pôle Habitat of Samu Social.

MWH

The complete list of MWHs was extracted from the FINESS database. More and more FTMs are being transformed into social residences, so it was important to check the accuracy of the data with the main social landlords (Coallia, ADEF and Adoma/CDC-Habitat).

Street/Camps/Slums/Squats/Subway

This stratum is by definition very heterogeneous and official data is sorely lacking.

The City of Paris, in partnership with APUR and a large number of social organizations, has been organizing a census overnight called « Solidarity Night » for several years, aiming to exhaustively count all the people living on the streets in Paris, along the Seine banks, ring road embankments and the Vincennes and Boulogne parks. The covered area is divided into 320 districts that 1900 volunteers walk through in one night to interview all the people they meet in the streets, parking lots, bus shelters, parks and gardens, train and subway stations. APUR graciously sent us the complete dataset of the last Solidarity Night (25 March 2021).

In parallel, data on informal campsites in Paris and along the ring road were provided by the Unité d'Assistance aux Sans-Abris (UASA) of the City of Paris. Data for the camps in the cities bordering Seine-Saint-Denis were provided by the France Terre d'Asile - FTDA association.

The Régie Autonome des Transport Parisien (RATP) conducts a daily census of homeless people living in the subway corridors and stations. Its social data collection team shared the latest data for the preparation of the survey, and was then a direct operational partner in the survey implementation in subway stations.

Finally, the Délégation Interministérielle à l'Hébergement et à l'Accès au Logement (DIHAL) has been in charge of slum clearance since 2018. In particular, it lists all the informal sites and slums in France, and compiles this information on a platform that can be accessed free of charge upon request. The list of all slums and informal settlements in Ile-de-France has thus been extracted during preparation phase.

Marseille

Prospective & Coopération and various partner organizations have been following a cohort of approximately 1,200 homeless, migrants and other highly precarious people in the municipality of Marseille since 2020. Epicentre randomly selected the sites beforehand. The random selection of participants was then organized by the local partner Prospective & Coopération following the same methodology as for the strata in IDF.

#### Appendix 4. ODK Collect Form

| topic | variable name | label | values |
| --- | --- | --- | --- |
| Socio-demographic features | sexe | Gender | M / F |
| Socio-demographic features | age | Age (numeric) |  |
| Socio-demographic features | age_cat.new | Age categories (18/25/40/65) | 18-25; 25-40 ; 40-65 ;>=65 |
| Socio-demographic features | pays | Country |  |
| Socio-demographic features | region2.new | Region of Birth |  |
| Socio-demographic features | arrivee_france.new | Length of stay in France | Less than 10 years, before COVID-19 Since COVID-19 pandemic start More than 10 years (or born in France) Does not want to answer Does not know |
| Socio-demographic features | education.new | Level of education | Secondary  Never attended school / Illiterate Primary and/or literate Higher (university) |
| Socio-demographic features | langue | Language |  |
| Socio-demographic features | francophone.new | Francophone |  |
| Socio-demographic features | situation_administrative.new | Administrative status | Residence permit / Refugee status Asylum seeker / Ongoing application Undocumented French or European nationality (EU zone) Refusal to answer |
| Vaccination | vaccin_effectue | Covid-19 vaccine doses | 1/2/3/not yet but planned/no never/DNK |
| Vaccination | statut_vaccinal | Received at least one dose | Yes/No/DNK |
| Vaccination | statut_vaccinal2 | Full initial vaccination schedule (no booster) | Yes/No/DNK |
| Vaccination | statut_vaccinal3 | Full initial vaccination schedule (with booster) | Yes/No/DNK |
| Vaccination / Opinion on vaccination | vaccin_vie | Vaccinated in childhood (any antigen) | Yes/No/DNK |
| Vaccination / Opinion on vaccination | vaccin_opinion_perso_general.new | Opinion on vaccination (in general) |  |
| Vaccination / Opinion on vaccination | vaccin_opinion_perso_covid | Opinion on Covid-19 vaccination |  |
| Vaccination / Opinion on vaccination | vaccin_utile | Perceived utility of Covid-19 vaccination | Yes/No/DNK |
| Vaccination / Opinion on vaccination | vaccin_peur.new | Fear of Covid-19 vaccination | Yes/No/DNK |
| Vaccination / Opinion on vaccination | vaccin_opinion_entourage2.new | Family and friends' opinion on Covid-19 vaccination |  |
| Vaccination / Opinion on vaccination | pass_usage.new | Needs/uses vaccine certificate in daily life | Yes/No/DNK |
| Vaccination / Vaccinated (timing and tolerance) | vaccin_periode | Vaccination period |  |
| Vaccination / Vaccinated (timing and tolerance) | vaccin_certificat | Presentation of vaccination proof |  |
| Vaccination / Vaccinated (timing and tolerance) | vaccin_tolerance | Tolerance/Side Effects |  |
| Vaccination / Vaccinated (timing and tolerance) | vaccin_oui_proposition | Vaccination opportunity |  |
| Vaccination / Vaccinated (timing and tolerance) | vaccin_unique_injection.new_1 | One dose only - according to plan (Janssen) or 2d dose already scheduled | Yes/No/DNK |
| Vaccination / Vaccinated (timing and tolerance) | vaccin_unique_injection.new_3 | One dose only - Access or information issues (2d dose) | Yes/No/DNK |
| Vaccination / Vaccinated (timing and tolerance) | vaccin_unique_injection.new_9 | One dose only - refusal to get 2d dose | Yes/No/DNK |
| Vaccination / Vaccinated (timing and tolerance) | vaccin_unique_injection.new_97 | One dose only - other reason | Yes/No/DNK |
| Vaccination / Vaccinated (timing and tolerance) | vaccin_unique_injection.new_99 | One dose only - do not know/do not answer |  |
| Vaccination / Vaccinated (places) | vaccin_lieu.new | Place of vaccination |  |
| Vaccination / Vaccinated (places) | vaccin_proche.new | Proximity of the place of vaccination |  |
| Vaccination / Reasons for vaccination | vaccin_raison_positive.new_1 | Reason - positive opinion on vaccination overall | Yes/No/DNK |
| Vaccination / Reasons for vaccination | vaccin_raison_positive.new_3 | Reason - personal protection | Yes/No/DNK |
| Vaccination / Reasons for vaccination | vaccin_raison_positive.new_4 | Reason - protection of vulnerable relatives/friends | Yes/No/DNK |
| Vaccination / Reasons for vaccination | vaccin_raison_positive.new_5 | Reason - civic gesture (protection of everyone) | Yes/No/DNK |
| Vaccination / Reasons for vaccination | vaccin_raison_positive.new_6 | Reason - Advice from health professionnal / outreach vaccination activity | Yes/No/DNK |
| Vaccination / Reasons for vaccination | vaccin_raison_positive.new_7 | Reason - advice from social worker/religious group/community | Yes/No/DNK |
| Vaccination / Reasons for vaccination | vaccin_raison_positive.new_11 | Reason - sense of compelment/constraint (to get vaccine certificate, asked by employer, to be able to travel…) | Yes/No/DNK |
| Vaccination / Reasons for vaccination | vaccin_raison_positive.new_97 | Reason - other | Yes/No/DNK |
| Vaccination / Reasons for vaccination | vaccin_raison_positive.new_99 | Reason - DNK/DNA |  |
| Vaccination / Reasons for vaccination | pass_absence.new | Vaccine certificate as the main driver | Yes/No/DNK |
| Vaccination / Vaccinated (additional) | vaccin_raison_positive_2_3_4.new | One dose only - Protection in general (self, relatives, everyone) | Yes/No/DNK |
| Vaccination / Vaccinated (additional) | vaccin_raison_positive_3_4.new | Reason - Protection of relatives/friends | Yes/No/DNK |
| Vaccination / Vaccinated (additional) | vaccin_mois | Month of vaccination |  |
| Vaccination / Not vaccinated | vaccin_intention | Intention to get vaccinated |  |
| Vaccination / Not vaccinated | vaccin_non_proposition | Vaccination opportunity |  |
| Vaccination / Not vaccinated | vaccin_raison_negative_1 | Reason - I was not able to | Yes/No/DNK |
| Vaccination / Not vaccinated | vaccin_raison_negative_2 | Reason - I did not want to | Yes/No/DNK |
| Vaccination / Not vaccinated | vaccin_raison_negative_99 | Reason - DNK/DNA |  |
| Vaccination / Not vaccinated (access issues) | vaccin_raison_negative_accessibilite.new2_1 | Reason -(Wrong) beliefs/info on accessibility (I thought medical coverage, money was required) | Yes/No/DNK |
| Vaccination / Not vaccinated (access issues) | vaccin_raison_negative_accessibilite.new2_3 | Reason - Lack of information (I did not know how or when) or opportunity (I did not know where) | Yes/No/DNK |
| Vaccination / Not vaccinated (access issues) | vaccin_raison_negative_accessibilite.new2_4 | Reason - Physical/Practical barriers (center was too far, to hard to reach, too crowded) | Yes/No/DNK |
| Vaccination / Not vaccinated (access issues) | vaccin_raison_negative_accessibilite.new2_9 | Reason - My peers advised me not to get vaccinated | Yes/No/DNK |
| Vaccination / Not vaccinated (access issues) | vaccin_raison_negative_accessibilite.new2_12 | Reason - I had a medical contra-indication (pregnant, even if wrong, allergy…) | Yes/No/DNK |
| Vaccination / Not vaccinated (access issues) | vaccin_raison_negative_accessibilite.new2_97 | Reason - other | Yes/No/DNK |
| Vaccination / Not vaccinated (access issues) | vaccin_raison_negative_accessibilite.new2_99 | Reason - DNK/DNA |  |
| Vaccination / Not vaccinated (access issues) | vaccin_solution_accessibilite_1 | Solution to improve access - make the vaccine fully free and unconditional | Yes/No/DNK |
| Vaccination / Not vaccinated (access issues) | vaccin_solution_accessibilite_2 | Solution to improve access - a close drugstore offers vaccination | Yes/No/DNK |
| Vaccination / Not vaccinated (access issues) | vaccin_solution_accessibilite_3 | Solution to improve access- a close GP offers vaccination | Yes/No/DNK |
| Vaccination / Not vaccinated (access issues) | vaccin_solution_accessibilite_4 | Solution to improve access- an outreach team offers vaccination on my place of living | Yes/No/DNK |
| Vaccination / Not vaccinated (access issues) | vaccin_solution_accessibilite_5 | Solution to improve access- an outreach team comes to me (for roofless/living in the streets) | Yes/No/DNK |
| Vaccination / Not vaccinated (access issues) | vaccin_solution_accessibilite_6 | Solution to improve access- a vaccination center closer to me | Yes/No/DNK |
| Vaccination / Not vaccinated (access issues) | vaccin_solution_accessibilite_7 | Solution to improve access- a mobile clinic closer to me | Yes/No/DNK |
| Vaccination / Not vaccinated (access issues) | vaccin_solution_accessibilite_8 | Solution to improve access- vaccination fully anonymous/no ID required | Yes/No/DNK |
| Vaccination / Not vaccinated (access issues) | vaccin_solution_accessibilite_9 | Solution to improve access- interpretation/translation in my language available | Yes/No/DNK |
| Vaccination / Not vaccinated (access issues) | vaccin_solution_accessibilite_10 | Solution to improve access- no appointment/booking needed | Yes/No/DNK |
| Vaccination / Not vaccinated (access issues) | vaccin_solution_accessibilite_11 | Solution to improve access- support in booking/taking appointment for vaccination | Yes/No/DNK |
| Vaccination / Not vaccinated (access issues) | vaccin_solution_accessibilite_97 | Solution to improve access- other | Yes/No/DNK |
| Vaccination / Not vaccinated (access issues) | vaccin_solution_accessibilite_99 | Solution to improve access- DNK/DNA |  |
| Vaccination / Not vaccinated (acceptability issues) | vaccin_raison_negative_acceptabilite.new_1 | Reason - Denial of the crisis/Conspiracy theories | Yes/No/DNK |
| Vaccination / Not vaccinated (acceptability issues) | vaccin_raison_negative_acceptabilite.new_3 | Reason - Vaccination not a priority/COVID-19 not a threat (complacency) | Yes/No/DNK |
| Vaccination / Not vaccinated (acceptability issues) | vaccin_raison_negative_acceptabilite.new_4 | Reason - Vaccine is useless/inefficient | Yes/No/DNK |
| Vaccination / Not vaccinated (acceptability issues) | vaccin_raison_negative_acceptabilite.new_10 | Reason -Fear of immediate side effects | Yes/No/DNK |
| Vaccination / Not vaccinated (acceptability issues) | vaccin_raison_negative_acceptabilite.new_11 | Reason - Fear of a serious disease because of vaccination | Yes/No/DNK |
| Vaccination / Not vaccinated (acceptability issues) | vaccin_raison_negative_acceptabilite.new_14 | Reason - Lack of advice /Not recommended by health professional | Yes/No/DNK |
| Vaccination / Not vaccinated (acceptability issues) | vaccin_raison_negative_acceptabilite.new_15 | Reason -Discouraged by relatives/friends/community | Yes/No/DNK |
| Vaccination / Not vaccinated (acceptability issues) | vaccin_raison_negative_acceptabilite.new_97 | Reason - other |  |
| Vaccination / Not vaccinated (acceptability issues) | vaccin_raison_negative_acceptabilite.new_99 | Reason - DNK/DNA |  |
| Vaccination / Not vaccinated (acceptability issues) | vaccin_solution_acceptabilite_1 | Solution to improve acceptability - we need a better vaccine without side effects | Yes/No/DNK |
| Vaccination / Not vaccinated (acceptability issues) | vaccin_solution_acceptabilite_2 | Solution to improve acceptability - I need information in my language | Yes/No/DNK |
| Vaccination / Not vaccinated (acceptability issues) | vaccin_solution_acceptabilite_3 | Solution to improve acceptability - I'd rather information were stable and less confusing | Yes/No/DNK |
| Vaccination / Not vaccinated (acceptability issues) | vaccin_solution_acceptabilite_4 | Solution to improve acceptability -I want advice from my GP | Yes/No/DNK |
| Vaccination / Not vaccinated (acceptability issues) | vaccin_solution_acceptabilite_5 | Solution to improve acceptability -I want advice by social workers/person of trust | Yes/No/DNK |
| Vaccination / Not vaccinated (acceptability issues) | vaccin_solution_acceptabilite_6 | Solution to improve acceptability - I want advice from my relatives/community | Yes/No/DNK |
| Vaccination / Not vaccinated (acceptability issues) | vaccin_solution_acceptabilite_7 | Solution to improve acceptability -vaccination needs to be mandatory | Yes/No/DNK |
| Vaccination / Not vaccinated (acceptability issues) | vaccin_solution_acceptabilite_8 | Solution to improve acceptability - financial compensation | Yes/No/DNK |
| Vaccination / Not vaccinated (acceptability issues) | vaccin_solution_acceptabilite_97 | Solution to improve acceptability - other | Yes/No/DNK |
| Vaccination / Not vaccinated (acceptability issues) | vaccin_solution_acceptabilite_98 | Solution to improve acceptability - no solution will work | Yes/No/DNK |
| Vaccination / Not vaccinated (acceptability issues) | vaccin_solution_acceptabilite_99 | Solution to improve acceptability - DNK/DNA |  |
| Place of living | strate.new | Survey stratum |  |
| Place of living | ethos_op.new | Ethos Typology (Operational categories) |  |
| Place of living | ethos_concept.new | Ethos Typology (Concept) |  |
| Place of living | ethos_light.new | Ethos Typology (Light) |  |
| Place of living | logement_mois.new | Main place of living over the past 3 months |  |
| Place of living (additional) | logement_hier.new | Place of stay (last night) |  |
| Place of living (additional) | strate_mois2.new | Stratum based on Main living place the last 3 months |  |
| Place of living (additional) | dep | Department |  |
| Place of living (mobility) | logement_changement | Change in housing/location over the past three months | Yes/No/DNK |
| Financial / food situation / Income | revenu2.new | Source of Income | No income/informal activities/no regular pension or aid Declared and official sources of income (salary, pensions, state aids…) DNK/DNA |
| Financial / food situation / Income | situation_financiere.new | Financial situation | DNK/DNA Acceptable/Good financial situation Very tough financial situation Borderline financial situation (can change day to day) |
| Financial / food situation / Income | situation_alimentaire3.new | Food security | Not enough food and not good enough Food enough in quantity and quality |
| Financial / food situation / Income | source_alimentaire_1 | Buys his/her own meals (most of the time) | Yes/No/DNK |
| Financial / food situation / Income | source_alimentaire_2 | Meals provided by relatives/family/friends | Yes/No/DNK |
| Financial / food situation / Income | source_alimentaire_3 | Meals through food distributions | Yes/No/DNK |
| Financial / food situation / Income | source_alimentaire_4 | Meals through panhandling | Yes/No/DNK |
| Financial / food situation / Income | source_alimentaire_5 | Meals through dumpster rummaging | Yes/No/DNK |
| Financial / food situation / Income | source_alimentaire_6 | Meals difficult to get/not enough in quantity | Yes/No/DNK |
| Financial / food situation / Income | source_alimentaire_7 | Meals provided by site managers | Yes/No/DNK |
| Financial / food situation / Income | source_alimentaire.new | Source of food/meals |  |
| Financial / food situation / Income | revenu_1 | Source of income - declared employment | Yes/No/DNK |
| Financial / food situation / Income | revenu_2 | Source of income - undeclared employment | Yes/No/DNK |
| Financial / food situation / Income | revenu_3 | Source of income - retirement pension and/or alimony | Yes/No/DNK |
| Financial / food situation / Income | revenu_4 | Source of income - unemployment benefit | Yes/No/DNK |
| Financial / food situation / Income | revenu_5 | Source of income - scholarship | Yes/No/DNK |
| Financial / food situation / Income | revenu_6 | Source of income - social solidarity benefit | Yes/No/DNK |
| Financial / food situation / Income | revenu_7 | Source of income - other(s) benefit(s) | Yes/No/DNK |
| Financial / food situation / Income | revenu_8 | Source of income - begging | Yes/No/DNK |
| Financial / food situation / Income | revenu_9 | Source of income - associative/housing support | Yes/No/DNK |
| Financial / food situation / Income | revenu_10 | Source of income - family/friend support | Yes/No/DNK |
| Financial / food situation / Income | revenu_11 | Source of income - none | Yes/No/DNK |
| Financial / food situation / Income | revenu_99 | Source of income - unknown | Yes/No/DNK |
| Place of living | logement_entourage.new | Household structure |  |
| Support and coping systems | soutien_dispositif.new_1 | Attends food distributions on a regular basis | Yes/No/DNK |
| Support and coping systems | soutien_dispositif.new_2 | Benefits from outreach activities | Yes/No/DNK |
| Support and coping systems | soutien_dispositif.new_6 | Supported by site managers/social workers on site | Yes/No/DNK |
| Support and coping systems | soutien_dispositif.new_7 | Supported by social workers (external) | Yes/No/DNK |
| Support and coping systems | soutien_dispositif.new_10 | Supported by state/city services | Yes/No/DNK |
| Support and coping systems | soutien_dispositif.new_11 | Supported by health professionals | Yes/No/DNK |
| Support and coping systems | soutien_dispositif.new_15 | Supported by religieux groups/community | Yes/No/DNK |
| Support and coping systems | soutien_dispositif.new_17 | Autonomous / Does not receive or need support whatsoever | Yes/No/DNK |
| Support and coping systems | soutien_dispositif.new2 | Benefits from at least one support system | Yes/No/DNK |
| Support and coping systems | soutien_accompagnement_medical.new | Benefits from support to attend and navigate the healthcare system |  |
| Moral and social support | ecoute_proche.new | Support from relatives (can speak to/be listened by relatives) |  |
| Moral and social support | materiel_proche.new | Material support from relatives (money, food, clothes etc) |  |
| Moral and social support | conseil_proche.new | Advices from relatives (to find a job etc) |  |
| Moral and social support | confiance_proche.new | Moral support by relatives (motivation) |  |
| Moral and social support | ecoute_asso.new | Support from organization members (can speak to/be listened by members) |  |
| Moral and social support | materiel_asso.new | Material support from org. members (money, food, clothes etc) |  |
| Moral and social support | conseil_asso.new | Advices from org. members (to find a job etc) |  |
| Moral and social support | confiance_asso.new | Moral support by org. members (motivation) |  |
| Moral and social support | soutien_asso2.new | Global support by organizations |  |
| Moral and social support | solitude.new | Self-reported feeling of loneliness | I feel taken care of/supported/not specially lonely I feel very taken care of/supported. I don't feel lonely at all I don't feel taken care of/supported. I feel lonely sometimes. I feel very lonely DNK/DNA |
| COVID-19 (vaccination) Information Sources | connexion.new | Internet connection | Other: another mean of connection  No, never uses internet Yes, seldom or often uses internet Does not know |
| COVID-19 (vaccination) Information Sources | information_masque.new | Compliance to protection measures (wear a facemask in public transportation etc) | Yes, I wear it and I think it's useful It depends on the context No, this is useless DNK/DNA |
| COVID-19 (vaccination) Information Sources | information_covid_1 | COVID-19 (Vaccine) information source - internet | Yes/No/DNK |
| COVID-19 (vaccination) Information Sources | information_covid_2 | COVID-19 (Vaccine) information source - social media | Yes/No/DNK |
| COVID-19 (vaccination) Information Sources | information_covid_1_2.new | COVID-19 (Vaccine) information source - internet / social media | Yes/No/DNK |
| COVID-19 (vaccination) Information Sources | information_covid_3 | COVID-19 (Vaccine) information source - television/radio | Yes/No/DNK |
| COVID-19 (vaccination) Information Sources | information_covid_4 | COVID-19 (Vaccine) information source - print media/classic media | Yes/No/DNK |
| COVID-19 (vaccination) Information Sources | information_covid_5 | COVID-19 (Vaccine) information source - posters/leaflets | Yes/No/DNK |
| COVID-19 (vaccination) Information Sources | information_covid_6 | COVID-19 (Vaccine) information source - social workers | Yes/No/DNK |
| COVID-19 (vaccination) Information Sources | information_covid_7 | COVID-19 (Vaccine) information source - site manager | Yes/No/DNK |
| COVID-19 (vaccination) Information Sources | information_covid_8 | COVID-19 (Vaccine) information source - health professionnals | Yes/No/DNK |
| COVID-19 (vaccination) Information Sources | information_covid_6_8.new | COVID-19 (Vaccine) information source - health or social workers/professionnals | Yes/No/DNK |
| COVID-19 (vaccination) Information Sources | information_covid_6_7_8.new | COVID-19 (Vaccine) information source - trusted third-party | Yes/No/DNK |
| COVID-19 (vaccination) Information Sources | information_covid_9 | COVID-19 (Vaccine) information source - close relatives | Yes/No/DNK |
| COVID-19 (vaccination) Information Sources | information_covid_10 | COVID-19 (Vaccine) information source - community | Yes/No/DNK |
| COVID-19 (vaccination) Information Sources | information_covid_11 | COVID-19 (Vaccine) information source - religious groups | Yes/No/DNK |
| COVID-19 (vaccination) Information Sources | information_covid_9_10_11.new | COVID-19 (Vaccine) information source - relatives/family/community | Yes/No/DNK |
| COVID-19 (vaccination) Information Sources | information_covid_12 | COVID-19 (Vaccine) information source - none | Yes/No/DNK |
| COVID-19 (vaccination) Information Sources | information_covid_suffisante.new | Satisfaction with COVID-19 information (overall) | Yes/No/DNK |
| COVID-19 (vaccination) Information Sources | information_covid_suffisante_1 | Yes, I'm satisfied and feel I am informed enough | Yes/No/DNK |
| COVID-19 (vaccination) Information Sources | information_covid_suffisante_2 | No, I didn't get enough information | Yes/No/DNK |
| COVID-19 (vaccination) Information Sources | information_covid_suffisante_3 | No, it was too confusing and not clear enough | Yes/No/DNK |
| COVID-19 (vaccination) Information Sources | information_covid_suffisante_4 | No, it was too much information, I feel saturated | Yes/No/DNK |
| COVID-19 (vaccination) Information Sources | information_covid_suffisante_5 | No, it was changing all the time, too contradictory | Yes/No/DNK |
| COVID-19 (vaccination) Information Sources | confiance_autorite.new | Trust in the crisis management by the authorities | None/Low/Medium/High/Total |
| COVID-19 (vaccination) Information Sources | confiance_autorite_covid | Trust in the crisis management by the authorities | 1 to 10 |
| Health-related information | couverture_maladie | Has medical coverage |  |
| Health-related information | couverture_maladie.new | Has medical coverage |  |
| Health-related information | etat_sante | Self reported health status (num.) |  |
| Health-related information | etat_sante.new | Self-reported health status |  |
| Health-related information | maladie_chronique_perso.new | Chronic condition (self-reported) |  |
| Health-related information | medecin_traitant | Is followed by a regular GP |  |
| Health-related information | derniere_consultation.new | Date of last consultation (hospital/GP) |  |
| Health-related information | covid_perso.new | Previous history of COVID-19 |  |
| Health-related information | covid_perso_hospitalise | Previous hospitalization for COVID-19 |  |
| Health-related information | covid_indiv.new | Previous history/hospitalization of COVID-19 |  |
| Health-related information | covid_entourage | Previous history of COVID-19 among family or friends |  |
| Health-related information | covid_entourage_hospitalise | Previous hospitalization for COVID-19 among family or friends |  |
| Health-related information | covid_ent.new | Previous history/hospitalization for COVID-19 among family or friends |  |
| Health-related information | maladie_chronique_entourage.new | Vulnerable/at risk for COVID-19 people among relatives |  |
| Health literacy | sante_litteratie_ecrite_affiche | Can read and understand written medical instructions |  |
| Health literacy | sante_litteratie_ecrite_medicament | Can read and understand drug notices |  |
| Health literacy | sante_litteratie_orale_medicament | Can understand oral medical instructions |  |
| Health literacy | sante_litteratie_orale_rdv | Can understand oral medical advice |  |
| Health literacy | sante_litteratie_formulaire | Can correctly fill out a medical form |  |
| Health literacy | sante_litteratie_ecrite_medicament.new.imput | Can read and understand drug notices |  |
| Health literacy | sante_litteratie_orale_medicament.new.imput | Can understand oral medical instructions |  |
| Health literacy | sante_litteratie_orale_rdv.new.imput | Can understand oral medical advice |  |
| Health literacy | sante_litteratie_ecrite_affiche.new.imput | Can read and understand written medical instructions |  |
| Health literacy | sante_litteratie_formulaire.new.imput | Can correctly fill out a medical form |  |
| Health literacy | litteratie_ecrit.new | Health Literacy (Written) |  |
| Health literacy | litteratie_oral.new | Health Literacy (Oral) |  |
| Health literacy | sante_litteratie_ecrite_affiche.new | Can read and understand written medical instructions |  |
| Health literacy | sante_litteratie_ecrite_medicament.new | Can read and understand drug notices |  |
| Health literacy | sante_litteratie_orale_medicament.new | Can understand oral medical instructions |  |
| Health literacy | sante_litteratie_orale_rdv.new | Can understand oral medical advice |  |
| Health literacy | sante_litteratie_formulaire.new | Can correctly fill out a medical form |  |
| Discrimination | discrimination_mefiance.new | Experienced discrimination since Covid-19 outbreak |  |
| Discrimination | discrimination_expulsion | Is more afraid of being expelled from France since COVID-19 crisis |  |
| Discrimination | discrimination_vaccin | Has been denied vaccination |  |
| Discrimination | discrimination_vaccin.imput | Has been denied vaccination |  |
| Discrimination | discrimination_soin | has been denied healthcare |  |
| Discrimination (additional) | discrimination_mefiance | Experienced discrimination since Covid-19 outbreak |  |
| Recruitment site information | pass_site | Vaccine certificate is mandatory to access site | Yes/No/DNK |
| Recruitment site information | support_social | Presence of social workers on site |  |
| Recruitment site information | support_social.new | Presence of social workers on site | Yes/No/DNK |
| Recruitment site information | support_medical | Presence of GP on site |  |
| Recruitment site information | support_medical.new | Presence of GP on site | Yes/No/DNK |
| Recruitment site information | support_sanitaire | Presence of health professionnals on site |  |
| Recruitment site information | support_sanitaire.new | Presence of health professionnals on site | Yes/No/DNK |
| Recruitment site information | dist_medecin.new | Proximity of a GP (>15 or <15 min by foot) | Yes/No/DNK |
| Recruitment site information | dist_pharmacie.new | Proximity of a drugstore (>15 or <15 min by foot) | Yes/No/DNK |
| Recruitment site information | dist_hopital.new | Proximity of a hospital (>15 or <15 min by foot) | Yes/No/DNK |
| Recruitment site information | dist_clinmob.new | Proximity of a mobile clinic (targeting PEH) | Yes/No/DNK |
| Recruitment site information | dist_centrecovid.new | Proximity of a vaccination center (>15 or <15 min by foot) | Yes/No/DNK |
| Recruitment site information | dist_specifprec.new | Proximity of vaccination site targeting PEH (>15 or <15 min by foot) | Yes/No/DNK |
| Recruitment site information | dist_siteprec.new_TR4 | Proximity of vaccination site targeting PEH (>15 or <15 min by foot) | Yes/No/DNK |
| Recruitment site information | dist_centrevac.new_TR4 | Proximity of a vaccination center (>15 or <15 min by foot) | Yes/No/DNK |
| Recruitment site information | dist_gene.new_TR4 | Proximity of a GP (>15 or <15 min by foot) | Yes/No/DNK |
| Recruitment site information | dist_pharm.new_TR4 | Proximity of a drugstore (>15 or <15 min by foot) | Yes/No/DNK |
| Recruitment site information | dist_clinique.new_TR4 | Proximity of a mobile clinic (targeting PEH) | Yes/No/DNK |
| Recruitment site information | messindiv_acteurs.new | Personalized sensitization/mobilization on site | Yes/No/DNK |
| Recruitment site information | reu_acteurs.new | Information/Mobilization meetings on site | Yes/No/DNK |
| Recruitment site information | sensib_site.new | Sensitization/Mobilization on site (any type) | Yes/No/DNK |
| Recruitment site information | dispositif_vaccin_1 | Vaccination activity on site | Yes/No/DNK |
| Recruitment site information | dispositif_vaccin_2 | Partnership with organization to address people for vaccination | Yes/No/DNK |
| Recruitment site information | dispositif_vaccin_3 | Support to book/take appointment for vaccination | Yes/No/DNK |
| Recruitment site information | dispositif_vaccin_4 | Personal accompaniment to vaccination place | Yes/No/DNK |
| Recruitment site information | dispositif_vaccin_5 | Practival information | Yes/No/DNK |
| Recruitment site information | dispositif_vaccin_6 | Indication to closest drugstore/pharmacist | Yes/No/DNK |
| Recruitment site information | dispositif_vaccin_7 | Indication to closest general practitioner | Yes/No/DNK |
| Recruitment site information | dispositif_vaccin_98 | No specific activity | Yes/No/DNK |
| Recruitment site information | action_site.new | on site Interventions around COVID-19 vaccination | Yes/No/DNK |
| Recruitment site information | action_site_sensibilisation.new | Intervention on site - Awareness/sensitization | Yes/No/DNK |
| Recruitment site information | action_site_accompagnement.new | Intervention on site - Personalized support/mobilization | Yes/No/DNK |
| Recruitment site information | action_site_vaccination.new | Intervention on site - Actual vaccination activity | Yes/No/DNK |
| Recruitment site information | site | Site |  |

#### Appendix 5. Statistical Analysis Plan

Direct Standardization to compare vaccine uptake

First dose intake and vaccine coverage in our study population were compared to French general population. A weighted direct standardization by age categories has been performed (age cut-offs : 18, 25, 40, 55, 65, >65) and 95% CI were computed for overall study population and for each strata (according to Naing *Easy way to learn standardization : direct and indirect methods* <https://www.ncbi.nlm.nih.gov/pmc/articles/PMC3406211/pdf/mjms-7-1-010.pdf> ).

Reference data were downloaded from the Assurance Maladie website (link below). These data include vaccination and population indicators (Insee) for the French metropolitan adult population, over the entire period). Source: [*https://datavaccin-covid.ameli.fr/explore/dataset/donnees-vaccination-par-tranche-dage-type-de-vaccin-et-departement/table/?sort=departement_residence&q=date%3D2022-01-09+and+not+(departement_residence:%27999%27+OR+departement_residence:%27Tout+d%C3%A9partement%27)&refine.type_vaccin=Tout+vaccin&refine.classe_age=TOUT_AGE*](https://datavaccin-covid.ameli.fr/explore/dataset/donnees-vaccination-par-tranche-dage-type-de-vaccin-et-departement/table/?sort=departement_residence&q=date%3D2022-01-09+and+not+(departement_residence:%27999%27+OR+departement_residence:%27Tout+d%C3%A9partement%27)&refine.type_vaccin=Tout+vaccin&refine.classe_age=TOUT_AGE)

Population breakdown by Age, for the Ile-de-France region and for the study population (All stratum and by stratum), are as follows:

| **Age** | **France (million)** | **France (%)** | **All stratum (%) (n=100567)** | **Accomodation (%) (N=73159)** | **Homeless (Streets) (%) (N=4620)** | **Housing (%) (N=22788)** |
| --- | --- | --- | --- | --- | --- | --- |
| 18·25) | 5·3 | 10·3 | 8·3 | 9·3 | 14·6 | 3·9 |
| 25·40) | 11·7 | 22·8 | 47·0 | 51·8 | 43·8 | 32·1 |
| 40·55) | 12·6 | 24·6 | 25·6 | 25·6 | 28·4 | 25·3 |
| 55·65) | 8·2 | 16·0 | 10·6 | 8·9 | 9·8 | 16·1 |
| 65·Inf) | 13·4 | 26·2 | 8·4 | 4·4 | 3·4 | 22·5 |
| Total | 51·2 |  |  |  |  |  |

Direct standardization was done using the “DSR” package in R : <https://rdrr.io/cran/dsr/>

*Missing values*

For variables with a high number of missing values (>5%), missing values mechanism assumed to be MAR mechanism (missing at random) and verified. For each variable, several imputations methods were compared (multiple regression, random forest) and the one which gave the lowest error rate was retained. Only imputed variables with an error rate lower than 20% were used for the multivariable model. Imputation was done with the MICE package in R : <https://cran.r-project.org/web/packages/mice/mice.pdf>

Univariate

Univariate logistic regression analysis explored vaccine uptake associated factors for all strata combined. A multilevel multivariable logistic regression model was constructed with random intercepts for specific recruitment sites to account for clustering and random effects on several variables after testing for validity (see below). We included variables that could explain differences in vaccine uptake proportions.

Multivariate

Only variables with p≤0·2 after univariate were retained in the full multivariate model. Multicollinearity was verified prior to model selection and variables with a Variance Inflation Factor (VIF, for continuous variables) > 5 or a generalized VIF (GVIF, for discrete variables) >2·5 were dropped (see Midi et al. *Collinearity diagnostics of binary logistic regression model* <https://doi.org/10.1080/09720502.2010.10700699> ). Random effects were tested on the full model and selected in order to minimize the second-order Akaike Information Criteria (AICc). After random effects selection, fixed effects were selected with a backwards procedure, minimizing AIC.

We also 'forced' a few factors that were not significant in univariate but were relevant for model adjustment (potential confounding factors or factors known to be linked with vaccine uptake in the literature). Moreover, some factors appeared too important/interesting to miss (opinions about vaccination, in particular).

Validation of the final model consisted in standardizing residuals analysis (overdispersion, distribution, outliers) and computing coefficients of determination (according to Menard et al. *Coefficients of determination for multiple logistic regression analysis* <https://doi.org/10.2307/2685605> ).

Stratified analyses

Descriptive, univariate and multilevel multivariable analyses were performed for each individual stratum, following global sample procedure.

Stratum final models may differ from the final global model, since the individuals characteristics can vary significantly.

Site-level analyses

We performed an unweighted negative binomial regression on the total number of vaccinated individuals by site as a count outcome and site-related variables as covariables following aforementioned procedure. Questions were usually asked to site managers for Housed and Acommodated participants moslty. Streets individuals were then excluded de facto from the analyses.

Variables retained for descriptive, univariate and multivariable analyses were:

Stratum

Presence of social workers on site

Presence of health professionals on site

Awareness Raising/sensitization activities

Personalized support/mobilization activities

Actual vaccination activities on site

Distance to closest vaccination site targeting PEH

Distance to closest pharmacist/drugstore

Distance to closest vaccination center

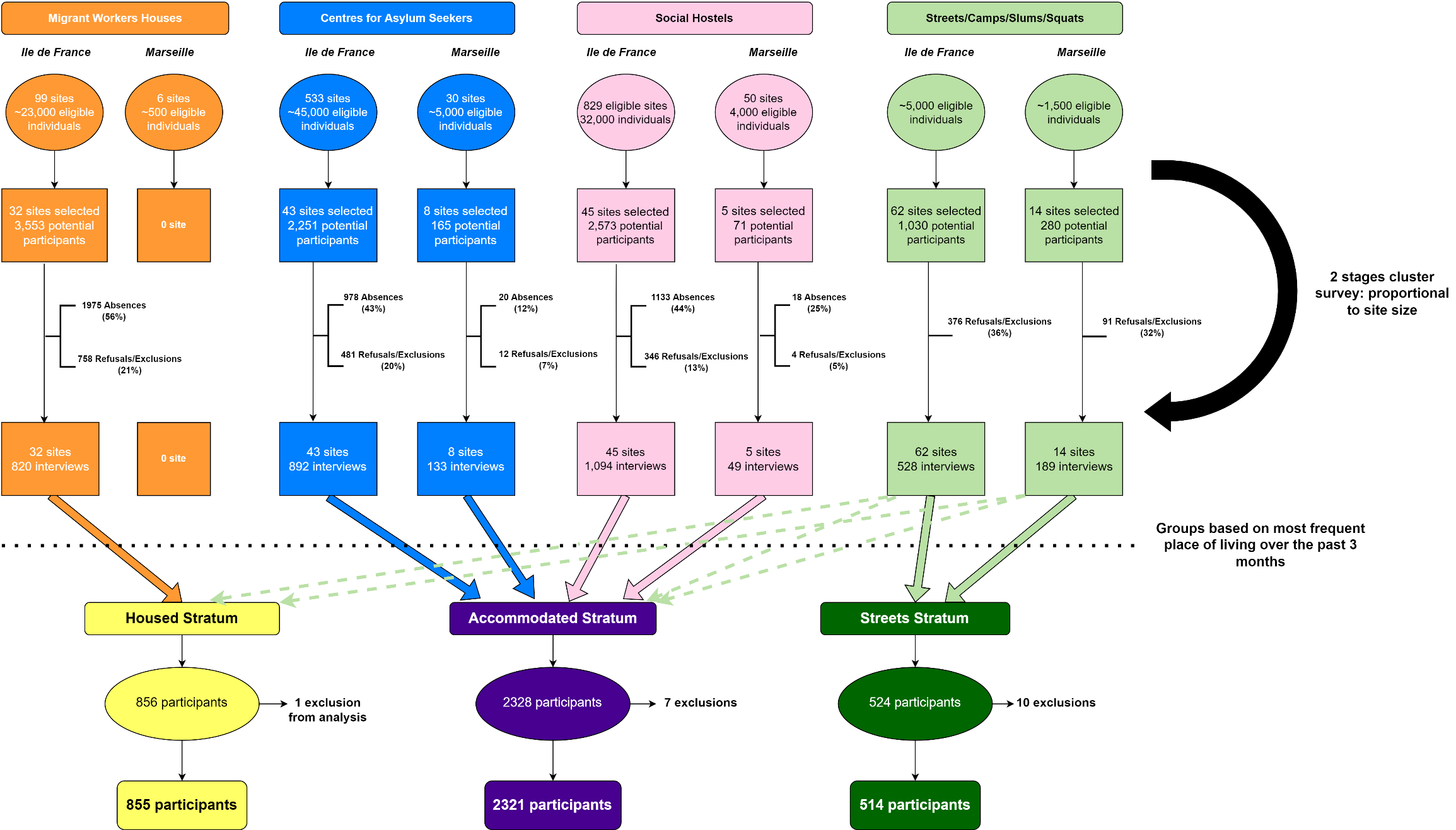

#### Appendix 6. Figure S1. Study flow chart

#### Appendix 7. Maps.

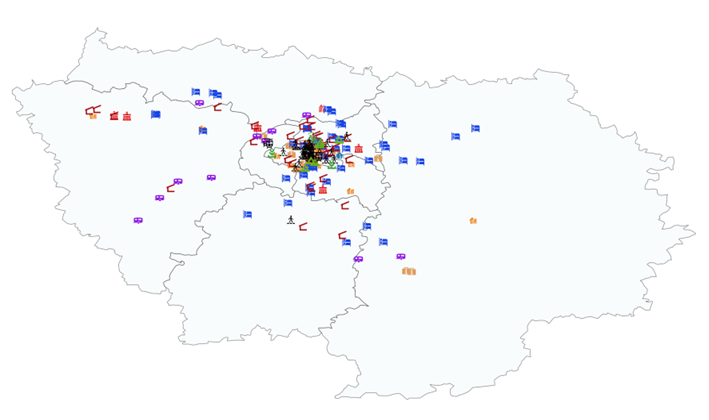
Recruitment Sites in Ile de France Region

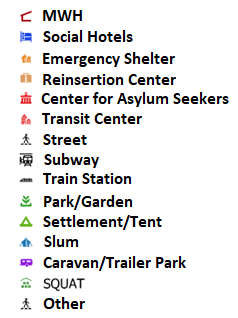

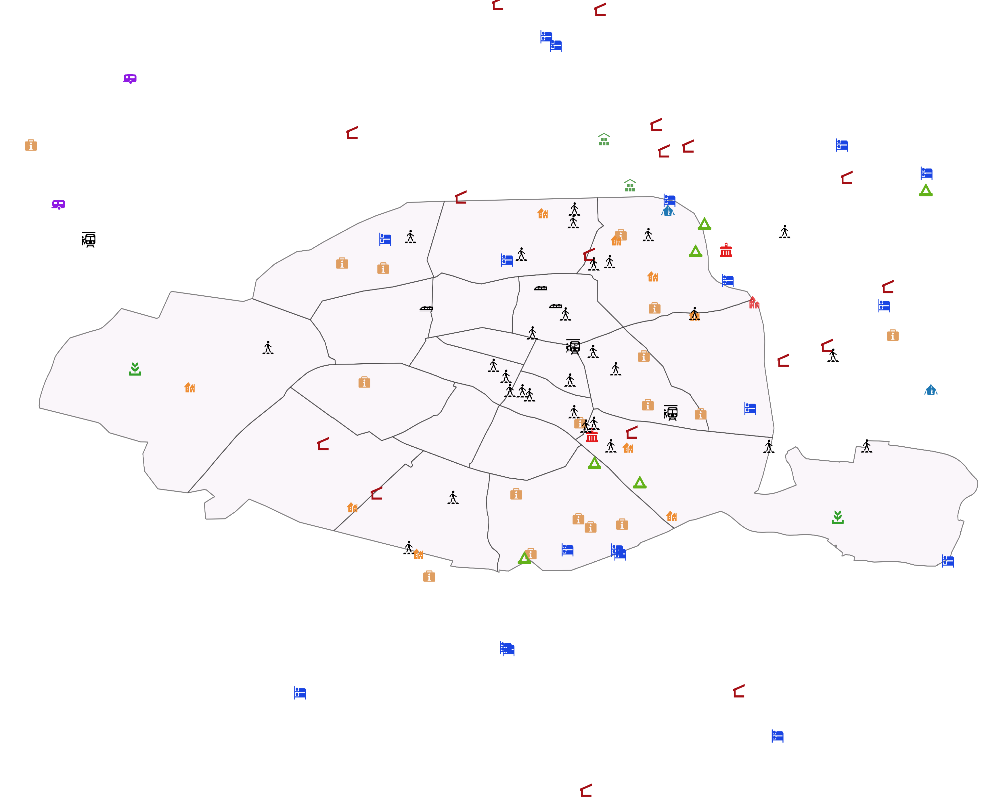
Paris and suburbs

Marseille Metropilitan Area

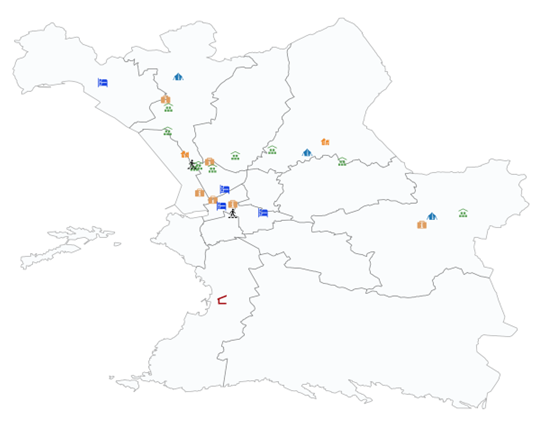

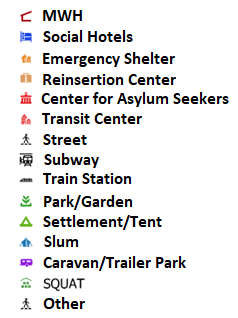

#### Appendix 8. Table S1. Descriptive Analysis (Global and by stratum) – Remaining Variables

|  | | **All stratum** | | | **Housing** | | | **Accomodation** | | | **Homeless (Streets)** | | |  |
| --- | --- | --- | --- | --- | --- | --- | --- | --- | --- | --- | --- | --- | --- | --- |
| **Variable** | **Categories** | **N** | **%** | **IC95 %** | **N** | **%** | **IC95 %** | **N** | **%** | **IC95 %** | **N** | **%** | **IC95 %** | **p-value** |
| **Place of living** | | | | | | | | | | | | | | |
| **Ethos Typology (Concept)** | Rents a flat / hosted by family | 41 | 1·1 | 0·1-2·1 | 38 | 4·6 | 0·4-8·8 |  |  |  | 3 | 1·7 | -0·9-4·3 | p<0·001*** |
|  | Rooflessness (without a shelter of any kind, sleeping rough) | 108 | 3·0 | 2·3-3·6 |  |  |  | 3 | 0·1 | 0-0·3 | 105 | 62·6 | 53·5-71·7 |  |
|  | Houselessness (with a place to sleep but temporary/emergency in institutions or shelter ) | 3,396 | 93·1 | 91·7-94·4 | 796 | 95·4 | 91·2-99·6 | 2,6 | 98·2 | 97·6-98·8 |  |  |  |  |
|  | Living in insecure housing (can be expelled easily, poor living conditions etc) | 72 | 2·0 | 1·4-2·5 |  |  |  | 45 | 1·7 | 1·1-2·3 | 27 | 16·2 | 9-23·4 |  |
|  | Living in inadequate housing (in caravans on illegal campsites, in unfit housing, in extreme overcrowding) | 33 | 0·9 | 0·6-1·2 |  |  |  |  |  |  | 33 | 19·5 | 12·8-26·2 |  |
|  | *Missing* | *42* | *1·1* |  | *4* | *0·4* |  | *35* | *1·3* |  | *3* | *1·7* |  |  |
| **Ethos Typology (Op· Cat·)** | Rents his/her own flat | 41 | 1·1 | 0·1-2·1 | 38 | 4·6 | 0·4-8·8 |  |  |  | 3 | 1·7 | -0·9-4·3 | p<0·001*** |
|  | Rooflessness (Living in the streets or public spaces without a shelter that can be defined as living quarters) | 120 | 3·3 | 2·7-3·9 |  |  |  |  |  |  | 120 | 71·9 | 63·3-80·4 |  |
|  | Emergency accommodation (Overnight shelters, People with no place of usual residence who move frequently between various types of accommodation) | 935 | 25·6 | 20·3-30·9 |  |  |  | 935 | 35·3 | 28·3-42·3 |  |  |  |  |
|  | People living in accommodation for the homeless /migrants (homeless hostels, centers for asylum seekers) | 2,464 | 67·5 | 62·3-72·8 | 796 | 95·4 | 91·2-99·6 | 1,668 | 63·0 | 56·1-69·9 |  |  |  |  |
|  | Inadequate Housing ( nonconventional dwellings, unfit housing, in extreme overcrowding) | 17 | 0·5 | 0·2-0·7 |  |  |  |  |  |  | 17 | 10·2 | 4·9-15·5 |  |
|  | Hosted by a third-part/Squat | 72 | 2·0 | 1·4-2·5 |  |  |  | 45 | 1·7 | 1·1-2·3 | 27 | 16·2 | 9-23·4 |  |
|  | *Missing* | *42* | *1·1* |  | *4* | *0·4* |  | *35* | *1·3* |  | *3* | *1·7* |  |  |
| **Financial / food situation / Income** | | | | | | | | | | | | | | |
| **Food security** | Food enough in quantity and quality | 1,879 | 51·5 | 48·1-54·8 | 587 | 70·5 | 63·7-77·3 | 1,237 | 46·6 | 42·6-50·6 | 56 | 33·7 | 25·8-41·7 | p<0·001*** |
|  | Not enough food and not good enough | 1,773 | 48·5 | 45·2-51·9 | 246 | 29·5 | 22·7-36·3 | 1,418 | 53·4 | 49·4-57·4 | 109 | 66·3 | 58·3-74·2 |  |
|  | *Missing* | *30* | *0·8* |  | *3* | *0·4* |  | *22* | *0·8* |  | *5* | *3·0* |  |  |
| **Source of income - declared employment** | No | 2,916 | 79·2 | 76·9-81·5 | 558 | 66·8 | 62·4-71·2 | 2,198 | 82·1 | 79·3-85 | 160 | 94·6 | 91·4-97·8 | p<0·001*** |
|  | Yes | 764 | 20·8 | 18·5-23·1 | 277 | 33·2 | 28·8-37·6 | 478 | 17·9 | 15-20·7 | 9 | 5·4 | 2·2-8·6 |  |
|  | *Missing* | *5* | *0·1* |  | *1* | *0·1* |  | *3* | *0·1* |  | *1* | *0·6* |  |  |
| **Source of income - undeclared employment** | No | 3,085 | 83·8 | 82-85·7 | 742 | 88·9 | 86·3-91·5 | 2,212 | 82·6 | 80·3-85 | 132 | 77·9 | 70·8-85 | p<0·001*** |
|  | Yes | 595 | 16·2 | 14·3-18 | 93 | 11·1 | 8·5-13·7 | 465 | 17·4 | 15-19·7 | 37 | 22·1 | 15-29·2 |  |
|  | *Missing* | *5* | *0·1* |  | *1* | *0·1* |  | *3* | *0·1* |  | *1* | *0·6* |  |  |
| **Source of income - retirement pension and/or alimony** | No | 3,451 | 93·8 | 92·5-95 | 679 | 81·3 | 77·4-85·2 | 2,605 | 97·3 | 96·3-98·4 | 167 | 99·1 | 98·1-100·1 | p<0·001*** |
|  | Yes | 229 | 6·2 | 5-7·5 | 156 | 18·7 | 14·8-22·6 | 71 | 2·7 | 1·6-3·7 | 1 | 0·9 | -0·1-1·9 |  |
|  | *Missing* | *5* | *0·1* |  | *1* | *0·1* |  | *3* | *0·1* |  | *1* | *0·6* |  |  |
| **Source of income - unemployment benefit** | No | 3,578 | 97·2 | 96·5-97·9 | 789 | 94·5 | 92·6-96·3 | 2,621 | 98·0 | 97·2-98·7 | 167 | 99·2 | 98·4-100 | p<0·001*** |
|  | Yes | 102 | 2·8 | 2·1-3·5 | 46 | 5·5 | 3·7-7·4 | 55 | 2·0 | 1·3-2·8 | 1 | 0·8 | 0-1·6 |  |
|  | *Missing* | *5* | *0·1* |  | *1* | *0·1* |  | *3* | *0·1* |  | *1* | *0·6* |  |  |
| **Source of income - scholarship** | No | 3,662 | 99·5 | 99·2-99·8 | 831 | 99·5 | 98·9-100·1 | 2,662 | 99·5 | 99·1-99·8 | 169 | 100·0 | 100-100 | 0·739 |
|  | Yes | 18 | 0·5 | 0·2-0·8 | 4 | 0·5 | -0·1-1·1 | 14 | 0·5 | 0·2-0·9 |  |  |  |  |
|  | *Missing* | *5* | *0·1* |  | *1* | *0·1* |  | *3* | *0·1* |  | *1* | *0·6* |  |  |
| **Source of income - social solidarity benefit** | No | 3,253 | 88·4 | 86·6-90·2 | 717 | 85·8 | 81·4-90·3 | 2,381 | 89·0 | 87-90·9 | 156 | 92·1 | 88·3-95·9 | 0·155· |
|  | Yes | 427 | 11·6 | 9·8-13·4 | 118 | 14·2 | 9·7-18·6 | 295 | 11·0 | 9·1-13 | 13 | 7·9 | 4·1-11·7 |  |
|  | *Missing* | *5* | *0·1* |  | *1* | *0·1* |  | *3* | *0·1* |  | *1* | *0·6* |  |  |
| **Source of income - other(s) benefit(s)** | No | 2,971 | 80·7 | 78·2-83·3 | 708 | 84·7 | 80·2-89·3 | 2,11 | 78·8 | 75·7-82 | 153 | 90·6 | 86·6-94·7 | 0·009** |
|  | Yes | 709 | 19·3 | 16·7-21·8 | 127 | 15·3 | 10·7-19·8 | 566 | 21·2 | 18-24·3 | 16 | 9·4 | 5·3-13·4 |  |
|  | *Missing* | *5* | *0·1* |  | *1* | *0·1* |  | *3* | *0·1* |  | *1* | *0·6* |  |  |
| **Source of income - begging** | No | 3,611 | 98·1 | 97·6-98·7 | 833 | 99·7 | 99·3-100·1 | 2,662 | 99·5 | 99·1-99·8 | 116 | 68·9 | 60·5-77·3 | p<0·001*** |
|  | Yes | 69 | 1·9 | 1·3-2·4 | 3 | 0·3 | -0·1-0·7 | 14 | 0·5 | 0·2-0·9 | 53 | 31·1 | 22·7-39·5 |  |
|  | *Missing* | *5* | *0·1* |  | *1* | *0·1* |  | *3* | *0·1* |  | *1* | *0·6* |  |  |
| **Source of income - associative/housing support** | No | 3,141 | 85·4 | 82·8-87·9 | 819 | 98·1 | 96·5-99·7 | 2,178 | 81·4 | 78-84·8 | 143 | 84·8 | 77·6-92·1 | p<0·001*** |
|  | Yes | 539 | 14·6 | 12·1-17·2 | 16 | 1·9 | 0·3-3·5 | 498 | 18·6 | 15·2-22 | 26 | 15·2 | 7·9-22·4 |  |
|  | *Missing* | *5* | *0·1* |  | *1* | *0·1* |  | *3* | *0·1* |  | *1* | *0·6* |  |  |
| **Source of income - family/friend support** | No | 3,433 | 93·3 | 91·9-94·7 | 806 | 96·6 | 95·2-97·9 | 2,468 | 92·2 | 90·4-94·1 | 158 | 93·9 | 90·8-96·9 | p<0·001*** |
|  | Yes | 247 | 6·7 | 5·3-8·1 | 29 | 3·4 | 2·1-4·8 | 208 | 7·8 | 5·9-9·6 | 10 | 6·1 | 3·1-9·2 |  |
|  | *Missing* | *5* | *0·1* |  | *1* | *0·1* |  | *3* | *0·1* |  | *1* | *0·6* |  |  |
| **Source of income - none** | No | 2,837 | 77·1 | 74·5-79·7 | 769 | 92·1 | 90·1-94·1 | 1,961 | 73·3 | 69·9-76·7 | 107 | 63·3 | 55·4-71·1 | p<0·001*** |
|  | Yes | 843 | 22·9 | 20·3-25·5 | 66 | 7·9 | 5·9-9·9 | 715 | 26·7 | 23·3-30·1 | 62 | 36·7 | 28·9-44·6 |  |
|  | *Missing* | *5* | *0·1* |  | *1* | *0·1* |  | *3* | *0·1* |  | *1* | *0·6* |  |  |
| **Source of income - unknown** | No | 3,66 | 99·5 | 99·1-99·8 | 835 | 100·0 | 99·9-100 | 2,659 | 99·3 | 98·9-99·8 | 166 | 98·5 | 96·7-100·4 | 0·015* |
|  | Yes | 20 | 0·5 | 0·2-0·9 | 0 | 0·0 | 0-0·1 | 17 | 0·7 | 0·2-1·1 | 2 | 1·5 | -0·4-3·3 |  |
|  | *Missing* | *5* | *0·1* |  | *1* | *0·1* |  | *3* | *0·1* |  | *1* | *0·6* |  |  |
| **Support and coping systems** | | | | | | | | | | | | | | |
| **Attends food distributions on a regular basis** | No | 2,246 | 61·5 | 57·6-65·4 | 729 | 88·6 | 85·5-91·8 | 1,434 | 53·9 | 48·8-59 | 82 | 49·3 | 40·4-58·1 | p<0·001*** |
|  | Yes | 1,407 | 38·5 | 34·6-42·4 | 94 | 11·4 | 8·2-14·5 | 1,229 | 46·1 | 41-51·2 | 85 | 50·7 | 41·9-59·6 |  |
|  | *Missing* | *34* | *0·9* |  | *14* | *1·7* |  | *17* | *0·6* |  | *3* | *1·9* |  |  |
| **Benefits from outreach activities** | No | 3,335 | 91·3 | 89·5-93·1 | 802 | 97·4 | 96·1-98·8 | 2,438 | 91·5 | 89·2-93·9 | 95 | 56·7 | 47·7-65·7 | p<0·001*** |
|  | Yes | 318 | 8·7 | 6·9-10·5 | 21 | 2·6 | 1·2-3·9 | 225 | 8·5 | 6·1-10·8 | 72 | 43·3 | 34·3-52·3 |  |
|  | *Missing* | *34* | *0·9* |  | *14* | *1·7* |  | *17* | *0·6* |  | *3* | *1·9* |  |  |
| **Supported by site managers/social workers on site** | No | 2,433 | 66·6 | 61·8-71·4 | 718 | 87·2 | 82·1-92·4 | 1,566 | 58·8 | 52·6-65 | 149 | 89·3 | 83-95·6 | p<0·001*** |
|  | Yes | 1,22 | 33·4 | 28·6-38·2 | 105 | 12·8 | 7·6-17·9 | 1,097 | 41·2 | 35-47·4 | 18 | 10·7 | 4·4-17 |  |
|  | *Missing* | *34* | *0·9* |  | *14* | *1·7* |  | *17* | *0·6* |  | *3* | *1·9* |  |  |
| **Supported by social workers (external)** | No | 2,79 | 76·4 | 72·9-79·9 | 668 | 81·2 | 76·1-86·2 | 1,997 | 75·0 | 70·5-79·4 | 126 | 75·3 | 68-82·6 | 0·111· |
|  | Yes | 863 | 23·6 | 20·1-27·1 | 155 | 18·8 | 13·8-23·9 | 667 | 25·0 | 20·6-29·5 | 41 | 24·7 | 17·4-32 |  |
|  | *Missing* | *34* | *0·9* |  | *14* | *1·7* |  | *17* | *0·6* |  | *3* | *1·9* |  |  |
| **Supported by state/city services** | No | 3,588 | 98·2 | 97·5-98·9 | 808 | 98·2 | 97-99·5 | 2,614 | 98·1 | 97·2-99 | 166 | 99·6 | 98·7-100·4 | 0·499 |
|  | Yes | 65 | 1·8 | 1·1-2·5 | 14 | 1·8 | 0·5-3 | 50 | 1·9 | 1-2·8 | 1 | 0·4 | -0·4-1·3 |  |
|  | *Missing* | *34* | *0·9* |  | *14* | *1·7* |  | *17* | *0·6* |  | *3* | *1·9* |  |  |
| **Supported by health professionals** | No | 3,423 | 93·7 | 92·2-95·2 | 790 | 96·0 | 94·1-98 | 2,474 | 92·9 | 90·9-94·9 | 160 | 95·8 | 93·4-98·2 | 0·034* |
|  | Yes | 230 | 6·3 | 4·8-7·8 | 33 | 4·0 | 2-5·9 | 190 | 7·1 | 5·1-9·1 | 7 | 4·2 | 1·8-6·6 |  |
|  | *Missing* | *34* | *0·9* |  | *14* | *1·7* |  | *17* | *0·6* |  | *3* | *1·9* |  |  |
| **Supported by religieux groups/community** | No | 3,471 | 95·0 | 93·8-96·2 | 792 | 96·3 | 94·1-98·4 | 2,526 | 94·8 | 93·3-96·4 | 152 | 91·4 | 85·7-97 | 0·2 |
|  | Yes | 182 | 5·0 | 3·8-6·2 | 31 | 3·7 | 1·6-5·9 | 137 | 5·2 | 3·6-6·7 | 14 | 8·6 | 3-14·3 |  |
|  | *Missing* | *34* | *0·9* |  | *14* | *1·7* |  | *17* | *0·6* |  | *3* | *1·9* |  |  |
| **Autonomous / Does not receive or need support whatsoever** | No | 2,593 | 71·0 | 67·6-74·3 | 317 | 38·6 | 32·3-44·8 | 2,153 | 80·8 | 77-84·7 | 122 | 73·3 | 65·7-80·9 | p<0·001*** |
|  | Yes | 1,06 | 29·0 | 25·7-32·4 | 506 | 61·4 | 55·2-67·7 | 510 | 19·2 | 15·3-23 | 44 | 26·7 | 19·1-34·3 |  |
|  | *Missing* | *34* | *0·9* |  | *14* | *1·7* |  | *17* | *0·6* |  | *3* | *1·9* |  |  |
| **Benefits from support to attend and navigate the healthcare system** | Yes (relatives, social workers etc) | 1,074 | 29·4 | 26·4-32·4 | 195 | 23·5 | 18·1-29 | 830 | 31·2 | 27·4-35 | 50 | 30·1 | 22·6-37·6 | p<0·001*** |
|  | I need help but I go by myself | 1,391 | 38·0 | 34·2-41·9 | 284 | 34·3 | 27·3-41·2 | 1,03 | 38·7 | 33·9-43·5 | 77 | 46·5 | 38-55·1 |  |
|  | Autonomous/independant | 1,122 | 30·7 | 26·3-35·1 | 336 | 40·6 | 31·8-49·4 | 760 | 28·5 | 23·2-33·9 | 26 | 15·8 | 10·7-20·8 |  |
|  | I never consult/It never happened so far | 68 | 1·9 | 1·3-2·4 | 13 | 1·6 | 0·6-2·6 | 42 | 1·6 | 0·9-2·3 | 12 | 7·6 | 3·9-11·3 |  |
|  | *Missing* | *23* | *0·6* |  | *7* | *0·9* |  | *11* | *0·4* |  | *5* | *2·9* |  |  |
| **Moral and social support** | | | | | | | | | | | | | | |
| **Support from relatives (can speak to/be listened by relatives)** | Yes, often | 1,335 | 36·6 | 33·5-39·6 | 361 | 43·6 | 37-50·1 | 944 | 35·5 | 31·8-39·1 | 31 | 18·7 | 13·2-24·3 | p<0·001*** |
|  | Yes, sometimes | 1,287 | 35·2 | 32·4-38·1 | 282 | 34·0 | 27·6-40·5 | 947 | 35·6 | 32·3-38·9 | 58 | 35·0 | 28·4-41·5 |  |
|  | No, almost never | 942 | 25·8 | 23·6-28 | 166 | 20·1 | 15·8-24·4 | 709 | 26·7 | 24-29·4 | 66 | 40·1 | 32·4-47·8 |  |
|  | NA: situation never occurred | 88 | 2·4 | 1·8-3·1 | 19 | 2·3 | 0·9-3·7 | 59 | 2·2 | 1·4-3 | 10 | 6·2 | 3·4-9·1 |  |
|  | *Missing* | *27* | *0·7* |  | *7* | *0·8* |  | *16* | *0·6* |  | *4* | *2·4* |  |  |
| **Material support from relatives (money, food, clothes etc)** | Yes, often | 479 | 13·2 | 11·4-15 | 153 | 18·5 | 13·9-23 | 305 | 11·5 | 9·5-13·5 | 21 | 13·0 | 8·1-17·9 | 0·001** |
|  | Yes, sometimes | 1,031 | 28·3 | 25·9-30·8 | 228 | 27·5 | 23·3-31·8 | 755 | 28·5 | 25·4-31·6 | 48 | 29·3 | 23·1-35·6 |  |
|  | No, almost never | 1,926 | 52·9 | 50·1-55·7 | 375 | 45·4 | 40·2-50·6 | 1,467 | 55·4 | 52-58·9 | 83 | 50·7 | 42·7-58·7 |  |
|  | NA: situation never occurred | 202 | 5·6 | 3·9-7·2 | 71 | 8·6 | 4·9-12·4 | 120 | 4·5 | 2·6-6·4 | 11 | 7·0 | 3·8-10·1 |  |
|  | *Missing* | *40* | *1·1* |  | *8* | *0·9* |  | *27* | *1·0* |  | *5* | *3·0* |  |  |
| **Advices from relatives (to find a job etc)** | Yes, often | 858 | 23·6 | 21·1-26·2 | 276 | 33·5 | 26·9-40·1 | 563 | 21·3 | 18·5-24 | 19 | 11·7 | 7·5-15·9 | p<0·001*** |
|  | Yes, sometimes | 1,442 | 39·7 | 36·8-42·7 | 308 | 37·4 | 31·1-43·8 | 1,082 | 40·9 | 37·4-44·4 | 52 | 32·3 | 25·6-39 |  |
|  | No, almost never | 1,202 | 33·1 | 30·6-35·6 | 205 | 24·9 | 20·4-29·3 | 922 | 34·9 | 31·8-38 | 76 | 47·0 | 38·7-55·3 |  |
|  | NA: situation never occurred | 127 | 3·5 | 2·6-4·4 | 35 | 4·2 | 1·8-6·6 | 77 | 2·9 | 1·9-3·9 | 14 | 8·9 | 5-12·9 |  |
|  | *Missing* | *50* | *1·4* |  | *11* | *1·4* |  | *30* | *1·1* |  | *9* | *5·0* |  |  |
| **Moral support by relatives (motivation)** | Yes, often | 938 | 26·0 | 23·2-28·8 | 267 | 32·6 | 26·3-39 | 648 | 24·6 | 21·4-27·9 | 23 | 13·9 | 8·9-18·8 | 0·004** |
|  | Yes, sometimes | 1,427 | 39·5 | 36·3-42·7 | 316 | 38·6 | 32·3-44·9 | 1,059 | 40·2 | 36·4-44·1 | 52 | 32·2 | 25·2-39·2 |  |
|  | No, almost never | 1,058 | 29·3 | 26·8-31·7 | 186 | 22·8 | 18·4-27·2 | 796 | 30·3 | 27·2-33·3 | 76 | 46·4 | 37·9-55 |  |
|  | NA: situation never occurred | 189 | 5·2 | 3·6-6·9 | 49 | 6·0 | 2-9·9 | 128 | 4·8 | 3-6·7 | 12 | 7·5 | 3·9-11·1 |  |
|  | *Missing* | *69* | *1·9* |  | *17* | *2·1* |  | *45* | *1·7* |  | *7* | *4·1* |  |  |
| **Support from organization members (can speak to/be listened by members)** | Yes, often | 520 | 14·3 | 12·1-16·4 | 42 | 5·1 | 2·8-7·5 | 471 | 17·7 | 15-20·5 | 7 | 4·4 | 2·3-6·5 | p<0·001*** |
|  | Yes, sometimes | 1,194 | 32·8 | 30·1-35·5 | 136 | 16·5 | 13·3-19·7 | 1,004 | 37·8 | 34·4-41·2 | 54 | 33·0 | 25·1-40·8 |  |
|  | No, almost never | 1,622 | 44·5 | 41·4-47·7 | 502 | 61·1 | 54·6-67·7 | 1,042 | 39·3 | 35·7-42·8 | 78 | 46·9 | 38·7-55·1 |  |
|  | NA: situation never occurred | 305 | 8·4 | 6·8-9·9 | 141 | 17·2 | 11·9-22·5 | 137 | 5·2 | 3·9-6·5 | 26 | 15·7 | 9·7-21·8 |  |
|  | *Missing* | *36* | *1·0* |  | *13* | *1·5* |  | *19* | *0·7* |  | *4* | *2·5* |  |  |
| **Material support from org· members (money, food, clothes etc)** | Yes, often | 555 | 15·3 | 12·7-17·8 | 29 | 3·5 | 1·7-5·3 | 506 | 19·1 | 15·8-22·5 | 19 | 11·7 | 6·8-16·7 | p<0·001*** |
|  | Yes, sometimes | 1,132 | 31·1 | 28·1-34·1 | 117 | 14·1 | 11·1-17·2 | 961 | 36·3 | 32·5-40·1 | 54 | 32·9 | 25·1-40·7 |  |
|  | No, almost never | 1,653 | 45·4 | 42-48·9 | 529 | 64·2 | 57·7-70·6 | 1,056 | 39·9 | 35·7-44 | 68 | 41·4 | 33·7-49·2 |  |
|  | NA: situation never occurred | 299 | 8·2 | 6·4-10 | 150 | 18·2 | 12·6-23·7 | 126 | 4·7 | 3·2-6·3 | 23 | 13·9 | 7·8-20 |  |
|  | *Missing* | *43* | *1·2* |  | *11* | *1·3* |  | *27* | *1·0* |  | *5* | *3·1* |  |  |
| **Advices from org· members (to find a job etc)** | Yes, often | 515 | 14·2 | 11·9-16·6 | 63 | 7·7 | 4·9-10·5 | 445 | 16·9 | 13·8-19·9 | 7 | 4·5 | 2·1-6·9 | p<0·001*** |
|  | Yes, sometimes | 1,291 | 35·6 | 32·6-38·6 | 144 | 17·6 | 13·8-21·3 | 1,102 | 41·7 | 38-45·5 | 45 | 27·8 | 20·2-35·4 |  |
|  | No, almost never | 1,557 | 42·9 | 39·6-46·3 | 493 | 60·0 | 53·2-66·7 | 983 | 37·2 | 33·3-41·1 | 81 | 50·3 | 42·2-58·4 |  |
|  | NA: situation never occurred | 261 | 7·2 | 5·7-8·7 | 122 | 14·8 | 9·7-19·9 | 112 | 4·2 | 3-5·4 | 28 | 17·3 | 10·9-23·8 |  |
|  | *Missing* | *52* | *1·4* |  | *13* | *1·5* |  | *31* | *1·2* |  | *8* | *4·7* |  |  |
| **Moral support by org· members (motivation)** | Yes, often | 378 | 10·5 | 8·4-12·5 | 36 | 4·5 | 2·5-6·4 | 335 | 12·8 | 10·1-15·5 | 6 | 3·7 | 1·4-5·9 | p<0·001*** |
|  | Yes, sometimes | 1,075 | 29·8 | 26·9-32·8 | 106 | 13·0 | 9·1-16·9 | 926 | 35·2 | 31·5-38·9 | 44 | 27·0 | 19·5-34·5 |  |
|  | No, almost never | 1,82 | 50·5 | 47-53·9 | 525 | 64·4 | 57·6-71·2 | 1,211 | 46·1 | 42-50·2 | 84 | 51·8 | 44-59·5 |  |
|  | NA: situation never occurred | 332 | 9·2 | 7·3-11·1 | 148 | 18·1 | 12·3-23·9 | 156 | 5·9 | 4·3-7·6 | 28 | 17·6 | 11·2-24 |  |
|  | *Missing* | *72* | *2·0* |  | *19* | *2·3* |  | *45* | *1·7* |  | *8* | *4·6* |  |  |
| **Self-reported feeling of loneliness** | I feel very lonely | 571 | 15·8 | 13·9-17·7 | 116 | 14·2 | 10·8-17·6 | 405 | 15·4 | 13·1-17·8 | 49 | 30·1 | 22·2-37·9 | p<0·001*** |
|  | I don't feel taken care of/supported· I feel lonely sometimes· | 1,194 | 33·1 | 30·9-35·3 | 282 | 34·5 | 30·1-38·9 | 854 | 32·5 | 29·8-35·2 | 58 | 35·5 | 28·2-42·9 |  |
|  | I feel taken care of/supported/not specially lonely | 1,375 | 38·1 | 35·6-40·7 | 278 | 34·0 | 29·2-38·7 | 1,051 | 40·0 | 36·9-43·1 | 46 | 28·3 | 21·2-35·4 |  |
|  | I feel very taken care of/supported· I don't feel lonely at all | 467 | 12·9 | 11·2-14·7 | 141 | 17·3 | 13·3-21·3 | 316 | 12·0 | 10-14·1 | 10 | 6·1 | 2·9-9·3 |  |
|  | *Missing* | *72* | *2·0* |  | *17* | *2·1* |  | *49* | *1·8* |  | *6* | *3·5* |  |  |
| **COVID-19 (vaccination) Information Sources** | | | | | | | | | | | | | | |
| **Internet connection** | Yes, seldom or often uses internet | 2,857 | 78·1 | 76-80·1 | 537 | 64·9 | 60·4-69·3 | 2,236 | 83·9 | 81·6-86·2 | 84 | 50·4 | 42·7-58·2 | p<0·001*** |
|  | No, never uses internet | 794 | 21·7 | 19·6-23·8 | 289 | 34·9 | 30·4-39·3 | 425 | 16·0 | 13·7-18·2 | 80 | 48·0 | 40·1-55·8 |  |
|  | Other: another mean of connection | 9 | 0·2 | 0·1-0·4 | 2 | 0·3 | -0·1-0·6 | 3 | 0·1 | 0-0·3 | 3 | 1·6 | -0·3-3·5 |  |
|  | *Missing* | *25* | *0·7* |  | *8* | *0·9* |  | *14* | *0·5* |  | *3* | *2·0* |  |  |
| **Compliance to protection measures (wear a facemask in public transportation etc)** | Yes, I wear it and I think it's useful | 2,882 | 81·1 | 79-83·1 | 682 | 84·3 | 80·9-87·7 | 2,083 | 80·6 | 78-83·2 | 117 | 72·7 | 65·7-79·8 | 0·035* |
|  | It depends on the context | 266 | 7·5 | 6·2-8·8 | 45 | 5·5 | 3·4-7·6 | 206 | 8·0 | 6·3-9·7 | 15 | 9·2 | 5·4-12·9 |  |
|  | No, this is useless | 407 | 11·5 | 9·9-13·1 | 82 | 10·2 | 7·5-12·9 | 296 | 11·4 | 9·4-13·4 | 29 | 18·1 | 12·6-23·6 |  |
|  | *Missing* | *126* | *3·4* |  | *26* | *3·1* |  | *91* | *3·4* |  | *9* | *5·3* |  |  |
| **COVID-19 (Vaccine) information source - internet / social media** | No | 2,342 | 63·5 | 60·9-66·1 | 558 | 66·7 | 61·6-71·9 | 1,665 | 62·1 | 59-65·2 | 119 | 70·2 | 63·5-77 | 0·092· |
|  | Yes | 1,345 | 36·5 | 33·9-39·1 | 278 | 33·3 | 28·1-38·4 | 1,016 | 37·9 | 34·8-41 | 51 | 29·8 | 23-36·5 |  |
|  | *Missing* | *0* | *0·0* |  | *0* | *0·0* |  | *0* | *0·0* |  | *0* | *0·0* |  |  |
| **COVID-19 (Vaccine) information source - television/radio** | No | 1,171 | 31·8 | 29·4-34·3 | 238 | 28·5 | 23·5-33·5 | 832 | 31·1 | 28·2-34 | 101 | 60·1 | 52·2-68 | p<0·001*** |
|  | Yes | 2,507 | 68·2 | 65·7-70·6 | 596 | 71·5 | 66·5-76·5 | 1,843 | 68·9 | 66-71·8 | 67 | 39·9 | 32-47·8 |  |
|  | *Missing* | *4* | *0·1* |  | *2* | *0·2* |  | *1* | *0·0* |  | *1* | *0·6* |  |  |
| **COVID-19 (Vaccine) information source - print media/classic media** | No | 3,46 | 94·1 | 92·8-95·4 | 784 | 94·0 | 91·7-96·3 | 2,528 | 94·5 | 92·9-96·1 | 148 | 87·7 | 82·4-92·9 | 0·033* |
|  | Yes | 218 | 5·9 | 4·6-7·2 | 50 | 6·0 | 3·7-8·3 | 147 | 5·5 | 3·9-7·1 | 21 | 12·3 | 7·1-17·6 |  |
|  | *Missing* | *4* | *0·1* |  | *2* | *0·2* |  | *1* | *0·0* |  | *1* | *0·6* |  |  |
| **COVID-19 (Vaccine) information source - posters/leaflets** | No | 3,485 | 94·7 | 93·4-96 | 791 | 94·9 | 92·8-97 | 2,532 | 94·6 | 93-96·3 | 162 | 95·7 | 92·9-98·5 | 0·846 |
|  | Yes | 193 | 5·3 | 4-6·6 | 43 | 5·1 | 3-7·2 | 144 | 5·4 | 3·7-7 | 7 | 4·3 | 1·5-7·1 |  |
|  | *Missing* | *4* | *0·1* |  | *2* | *0·2* |  | *1* | *0·0* |  | *1* | *0·6* |  |  |
| **COVID-19 (Vaccine) information source - site manager** | No | 3,483 | 94·7 | 93·2-96·2 | 784 | 94·1 | 91·3-96·8 | 2,531 | 94·6 | 92·7-96·5 | 168 | 99·4 | 98·8-100 | 0·132· |
|  | Yes | 195 | 5·3 | 3·8-6·8 | 50 | 5·9 | 3·2-8·7 | 145 | 5·4 | 3·5-7·3 | 1 | 0·6 | 0-1·2 |  |
|  | *Missing* | *4* | *0·1* |  | *2* | *0·2* |  | *1* | *0·0* |  | *1* | *0·6* |  |  |
| **COVID-19 (Vaccine) information source - health or social workers/professionnals** | No | 2,953 | 80·1 | 77·4-82·8 | 723 | 86·3 | 83·1-89·6 | 2,092 | 78·0 | 74·6-81·5 | 139 | 81·6 | 75·3-87·9 | 0·001** |
|  | Yes | 734 | 19·9 | 17·2-22·6 | 114 | 13·7 | 10·4-16·9 | 588 | 22·0 | 18·5-25·4 | 31 | 18·4 | 12·1-24·7 |  |
|  | *Missing* | *0* | *0·0* |  | *0* | *0·0* |  | *0* | *0·0* |  | *0* | *0·0* |  |  |
| **COVID-19 (Vaccine) information source - relatives/family/community** | No | 2,572 | 69·8 | 67·5-72·1 | 545 | 65·1 | 60·4-69·9 | 1,941 | 72·4 | 69·7-75·1 | 86 | 50·6 | 42·3-58·9 | p<0·001*** |
|  | Yes | 1,115 | 30·2 | 27·9-32·5 | 292 | 34·9 | 30·1-39·6 | 739 | 27·6 | 24·9-30·3 | 84 | 49·4 | 41·1-57·7 |  |
|  | *Missing* | *0* | *0·0* |  | *0* | *0·0* |  | *0* | *0·0* |  | *0* | *0·0* |  |  |
| **COVID-19 (Vaccine) information source - none** | No | 3,607 | 98·1 | 97·5-98·7 | 816 | 97·9 | 97-98·8 | 2,637 | 98·6 | 97·9-99·3 | 153 | 90·9 | 86·2-95·7 | p<0·001*** |
|  | Yes | 71 | 1·9 | 1·3-2·5 | 17 | 2·1 | 1·2-3 | 38 | 1·4 | 0·7-2·1 | 15 | 9·1 | 4·3-13·8 |  |
|  | *Missing* | *4* | *0·1* |  | *2* | *0·2* |  | *1* | *0·0* |  | *1* | *0·6* |  |  |
| **Satisfaction with COVID-19 information (overall)** | No | 1,712 | 46·4 | 43·8-49·1 | 356 | 42·6 | 37-48·1 | 1,259 | 47·0 | 43·7-50·2 | 97 | 57·1 | 49·8-64·4 | 0·002** |
|  | Yes | 1,79 | 48·5 | 45·8-51·3 | 440 | 52·6 | 46·9-58·2 | 1,295 | 48·3 | 45·1-51·6 | 55 | 32·3 | 25·2-39·3 |  |
|  | DNK/DNA | 185 | 5·0 | 4·1-5·9 | 41 | 4·9 | 3·2-6·5 | 126 | 4·7 | 3·6-5·8 | 18 | 10·6 | 6·6-14·7 |  |
|  | *Missing* | *0* | *0·0* |  | *0* | *0·0* |  | *0* | *0·0* |  | *0* | *0·0* |  |  |
| **Trust in the crisis management by the authorities** | Zero trust (0) | 218 | 5·9 | 5-6·9 | 38 | 4·5 | 3-6·1 | 145 | 5·4 | 4·3-6·5 | 36 | 21·2 | 15·2-27·3 | p<0·001*** |
|  | Low trust (1-3) | 258 | 7·0 | 6·1-8 | 52 | 6·3 | 4·6-8 | 186 | 7·0 | 5·8-8·1 | 20 | 11·8 | 7·4-16·1 |  |
|  | Medium trust (4-6) | 849 | 23·1 | 21·4-24·9 | 189 | 22·7 | 19·6-25·8 | 630 | 23·6 | 21·4-25·8 | 30 | 17·7 | 13·1-22·3 |  |
|  | High trust (7-9) | 1,142 | 31·1 | 28·8-33·4 | 264 | 31·8 | 27·2-36·4 | 849 | 31·8 | 29-34·6 | 29 | 17·0 | 12·3-21·7 |  |
|  | Total trust (10) | 748 | 20·4 | 18·4-22·4 | 169 | 20·3 | 16·5-24·1 | 541 | 20·3 | 17·8-22·7 | 38 | 22·6 | 15·4-29·8 |  |
|  | No opinion | 456 | 12·4 | 10·8-14·1 | 121 | 14·5 | 11-18 | 319 | 11·9 | 10-13·9 | 16 | 9·7 | 5·3-14·1 |  |
|  | *Missing* | *10* | *0·3* |  | *3* | *0·3* |  | *6* | *0·2* |  | *1* | *0·6* |  |  |
| **Health-related information** | | | | | | | | | | | | | | |
| **Has medical coverage** | Yes (national social security, state medical aid) | 3,152 | 86·1 | 84·5-87·6 | 738 | 88·8 | 86·1-91·5 | 2,335 | 87·6 | 85·8-89·4 | 78 | 47·3 | 39·8-54·9 | p<0·001*** |
|  | No (never, lost or ongoing application) | 510 | 13·9 | 12·4-15·5 | 93 | 11·2 | 8·5-13·9 | 330 | 12·4 | 10·6-14·2 | 87 | 52·7 | 45·1-60·2 |  |
|  | *Missing* | *16* | *0·4* |  | *4* | *0·4* |  | *8* | *0·3* |  | *4* | *2·5* |  |  |
| **Self-reported health status** | Bad status overall | 762 | 21·0 | 18·8-23·2 | 152 | 18·6 | 15·1-22·1 | 561 | 21·2 | 18·5-23·9 | 48 | 29·9 | 23·9-35·9 | 0·152· |
|  | Good status overall | 1,76 | 48·5 | 45·8-51·2 | 408 | 49·7 | 44·3-55·1 | 1,288 | 48·6 | 45·3-52 | 65 | 40·1 | 32·5-47·7 |  |
|  | Perfect condition | 1,107 | 30·5 | 28-33 | 260 | 31·7 | 26·7-36·7 | 799 | 30·2 | 27·1-33·2 | 48 | 30·0 | 23·4-36·5 |  |
|  | *Missing* | *49* | *1·3* |  | *15* | *1·8* |  | *26* | *1·0* |  | *8* | *4·5* |  |  |
| **Chronic condition (self-reported)** | No | 2,561 | 70·2 | 67·5-72·9 | 558 | 67·5 | 62·9-72·1 | 1,872 | 70·5 | 67·1-73·9 | 130 | 79·5 | 74·3-84·7 | 0·066· |
|  | Yes | 1,086 | 29·8 | 27·1-32·5 | 269 | 32·5 | 27·9-37·1 | 784 | 29·5 | 26·1-32·9 | 34 | 20·5 | 15·3-25·7 |  |
|  | *Missing* | *31* | *0·8* |  | *8* | *0·9* |  | *17* | *0·6* |  | *6* | *3·5* |  |  |
| **Is followed by a regular GP** | Yes | 2,465 | 67·4 | 65-69·7 | 629 | 75·8 | 72·1-79·5 | 1,791 | 67·2 | 64·3-70·2 | 45 | 27·5 | 21-33·9 | p<0·001*** |
|  | No | 1,193 | 32·6 | 30·3-35 | 201 | 24·2 | 20·5-27·9 | 872 | 32·8 | 29·8-35·7 | 120 | 72·5 | 66·1-79 |  |
|  | *Missing* | *18* | *0·5* |  | *4* | *0·5* |  | *9* | *0·3* |  | *5* | *2·7* |  |  |
| **Date of last consultation (hospital/GP)** | After the start of vaccination campaign (>may 2021) | 2,595 | 71·3 | 68·8-73·7 | 563 | 68·3 | 63·5-73·1 | 1,949 | 73·5 | 70·5-76·4 | 83 | 50·5 | 43·2-57·9 | p<0·001*** |
|  | Between 2d lockdown and start of vaccination (dec 2020-may 2021) | 334 | 9·2 | 7·8-10·6 | 85 | 10·3 | 7·9-12·6 | 237 | 8·9 | 7·1-10·7 | 13 | 8·0 | 4·7-11·2 |  |
|  | Between 1st and 2d lockdown (mar-dec 2020) | 387 | 10·6 | 8·6-12·6 | 94 | 11·3 | 7·5-15·2 | 277 | 10·4 | 7·9-12·9 | 16 | 10·0 | 6·5-13·5 |  |
|  | Before the pandemic (<march 2020) | 203 | 5·6 | 4·5-6·6 | 58 | 7·1 | 4·9-9·2 | 125 | 4·7 | 3·5-6 | 19 | 11·7 | 7·3-16·1 |  |
|  | Never consulted in France/Does not apply | 123 | 3·4 | 2·6-4·2 | 25 | 3·0 | 1·8-4·3 | 66 | 2·5 | 1·6-3·3 | 32 | 19·8 | 13·2-26·3 |  |
|  | *Missing* | *28* | *0·8* |  | *8* | *1·0* |  | *15* | *0·6* |  | *5* | *2·9* |  |  |
| **Previous history/hospitalization of COVID-19** | No | 2,991 | 81·4 | 79·7-83·1 | 702 | 84·2 | 81·4-87·1 | 2,137 | 80·0 | 77·9-82 | 152 | 90·2 | 86·3-94·2 | 0·003** |
|  | Yes, without hospitalization | 588 | 16·0 | 14·5-17·5 | 111 | 13·3 | 10·8-15·8 | 462 | 17·3 | 15·4-19·2 | 15 | 9·0 | 5·3-12·8 |  |
|  | Yes, with hospitalization | 96 | 2·6 | 1·9-3·3 | 21 | 2·5 | 1·2-3·7 | 74 | 2·8 | 1·9-3·6 | 1 | 0·7 | -0·1-1·5 |  |
|  | *Missing* | *10* | *0·3* |  | *3* | *0·3* |  | *7* | *0·2* |  | *1* | *0·7* |  |  |
| **Previous history/hospitalization for COVID-19 among family or friends** | No | 1,988 | 53·9 | 51·4-56·4 | 504 | 60·3 | 56-64·5 | 1,378 | 51·4 | 48·3-54·5 | 105 | 61·8 | 54·4-69·3 | p<0·001*** |
|  | Yes, without hospitalization | 959 | 26·0 | 23·8-28·2 | 157 | 18·8 | 15·1-22·4 | 767 | 28·6 | 25·9-31·3 | 35 | 20·3 | 14·9-25·8 |  |
|  | Yes, with hospitalization | 740 | 20·1 | 18·3-21·9 | 175 | 21·0 | 17·4-24·5 | 535 | 20·0 | 17·8-22·1 | 30 | 17·8 | 11·5-24·2 |  |
|  | *Missing* | *0* | *0·0* |  | *0* | *0·0* |  | *0* | *0·0* |  | *0* | *0·0* |  |  |
| **Vulnerable/at risk for COVID-19 people among relatives** | No | 2,419 | 65·8 | 62·7-68·8 | 511 | 61·1 | 55·6-66·6 | 1,815 | 67·8 | 64·1-71·6 | 94 | 55·6 | 47·5-63·7 | 0·015* |
|  | Yes | 1,26 | 34·2 | 31·2-37·3 | 325 | 38·9 | 33·4-44·4 | 860 | 32·2 | 28·4-35·9 | 75 | 44·4 | 36·3-52·5 |  |
|  | *Missing* | *4* | *0·1* |  | *1* | *0·1* |  | *3* | *0·1* |  | *1* | *0·5* |  |  |
| **Health literacy** | | | | | | | | | | | | | | |
| **Can read and understand drug notices** | With ease overall, alone | 2,037 | 55·8 | 53·2-58·5 | 422 | 50·9 | 45·8-56 | 1,544 | 58·2 | 55-61·3 | 72 | 43·5 | 35·7-51·2 | 0·002** |
|  | With difficulty overall, alone | 1,611 | 44·2 | 41·5-46·8 | 407 | 49·1 | 44-54·2 | 1,111 | 41·8 | 38·7-45 | 93 | 56·5 | 48·8-64·3 |  |
|  | *Missing* | *27* | *0·7* |  | *5* | *0·6* |  | *17* | *0·6* |  | *5* | *2·9* |  |  |
| **Can understand oral medical instructions** | With ease overall, alone | 2,86 | 78·3 | 75·9-80·8 | 654 | 79·0 | 73·4-84·5 | 2,113 | 79·5 | 76·7-82·3 | 93 | 56·3 | 48·1-64·5 | p<0·001*** |
|  | With difficulty overall, alone | 791 | 21·7 | 19·2-24·1 | 174 | 21·0 | 15·5-26·6 | 545 | 20·5 | 17·7-23·3 | 72 | 43·7 | 35·5-51·9 |  |
|  | *Missing* | *23* | *0·6* |  | *5* | *0·6* |  | *13* | *0·5* |  | *5* | *2·9* |  |  |
| **Can understand oral medical advice** | With ease overall, alone | 2,663 | 72·9 | 70·3-75·6 | 584 | 70·5 | 64·5-76·6 | 1,99 | 74·9 | 71·9-77·9 | 89 | 54·2 | 45·9-62·5 | 0·002** |
|  | With difficulty overall, alone | 988 | 27·1 | 24·4-29·7 | 244 | 29·5 | 23·4-35·5 | 668 | 25·1 | 22·1-28·1 | 75 | 45·8 | 37·5-54·1 |  |
|  | *Missing* | *23* | *0·6* |  | *5* | *0·6* |  | *13* | *0·5* |  | *5* | *2·9* |  |  |
| **Can read and understand written medical instructions** | With ease overall, alone | 2,152 | 59·0 | 56·3-61·6 | 439 | 52·9 | 47·9-58 | 1,64 | 61·8 | 58·6-65 | 73 | 44·3 | 36·6-52·1 | p<0·001*** |
|  | With difficulty overall, alone | 1,496 | 41·0 | 38·4-43·7 | 390 | 47·1 | 42-52·1 | 1,015 | 38·2 | 35-41·4 | 92 | 55·7 | 47·9-63·4 |  |
|  | *Missing* | *27* | *0·7* |  | *5* | *0·6* |  | *17* | *0·6* |  | *5* | *2·9* |  |  |
| **Can correctly fill out a medical form** | With ease overall, alone | 1,65 | 45·2 | 42·6-47·8 | 323 | 39·0 | 34·8-43·2 | 1,27 | 47·8 | 44·5-51 | 57 | 34·5 | 27·6-41·5 | p<0·001*** |
|  | With difficulty overall, alone | 2,001 | 54·8 | 52·2-57·4 | 506 | 61·0 | 56·8-65·2 | 1,388 | 52·2 | 49-55·5 | 108 | 65·5 | 58·5-72·4 |  |
|  | *Missing* | *23* | *0·6* |  | *5* | *0·6* |  | *13* | *0·5* |  | *5* | *2·9* |  |  |
| **Discrimination** | | | | | | | | | | | | | | |
| **Experienced discrimination since Covid-19 outbreak** | No | 2,714 | 73·6 | 70·9-76·3 | 664 | 79·3 | 75·3-83·3 | 1,93 | 72·0 | 68·5-75·5 | 120 | 70·7 | 63·4-78·1 | 0·032* |
|  | Yes | 590 | 16·0 | 13·8-18·2 | 93 | 11·2 | 8·2-14·1 | 462 | 17·2 | 14·4-20·1 | 34 | 20·3 | 14·2-26·3 |  |
|  | Does not know/Does not want to answer | 383 | 10·4 | 8·4-12·4 | 80 | 9·5 | 6·1-12·9 | 288 | 10·8 | 8·2-13·3 | 15 | 9·0 | 4·2-13·8 |  |
|  | *Missing* | *0* | *0·0* |  | *0* | *0·0* |  | *0* | *0·0* |  | *0* | *0·0* |  |  |
| **Is more afraid of being expelled from France since COVID-19 crisis** | No | 1,234 | 81·3 | 78·1-84·5 | 142 | 85·2 | 78·7-91·8 | 1,048 | 81·4 | 77·7-85 | 44 | 69·2 | 57·1-81·2 | 0·065· |
|  | Yes | 284 | 18·7 | 15·5-21·9 | 25 | 14·8 | 8·2-21·3 | 240 | 18·6 | 15-22·3 | 20 | 30·8 | 18·8-42·9 |  |
|  | *Missing* | *175* | *10·2* |  | *17* | *9·2* |  | *149* | *10·2* |  | *9* | *12·2* |  |  |
| **Has been denied vaccination** | No | 3,637 | 99·6 | 99·4-99·8 | 825 | 99·5 | 98·9-100·1 | 2,649 | 99·7 | 99·4-99·9 | 163 | 99·4 | 98·5-100·2 | 0·715 |
|  | Yes | 14 | 0·4 | 0·2-0·6 | 4 | 0·5 | -0·1-1·1 | 9 | 0·3 | 0·1-0·6 | 1 | 0·6 | -0·2-1·5 |  |
|  | *Missing* | *23* | *0·6* |  | *5* | *0·6* |  | *13* | *0·5* |  | *5* | *2·9* |  |  |
| **has been denied healthcare** | No | 3,445 | 94·6 | 93·7-95·6 | 798 | 96·8 | 95·5-98·1 | 2,503 | 94·2 | 93-95·4 | 144 | 90·2 | 86·5-93·9 | p<0·001*** |
|  | Yes | 196 | 5·4 | 4·4-6·3 | 26 | 3·2 | 1·9-4·5 | 154 | 5·8 | 4·6-7 | 16 | 9·8 | 6·1-13·5 |  |
|  | *Missing* | *28* | *0·7* |  | *8* | *0·9* |  | *11* | *0·4* |  | *9* | *5·4* |  |  |
| **Recruitment site information** | | | | | | | | | | | | | | |
| **Vaccine certificate is mandatory to access site** | No | 3,524 | 98·6 | 97·2-100 | 814 | 100·0 | 100-100 | 2,556 | 98·1 | 96·2-100 | 154 | 99·8 | 99·4-100·2 | 0·232 |
|  | Yes | 50 | 1·4 | 0-2·8 |  |  |  | 50 | 1·9 | 0-3·8 | 0 | 0·2 | -0·2-0·6 |  |
|  | *Missing* | *13* | *0·3* |  | *0* | *0·0* |  | *1* | *0·0* |  | *12* | *6·9* |  |  |
| **Presence of social workers on site** | At least once per week | 2,726 | 73·9 | 69·3-78·6 | 699 | 83·5 | 76·1-90·9 | 1,991 | 74·3 | 68·4-80·2 | 37 | 21·8 | 13·5-30·2 | p<0·001*** |
|  | Less than once per week | 825 | 22·4 | 17·8-26·9 | 128 | 15·3 | 8-22·7 | 645 | 24·1 | 18·3-29·8 | 52 | 30·7 | 21·7-39·7 |  |
|  | Unknown/Not applicable | 136 | 3·7 | 2·5-4·9 | 10 | 1·2 | 0·3-2 | 45 | 1·7 | 0·3-3 | 81 | 47·4 | 37·3-57·6 |  |
|  | *Missing* | *0* | *0·0* |  | *0* | *0·0* |  | *0* | *0·0* |  | *0* | *0·0* |  |  |
| **Presence of GP on site** | At least once per week | 365 | 9·9 | 6·6-13·1 | 77 | 9·2 | 2·8-15·5 | 278 | 10·4 | 6·4-14·4 | 9 | 5·5 | 1-9·9 | p<0·001*** |
|  | Less than once per week | 3,14 | 85·2 | 81·6-88·7 | 725 | 86·6 | 79·4-93·8 | 2,352 | 87·8 | 83·6-91·9 | 63 | 37·3 | 27·5-47·1 |  |
|  | Unknown/Not applicable | 183 | 5·0 | 3·5-6·4 | 35 | 4·2 | 0·5-8 | 50 | 1·9 | 0·5-3·2 | 97 | 57·2 | 47·2-67·2 |  |
|  | *Missing* | *0* | *0·0* |  | *0* | *0·0* |  | *0* | *0·0* |  | *0* | *0·0* |  |  |
| **Presence of health professionnals on site** | At least once per week | 1,01 | 27·4 | 22·2-32·6 | 157 | 18·8 | 10·2-27·3 | 827 | 30·9 | 24·3-37·4 | 26 | 15·2 | 7·8-22·5 | p<0·001*** |
|  | Less than once per week | 2,524 | 68·5 | 63·2-73·7 | 670 | 80·1 | 71·5-88·6 | 1,804 | 67·3 | 60·7-73·9 | 51 | 29·9 | 20·8-39·1 |  |
|  | Unknown/Not applicable | 153 | 4·1 | 2·9-5·3 | 10 | 1·2 | 0·3-2 | 49 | 1·8 | 0·5-3·2 | 93 | 54·9 | 44·9-65 |  |
|  | *Missing* | *0* | *0·0* |  | *0* | *0·0* |  | *0* | *0·0* |  | *0* | *0·0* |  |  |
| **Proximity of vaccination site targeting PEH (>15 or <15 min by foot)** | Close | 1,379 | 37·4 | 31·8-43 | 379 | 45·3 | 34·5-56·2 | 938 | 35·0 | 28·2-41·8 | 61 | 36·0 | 26·2-45·7 | 0·136· |
|  | Far | 2,308 | 62·6 | 57-68·2 | 457 | 54·7 | 43·8-65·5 | 1,742 | 65·0 | 58·2-71·8 | 109 | 64·0 | 54·3-73·8 |  |
|  | *Missing* | *0* | *0·0* |  | *0* | *0·0* |  | *0* | *0·0* |  | *0* | *0·0* |  |  |
| **Proximity of a vaccination center (>15 or <15 min by foot)** | Close | 1,814 | 49·2 | 43·4-55 | 449 | 53·6 | 42·7-64·5 | 1,308 | 48·8 | 41·7-55·9 | 57 | 33·6 | 24-43·1 | 0·188· |
|  | Far | 1,873 | 50·8 | 45-56·6 | 388 | 46·4 | 35·5-57·3 | 1,372 | 51·2 | 44·1-58·3 | 113 | 66·4 | 56·9-76 |  |
|  | *Missing* | *0* | *0·0* |  | *0* | *0·0* |  | *0* | *0·0* |  | *0* | *0·0* |  |  |
| **Proximity of a GP (>15 or <15 min by foot)** | Close | 3,223 | 87·4 | 83·6-91·2 | 784 | 93·7 | 88·8-98·6 | 2,302 | 85·9 | 80·9-90·9 | 136 | 80·3 | 72·6-88 | 0·04* |
|  | Far | 464 | 12·6 | 8·8-16·4 | 52 | 6·3 | 1·4-11·2 | 378 | 14·1 | 9·1-19·1 | 34 | 19·7 | 12-27·4 |  |
|  | *Missing* | *0* | *0·0* |  | *0* | *0·0* |  | *0* | *0·0* |  | *0* | *0·0* |  |  |
| **Proximity of a drugstore (>15 or <15 min by foot)** | Close | 3,287 | 89·2 | 85·6-92·8 | 793 | 94·8 | 89·2-100·4 | 2,346 | 87·5 | 82·9-92·2 | 148 | 86·9 | 80·5-93·4 | 0·131· |
|  | Far | 400 | 10·8 | 7·2-14·4 | 44 | 5·2 | -0·4-10·8 | 334 | 12·5 | 7·8-17·1 | 22 | 13·1 | 6·6-19·5 |  |
|  | *Missing* | *0* | *0·0* |  | *0* | *0·0* |  | *0* | *0·0* |  | *0* | *0·0* |  |  |
| **Proximity of a mobile clinic (targeting PEH)** | Close | 1,007 | 27·3 | 22·5-32·2 | 408 | 48·8 | 37·9-59·8 | 519 | 19·4 | 13·9-24·8 | 80 | 47·1 | 37-57·2 | p<0·001*** |
|  | Far | 2,68 | 72·7 | 67·8-77·5 | 428 | 51·2 | 40·2-62·1 | 2,162 | 80·6 | 75·2-86·1 | 90 | 52·9 | 42·8-63 |  |
|  | *Missing* | *0* | *0·0* |  | *0* | *0·0* |  | *0* | *0·0* |  | *0* | *0·0* |  |  |
| **Intervention on site - Awareness/sensitization** | No | 319 | 8·6 | 6·3-11 | 10 | 1·2 | 0·3-2 | 183 | 6·8 | 3·7-9·9 | 126 | 74·2 | 65·4-83 | p<0·001*** |
|  | Yes | 3,368 | 91·4 | 89-93·7 | 827 | 98·8 | 98-99·7 | 2,498 | 93·2 | 90·1-96·3 | 44 | 25·8 | 17-34·6 |  |
|  | *Missing* | *0* | *0·0* |  | *0* | *0·0* |  | *0* | *0·0* |  | *0* | *0·0* |  |  |
| **Intervention on site - Personalized support/mobilization** | No | 2,009 | 54·5 | 48·7-60·3 | 420 | 50·2 | 39·3-61·2 | 1,441 | 53·8 | 46·6-60·9 | 148 | 87·1 | 80·7-93·5 | 0·008** |
|  | Yes | 1,678 | 45·5 | 39·7-51·3 | 417 | 49·8 | 38·8-60·7 | 1,239 | 46·2 | 39·1-53·4 | 22 | 12·9 | 6·5-19·3 |  |
|  | *Missing* | *0* | *0·0* |  | *0* | *0·0* |  | *0* | *0·0* |  | *0* | *0·0* |  |  |
| **Intervention on site - Actual vaccination activity** | No | 2,023 | 54·9 | 49·1-60·6 | 329 | 39·3 | 28·5-50·1 | 1,546 | 57·7 | 50·6-64·8 | 147 | 86·7 | 79·5-93·9 | p<0·001*** |
|  | Yes | 1,664 | 45·1 | 39·4-50·9 | 508 | 60·7 | 49·9-71·5 | 1,134 | 42·3 | 35·2-49·4 | 23 | 13·3 | 6·1-20·5 |  |
|  | *Missing* | *0* | *0·0* |  | *0* | *0·0* |  | *0* | *0·0* |  | *0* | *0·0* |  |  |

#### Appendix 9. Table S2. Univariate Analysis

|  | | **Weighted data for sample design** | | | **Crude data** | | | |
| --- | --- | --- | --- | --- | --- | --- | --- | --- |
| **Variable** | **Categories** | **Vaccinated/Total** | **%** | **IC95 %** | **OR** | **IC95 %** | **Mod· p-value** | **Wald p-value** |
| **Socio-demographic features** | | | | | | | | |
| **Gender** | Male | 1571/1979 | 79·38 | 77·2-81·6 |  |  | Ref· | <0·001*** |
|  | Female | 1233/1703 | 72·39 | 69·5-75·3 | 0·7 | 0·6-0·8 | <0·001*** |  |
| **Age categories** | Less than 35 years | 1017/1479 | 68·77 | 65·8-71·8 |  |  | Ref· | <0·001*** |
|  | From 35 to 65 years | 1503/1890 | 79·52 | 77·4-81·6 | 1·6 | 1·4-1·9 | <0·001*** |  |
|  | More than 65 years | 285/311 | 91·61 | 88·1-95·1 | 4·8 | 3·2-7·2 | <0·001*** |  |
| **Region of Birth** | France | 204/284 | 71·87 | 64·9-78·8 |  |  | Ref· | <0·001*** |
|  | European Union (EU) | 55/153 | 36·10 | 25-47·2 | 0·2 | 0·1-0·3 | <0·001*** |  |
|  | Europe (outside of EU) | 35/64 | 54·62 | 40·9-68·3 | 0·5 | 0·3-0·9 | 0·019* |  |
|  | Maghreb (North Africa) | 464/596 | 77·80 | 73·5-82·2 | 1·6 | 1·2-2·2 | 0·003** |  |
|  | Central Africa/Horn of Africai/Southern Africa | 466/630 | 73·91 | 69·4-78·4 | 1·4 | 1·1-1·9 | 0·023* |  |
|  | West Africa | 1277/1579 | 80·89 | 78·3-83·5 | 2·2 | 1·6-2·9 | <0·001*** |  |
|  | Middle East and Central Asia | 179/231 | 77·29 | 69·9-84·7 | 1·8 | 1·2-2·8 | 0·009** |  |
|  | Other (Americas, Eastern Asia) | 114/133 | 85·95 | 78·9-93 | 3·2 | 1·9-5·5 | <0·001*** |  |
| **Length of stay in France** | Since COVID-19 pandemic start | 281/392 | 71·68 | 66·5-76·8 |  |  | Ref· | 0·073· |
|  | Less than 10 years, before COVID-19 | 1515/1997 | 75·84 | 73·1-78·6 | 1·3 | 1-1·7 | 0·029* |  |
|  | More than 10 years (or born in France) | 998/1276 | 78·21 | 75·4-81 | 1·3 | 1-1·7 | 0·037* |  |
| **Level of education** | Never attended school / Illiterate | 655/802 | 81·67 | 78·5-84·9 |  |  | Ref· | 0·002** |
|  | Primary and/or literate | 521/675 | 77·13 | 73·5-80·8 | 0·7 | 0·6-0·9 | 0·016* |  |
|  | Secondary | 1146/1580 | 72·52 | 69·4-75·6 | 0·7 | 0·5-0·8 | <0·001*** |  |
|  | Higher (university) | 487/630 | 77·32 | 73-81·6 | 0·8 | 0·6-1·1 | 0·17· |  |
| **Francophone** | Yes | 2194/2849 | 77·02 | 75-79·1 |  |  | Ref· | <0·001*** |
|  | No | 614/838 | 73·28 | 69·5-77 | 0·7 | 0·5-0·8 | <0·001*** |  |
| **Administrative status** | French or European nationality (EU zone) | 358/555 | 64·51 | 59·1-70 |  |  | Ref· | <0·001*** |
|  | Asylum seeker / Ongoing application | 1125/1369 | 82·16 | 79·4-84·9 | 3·5 | 2·7-4·5 | <0·001*** |  |
|  | Residence permit / Refugee status | 432/562 | 76·79 | 72·5-81·1 | 2·4 | 1·7-3·2 | <0·001*** |  |
|  | Undocumented | 890/1196 | 74·44 | 71·2-77·6 | 2·1 | 1·7-2·8 | <0·001*** |  |
| **Vaccination / Opinion on vaccination** | | | | | | | | |
| **Vaccinated in childhood (any antigen)** | Yes | 2553/3328 | 76·72 | 74·7-78·7 |  |  | Ref· | 0·01* |
|  | No | 182/263 | 69·26 | 62·8-75·7 | 0·7 | 0·5-0·9 | 0·01* |  |
| **Opinion on vaccination (in general)** | Positive | 2576/3292 | 78·25 | 76·4-80·1 |  |  | Ref· | <0·001*** |
|  | Negative | 233/392 | 59·46 | 53·4-65·6 | 0·4 | 0·3-0·5 | <0·001*** |  |
| **Opinion on Covid-19 vaccination** | Positive | 2315/2556 | 90·57 | 89·1-92 |  |  | Ref· | <0·001*** |
|  | Negative | 357/893 | 39·99 | 35·5-44·4 | 0·1 | 0-0·1 | <0·001*** |  |
|  | No opinion | 129/224 | 57·64 | 50·1-65·1 | 0·1 | 0·1-0·2 | <0·001*** |  |
| **Perceived utility of Covid-19 vaccination** | Yes | 2043/2345 | 87·14 | 85·4-88·9 |  |  | Ref· | <0·001*** |
|  | No | 511/971 | 52·64 | 48·2-57·1 | 0·1 | 0·1-0·2 | <0·001*** |  |
|  | No opinion | 245/357 | 68·58 | 63-74·2 | 0·3 | 0·2-0·4 | <0·001*** |  |
| **Fear of Covid-19 vaccination** | No | 1636/1972 | 82·96 | 80·8-85·2 |  |  | Ref· | <0·001*** |
|  | Yes | 1159/1691 | 68·55 | 65·7-71·4 | 0·4 | 0·3-0·5 | <0·001*** |  |
| **Family and friends' opinion on Covid-19 vaccination** | Mixed positive and negative / Unknown | 1031/1388 | 74·30 | 71·4-77·2 |  |  | Ref· | <0·001*** |
|  | Overall positive | 1309/1558 | 84·04 | 81·8-86·3 | 1·8 | 1·5-2·1 | <0·001*** |  |
|  | Overall negative | 469/742 | 63·17 | 58·5-67·8 | 0·4 | 0·3-0·5 | <0·001*** |  |
| **Needs/uses vaccine certificate in daily life** | Yes | 2375/2962 | 80·18 | 78·2-82·2 |  |  | Ref· | <0·001*** |
|  | No | 426/714 | 59·62 | 55·4-63·9 | 0·3 | 0·3-0·4 | <0·001*** |  |
| **Place of living** | | | | | | | | |
| **Stratum** | Housing | 716/837 | 85·57 | 83-88·2 |  |  | Ref· | <0·001*** |
|  | Accomodation | 2021/2680 | 75·40 | 73-77·8 | 0·5 | 0·4-0·7 | <0·001*** |  |
|  | Homeless (Streets) | 71/170 | 42·02 | 34·3-49·7 | 0·1 | 0·1-0·2 | <0·001*** |  |
| **Main living place the last 3 months** | Rents his/her own flat/live with family | 25/37 | 67·24 | 42·9-91·6 |  |  | Ref· | <0·001*** |
|  | Shelters (emergency, long term etc) | 1143/1430 | 79·91 | 76·6-83·2 | 1·5 | 0·8-3 | 0·25 |  |
|  | Social hostel | 878/1250 | 70·25 | 67-73·5 | 1·0 | 0·5-1·9 | 0·903 |  |
|  | Migrant Workers House | 691/800 | 86·42 | 83·8-89 | 2·2 | 1·1-4·4 | 0·027* |  |
|  | Travellers campsite | 0/3 | 5·00 | -7·1-17 | 0·2 | 0-2·4 | 0·202 |  |
|  | Street (roof: tent, parking, slum) | 12/37 | 32·51 | 19-46 | 0·2 | 0·1-0·5 | <0·001*** |  |
|  | Street (no roof) | 59/130 | 45·56 | 36·2-54·9 | 0·4 | 0·2-0·7 | 0·004** |  |
| **Ethos Typology** | 0· Rents his/her own flat | 26/41 | 63·98 | 39·1-88·9 |  |  | Ref· | <0·001*** |
|  | 1· People Living Rough (Public space or external space, Living in the streets or public spaces, without a shelter that can be defined as living quarters) | 53/105 | 50·56 | 39·6-61·6 | 0·5 | 0·3-1 | 0·046* |  |
|  | 11· People living in temporary/non-conventional structures (Semi-permanent structure hut or cabin) | 9/30 | 29·92 | 16·7-43·1 | 0·2 | 0·1-0·5 | <0·001*** |  |
|  | 12· People living in unfit housing (Defined as unfit for habitation by national legislation or building regulations) | 0/2 | 20·40 | -17·4-58·2 | 1·7 | 0·2-17·2 | 0·638 |  |
|  | 2· People in emergency accommodation (People with no usual place of residence who make use of overnight shelter, low threshold shelter) | 1/3 | 43·15 | -19·6-105·9 | 0·9 | 0·1-8·6 | 0·902 |  |
|  | 3· People in accommodation for the homeless (Homeless hostels, temporary accomodation, normally short-term) | 346/461 | 75·05 | 68·6-81·5 | 1·2 | 0·6-2·5 | 0·598 |  |
|  | 5· People in accommodation for immigrants (Immigrants in reception or short term accommodation due to their immigrant status) | 1451/1727 | 84·00 | 81·6-86·4 | 2·2 | 1·1-4·2 | 0·02* |  |
|  | 7· Supported accommodation for formerly homeless people(Stay longer than needed due to lack of housing) | 850/1207 | 70·46 | 67·2-73·7 | 1·0 | 0·5-2 | 0·892 |  |
|  | 8· People living in insecure accommodation (Occupation of land with no legal rights, squats) | 37/72 | 51·71 | 40·1-63·3 | 0·4 | 0·2-0·9 | 0·025* |  |
|  | Rents a flat / hosted by family | 26/41 | 63·98 | 39·1-88·9 |  |  | Ref· | <0·001*** |
|  | Rooflessness (without a shelter of any kind, sleeping rough) | 54/107 | 50·35 | 39·5-61·2 | 0·5 | 0·3-1 | 0·05· |  |
|  | Houselessness (with a place to sleep but temporary/emergency in institutions or shelter ) | 2647/3395 | 77·97 | 76-79·9 | 1·5 | 0·8-2·8 | 0·247 |  |
|  | Living in insecure housing (can be expelled easily, poor living conditions etc) | 37/72 | 51·71 | 40·1-63·3 | 0·4 | 0·2-0·9 | 0·025* |  |
|  | Living in inadequate housing (in caravans on illegal campsites, in unfit housing, in extreme overcrowding) | 10/33 | 29·24 | 16·5-42 | 0·2 | 0·1-0·5 | <0·001*** |  |
|  | Rents his/her own flat | 26/41 | 63·98 | 39·1-88·9 |  |  | Ref· | <0·001*** |
|  | Rooflessness (Living in the streets or public spaces without a shelter that can be defined as living quarters) | 60/120 | 50·18 | 40·3-60 | 0·5 | 0·2-0·9 | 0·031* |  |
|  | Emergency accommodation (Overnight shelters, People with no place of usual residence who move frequently between various types of accommodation) | 765/934 | 81·87 | 78·1-85·7 | 1·9 | 1-3·8 | 0·057· |  |
|  | People living in accommodation for the homeless /migrants (homeless hostels, centers for asylum seekers) | 1884/2464 | 76·44 | 74·2-78·7 | 1·4 | 0·7-2·6 | 0·343 |  |
|  | Inadequate Housing ( nonconventional dwellings, unfit housing, in extreme overcrowding) | 2/17 | 12·50 | 1·4-23·6 | 0·1 | 0-0·3 | <0·001*** |  |
|  | Hosted by a third-part/Squat | 37/72 | 51·71 | 40·1-63·3 | 0·4 | 0·2-0·9 | 0·025* |  |
| **Change in housing/location over the past three months** | One location only | 2544/3293 | 77·26 | 75·3-79·3 |  |  | Ref· | <0·001*** |
|  | Several locations | 251/376 | 66·76 | 61·1-72·4 | 0·6 | 0·5-0·7 | <0·001*** |  |
| **Household structure** | Live by him/herself | 983/1205 | 81·59 | 78·9-84·3 |  |  | Ref· | <0·001*** |
|  | Live with his/her partner and children | 606/879 | 68·89 | 64·9-72·9 | 0·5 | 0·4-0·7 | <0·001*** |  |
|  | Live with his/her children, without other adults | 486/679 | 71·55 | 66·9-76·2 | 0·7 | 0·6-0·9 | 0·017* |  |
|  | Live with his/her partner or related adults, without children | 301/413 | 72·81 | 67·3-78·4 | 0·7 | 0·5-0·9 | 0·006** |  |
|  | Live with other non-related adults | 422/497 | 84·93 | 80·7-89·2 | 1·3 | 1-1·7 | 0·094· |  |
| **Financial / food situation / Income** | | | | | | | | |
| **Source of Income** | Declared and official sources of income (salary, pensions, state aids…) | 1314/1642 | 80·00 | 77·2-82·8 |  |  | Ref· | <0·001*** |
|  | No income/informal activities/no regular pension or aid | 1478/2022 | 73·09 | 70·4-75·8 | 0·6 | 0·5-0·7 | <0·001*** |  |
| **Food security** | Food enough in quantity and quality | 1459/1879 | 77·65 | 75·1-80·2 |  |  | Ref· | 0·08· |
|  | Not enough food and not good enough | 1329/1773 | 74·97 | 72·5-77·4 | 0·9 | 0·7-1 | 0·08· |  |
| **Buys his/her own meals (most of the time)** | No | 838/1118 | 74·92 | 71·7-78·1 |  |  | Ref· | 0·419 |
|  | Yes | 1964/2560 | 76·71 | 74·5-78·9 | 1·1 | 0·9-1·3 | 0·419 |  |
| **Meals provided by relatives/family/friends** | No | 2519/3275 | 76·91 | 74·9-78·9 |  |  | Ref· | <0·001*** |
|  | Yes | 284/405 | 70·11 | 64·3-75·9 | 0·7 | 0·5-0·8 | <0·001*** |  |
| **Meals through food distributions** | No | 1756/2236 | 78·53 | 76·2-80·8 |  |  | Ref· | 0·079· |
|  | Yes | 1046/1443 | 72·50 | 69·5-75·5 | 0·9 | 0·7-1 | 0·079· |  |
| **Meals through panhandling** | No | 2708/3510 | 77·16 | 75·2-79·1 |  |  | Ref· | <0·001*** |
|  | Yes | 95/170 | 55·70 | 47·4-64 | 0·3 | 0·3-0·5 | <0·001*** |  |
| **Meals through dumpster rummaging** | No | 2788/3649 | 76·40 | 74·5-78·3 |  |  | Ref· | <0·001*** |
|  | Yes | 15/31 | 48·30 | 31·8-64·7 | 0·2 | 0·1-0·4 | <0·001*** |  |
| **Meals difficult to get/not enough in quantity** | No | 2647/3456 | 76·58 | 74·6-78·5 |  |  | Ref· | 0·01* |
|  | Yes | 156/223 | 69·76 | 62·1-77·4 | 0·7 | 0·5-0·9 | 0·01* |  |
| **Meals provided by site managers** | No | 2230/3008 | 74·14 | 72·1-76·2 |  |  | Ref· | <0·001*** |
|  | Yes | 572/671 | 85·24 | 81·3-89·2 | 2·1 | 1·6-2·7 | <0·001*** |  |
| **Source of income - declared employment** | No | 2205/2916 | 75·63 | 73·5-77·8 |  |  | Ref· | 0·015* |
|  | Yes | 597/764 | 78·08 | 74-82·2 | 1·3 | 1·1-1·6 | 0·015* |  |
| **Source of income - undeclared employment** | No | 2361/3085 | 76·52 | 74·5-78·5 |  |  | Ref· | 0·011* |
|  | Yes | 441/595 | 74·18 | 70-78·4 | 0·7 | 0·6-0·9 | 0·011* |  |
| **Source of income - retirement pension and/or alimony** | No | 2591/3451 | 75·08 | 73·1-77·1 |  |  | Ref· | <0·001*** |
|  | Yes | 211/229 | 92·09 | 88·2-95·9 | 3·5 | 2·2-5·5 | <0·001*** |  |
| **Source of income - unemployment benefit** | No | 2723/3577 | 76·13 | 74·2-78 |  |  | Ref· | 0·548 |
|  | Yes | 78/102 | 76·56 | 65·8-87·4 | 1·2 | 0·7-2 | 0·548 |  |
| **Source of income - scholarship** | No | 2786/3662 | 76·08 | 74·1-78 |  |  | Ref· | 0·098· |
|  | Yes | 16/18 | 87·63 | 71·8-103·5 | 3·3 | 0·8-13·4 | 0·098· |  |
| **Source of income - social solidarity benefit** | No | 2480/3253 | 76·24 | 74·2-78·3 |  |  | Ref· | 0·384 |
|  | Yes | 322/427 | 75·38 | 70·5-80·3 | 0·9 | 0·7-1·1 | 0·384 |  |
| **Source of income - other(s) benefit(s)** | No | 2248/2971 | 75·67 | 73·5-77·8 |  |  | Ref· | 0·03* |
|  | Yes | 554/709 | 78·10 | 74-82·2 | 1·3 | 1-1·6 | 0·03* |  |
| **Source of income - begging** | No | 2775/3610 | 76·87 | 74·9-78·8 |  |  | Ref· | <0·001*** |
|  | Yes | 27/70 | 38·35 | 25·8-50·9 | 0·2 | 0·1-0·3 | <0·001*** |  |
| **Source of income - associative/housing support** | No | 2409/3141 | 76·71 | 74·7-78·7 |  |  | Ref· | 0·794 |
|  | Yes | 393/539 | 72·83 | 67-78·7 | 1·0 | 0·8-1·3 | 0·794 |  |
| **Source of income - family/friend support** | No | 2626/3433 | 76·50 | 74·6-78·4 |  |  | Ref· | 0·023* |
|  | Yes | 175/246 | 71·08 | 62·9-79·2 | 0·7 | 0·5-0·9 | 0·023* |  |
| **Source of income - none** | No | 2186/2838 | 77·04 | 74·8-79·2 |  |  | Ref· | 0·151· |
|  | Yes | 616/843 | 73·12 | 69·7-76·6 | 0·9 | 0·7-1·1 | 0·151· |  |
| **Source of income - unknown** | No | 2788/3660 | 76·17 | 74·2-78·1 |  |  | Ref· | 0·268 |
|  | Yes | 14/20 | 70·53 | 47·2-93·8 | 0·6 | 0·2-1·5 | 0·268 |  |
| **Support and coping systems** | | | | | | | | |
| **Attends food distributions on a regular basis** | No | 1759/2246 | 78·33 | 76-80·7 |  |  | Ref· | 0·546 |
|  | Yes | 1024/1407 | 72·75 | 69·5-76 | 0·9 | 0·8-1·1 | 0·546 |  |
| **Benefits from outreach activities** | No | 2569/3335 | 77·03 | 75-79·1 |  |  | Ref· | 0·005** |
|  | Yes | 214/318 | 67·22 | 61·3-73·2 | 0·7 | 0·6-0·9 | 0·005** |  |
| **Supported by site managers/social workers on site** | No | 1834/2433 | 75·36 | 72·9-77·8 |  |  | Ref· | <0·001*** |
|  | Yes | 949/1220 | 77·81 | 74·6-81 | 1·4 | 1·2-1·7 | 0·001** |  |
| **Supported by social workers (external)** | No | 2124/2791 | 76·11 | 73·9-78·3 |  |  | Ref· | 0·502 |
|  | Yes | 659/863 | 76·40 | 72·4-80·4 | 1·1 | 0·9-1·3 | 0·502 |  |
| **Supported by state/city services** | No | 2730/3588 | 76·08 | 74·1-78 |  |  | Ref· | 0·358 |
|  | Yes | 53/65 | 81·75 | 70·8-92·7 | 1·3 | 0·7-2·5 | 0·358 |  |
| **Supported by health professionals** | No | 2595/3423 | 75·80 | 73·8-77·8 |  |  | Ref· | 0·238 |
|  | Yes | 188/230 | 81·78 | 75-88·5 | 1·2 | 0·9-1·8 | 0·238 |  |
| **Supported by religieux groups/community** | No | 2655/3470 | 76·50 | 74·6-78·4 |  |  | Ref· | 0·773 |
|  | Yes | 128/183 | 69·98 | 60·3-79·7 | 0·9 | 0·7-1·4 | 0·773 |  |
| **Autonomous / Does not receive or need support whatsoever** | No | 1943/2593 | 74·92 | 72·6-77·3 |  |  | Ref· | 0·665 |
|  | Yes | 840/1060 | 79·25 | 76·1-82·4 | 1·0 | 0·8-1·2 | 0·665 |  |
| **Benefits from support to attend and navigate the healthcare system** | Yes (relatives, social workers etc) | 813/1074 | 75·67 | 72·6-78·8 |  |  | Ref· | 0·002** |
|  | I need help but I go by myself | 1038/1391 | 74·64 | 71·5-77·8 | 1·1 | 0·9-1·3 | 0·576 |  |
|  | Autonomous/independant | 896/1122 | 79·90 | 76·7-83·1 | 1·3 | 1-1·6 | 0·023* |  |
|  | I never consult/It never happened so far | 39/68 | 57·54 | 46·9-68·2 | 0·6 | 0·4-0·9 | 0·012* |  |
| **Moral and social support** | | | | | | | | |
| **Support from relatives (can speak to/be listened by relatives)** | Yes, often | 1021/1335 | 76·45 | 73·5-79·4 |  |  | Ref· | 0·546 |
|  | Yes, sometimes | 973/1287 | 75·59 | 72·3-78·9 | 1·0 | 0·8-1·3 | 0·722 |  |
|  | No, almost never | 722/942 | 76·63 | 73·4-79·9 | 1·0 | 0·8-1·3 | 0·703 |  |
|  | NA: situation never occurred | 67/88 | 76·65 | 67·1-86·2 | 0·8 | 0·5-1·2 | 0·218 |  |
| **Material support from relatives (money, food, clothes etc)** | Yes, often | 357/479 | 74·61 | 69·6-79·6 |  |  | Ref· | 0·025* |
|  | Yes, sometimes | 789/1032 | 76·49 | 73·4-79·6 | 1·3 | 1-1·7 | 0·025* |  |
|  | No, almost never | 1452/1927 | 75·36 | 72·9-77·9 | 1·4 | 1·1-1·9 | 0·006** |  |
|  | NA: situation never occurred | 175/202 | 86·68 | 80·5-92·8 | 1·8 | 1·1-2·8 | 0·013* |  |
| **Advices from relatives (to find a job etc)** | Yes, often | 667/857 | 77·81 | 74·3-81·4 |  |  | Ref· | 0·85 |
|  | Yes, sometimes | 1097/1443 | 76·04 | 72·9-79·2 | 1·0 | 0·8-1·2 | 0·969 |  |
|  | No, almost never | 906/1202 | 75·35 | 72·4-78·3 | 1·0 | 0·8-1·2 | 0·81 |  |
|  | NA: situation never occurred | 99/126 | 78·50 | 70·2-86·8 | 0·8 | 0·5-1·3 | 0·407 |  |
| **Moral support by relatives (motivation)** | Yes, often | 726/938 | 77·37 | 74-80·8 |  |  | Ref· | 0·837 |
|  | Yes, sometimes | 1081/1427 | 75·72 | 72·6-78·8 | 1·0 | 0·8-1·3 | 0·921 |  |
|  | No, almost never | 797/1057 | 75·38 | 72·2-78·6 | 0·9 | 0·7-1·2 | 0·516 |  |
|  | NA: situation never occurred | 146/188 | 77·57 | 70·1-85 | 1·0 | 0·6-1·4 | 0·852 |  |
| **Support from organization members (can speak to/be listened by members)** | Yes, often | 396/521 | 76·04 | 71·5-80·6 |  |  | Ref· | 0·021* |
|  | Yes, sometimes | 931/1195 | 77·92 | 74·7-81·2 | 1·3 | 1-1·6 | 0·085· |  |
|  | No, almost never | 1203/1622 | 74·18 | 71·4-77 | 0·9 | 0·7-1·2 | 0·634 |  |
|  | NA: situation never occurred | 243/305 | 79·74 | 74·5-85 | 0·9 | 0·7-1·4 | 0·769 |  |
| **Material support from org· members (money, food, clothes etc)** | Yes, often | 412/555 | 74·25 | 69·2-79·3 |  |  | Ref· | 0·406 |
|  | Yes, sometimes | 865/1132 | 76·44 | 73-79·9 | 1·1 | 0·9-1·5 | 0·408 |  |
|  | No, almost never | 1252/1653 | 75·73 | 72·7-78·7 | 0·9 | 0·7-1·2 | 0·682 |  |
|  | NA: situation never occurred | 243/298 | 81·45 | 76·3-86·6 | 0·9 | 0·6-1·4 | 0·647 |  |
| **Advices from org· members (to find a job etc)** | Yes, often | 391/516 | 75·78 | 71·6-80 |  |  | Ref· | 0·099· |
|  | Yes, sometimes | 1001/1291 | 77·53 | 74·2-80·8 | 1·1 | 0·9-1·4 | 0·48 |  |
|  | No, almost never | 1165/1557 | 74·82 | 72·1-77·6 | 0·9 | 0·7-1·1 | 0·263 |  |
|  | NA: situation never occurred | 205/262 | 78·35 | 72·5-84·2 | 0·8 | 0·6-1·1 | 0·199· |  |
| **Moral support by org· members (motivation)** | Yes, often | 299/378 | 79·21 | 74·2-84·2 |  |  | Ref· | 0·02* |
|  | Yes, sometimes | 829/1075 | 77·11 | 73·8-80·4 | 1·1 | 0·8-1·4 | 0·668 |  |
|  | No, almost never | 1360/1820 | 74·73 | 72-77·5 | 0·8 | 0·6-1·1 | 0·112· |  |
|  | NA: situation never occurred | 259/333 | 77·87 | 72·4-83·3 | 0·8 | 0·6-1·2 | 0·272 |  |
| **Self-reported feeling of loneliness** | I feel very lonely | 430/570 | 75·41 | 71·5-79·4 |  |  | Ref· | 0·337 |
|  | I don't feel taken care of/supported· I feel lonely sometimes· | 912/1194 | 76·37 | 73·5-79·3 | 1·2 | 1-1·5 | 0·08· |  |
|  | I feel taken care of/supported/not specially lonely | 1058/1375 | 76·94 | 74-79·9 | 1·2 | 0·9-1·5 | 0·209 |  |
|  | I feel very taken care of/supported· I don't feel lonely at all | 354/467 | 75·73 | 70·5-80·9 | 1·1 | 0·8-1·4 | 0·604 |  |
| **COVID-19 (vaccination) Information Sources** | | | | | | | | |
| **Internet connection** | Yes, seldom or often uses internet | 2155/2857 | 75·44 | 73·3-77·6 |  |  | Ref· | 0·347 |
|  | No, never uses internet | 628/794 | 79·14 | 75·7-82·5 | 0·9 | 0·8-1·1 | 0·508 |  |
|  | Other: another mean of connection | 4/8 | 50·95 | 17·6-84·3 | 0·5 | 0·1-1·5 | 0·191· |  |
| **Compliance to protection measures (wear a facemask in public transportation etc)** | Yes, I wear it and I think it's useful | 2235/2882 | 77·56 | 75·4-79·7 |  |  | Ref· | <0·001*** |
|  | It depends on the context | 196/266 | 73·57 | 67·2-79·9 | 0·8 | 0·6-1 | 0·087· |  |
|  | No, this is useless | 284/407 | 69·68 | 64·1-75·3 | 0·6 | 0·5-0·8 | <0·001*** |  |
| **COVID-19 (Vaccine) information source - internet / social media** | No | 1867/2342 | 79·73 | 77·5-82 |  |  | Ref· | <0·001*** |
|  | Yes | 941/1345 | 69·98 | 66·8-73·1 | 0·6 | 0·5-0·7 | <0·001*** |  |
| **COVID-19 (Vaccine) information source - television/radio** | No | 880/1171 | 75·12 | 71·8-78·4 |  |  | Ref· | 0·002** |
|  | Yes | 1922/2507 | 76·65 | 74·6-78·7 | 1·3 | 1·1-1·5 | 0·002** |  |
| **COVID-19 (Vaccine) information source - print media/classic media** | No | 2647/3460 | 76·49 | 74·5-78·5 |  |  | Ref· | 0·037* |
|  | Yes | 155/218 | 70·98 | 64-77·9 | 0·8 | 0·6-1 | 0·037* |  |
| **COVID-19 (Vaccine) information source - posters/leaflets** | No | 2652/3485 | 76·10 | 74·1-78·1 |  |  | Ref· | 0·154· |
|  | Yes | 150/194 | 77·30 | 71·1-83·5 | 1·3 | 0·9-1·8 | 0·154· |  |
| **COVID-19 (Vaccine) information source - site manager** | No | 2628/3483 | 75·46 | 73·5-77·4 |  |  | Ref· | <0·001*** |
|  | Yes | 173/195 | 88·76 | 83·7-93·8 | 2·4 | 1·6-3·8 | <0·001*** |  |
| **COVID-19 (Vaccine) information source - health or social workers/professionnals** | No | 2197/2953 | 74·40 | 72·3-76·5 |  |  | Ref· | <0·001*** |
|  | Yes | 612/734 | 83·32 | 79·5-87·2 | 1·6 | 1·3-2 | <0·001*** |  |
| **COVID-19 (Vaccine) information source - relatives/family/community** | No | 1980/2572 | 76·99 | 74·7-79·2 |  |  | Ref· | <0·001*** |
|  | Yes | 828/1115 | 74·29 | 71·2-77·4 | 0·7 | 0·5-0·8 | <0·001*** |  |
| **COVID-19 (Vaccine) information source - none** | No | 2754/3607 | 76·35 | 74·4-78·3 |  |  | Ref· | 0·013* |
|  | Yes | 47/71 | 66·58 | 53·6-79·6 | 0·6 | 0·4-0·9 | 0·013* |  |
| **Satisfaction with COVID-19 information (overall)** | No | 1176/1712 | 68·68 | 65·7-71·7 |  |  | Ref· | <0·001*** |
|  | Yes | 1500/1790 | 83·79 | 81·6-86 | 2·3 | 1·9-2·7 | <0·001*** |  |
|  | DNK/DNA | 133/185 | 71·81 | 64·4-79·2 | 1·2 | 0·9-1·6 | 0·339 |  |
| **Trust in the crisis management by the authorities** | Zero trust (0) | 115/219 | 52·51 | 44·6-60·4 |  |  | Ref· | <0·001*** |
|  | Low trust (1-3) | 171/258 | 66·28 | 59·4-73·2 | 1·8 | 1·3-2·6 | 0·001** |  |
|  | Medium trust (4-6) | 621/848 | 73·20 | 69·5-77 | 3·0 | 2·1-4·1 | <0·001*** |  |
|  | High trust (7-9) | 949/1142 | 83·10 | 80·3-85·9 | 5·2 | 3·8-7·1 | <0·001*** |  |
|  | Total trust (10) | 603/748 | 80·57 | 76·5-84·7 | 4·7 | 3·3-6·8 | <0·001*** |  |
|  | No opinion | 336/455 | 73·78 | 68·7-78·9 | 3·5 | 2·5-5 | <0·001*** |  |
| **Health-related information** | | | | | | | | |
| **Has medical coverage** | Yes (national social security, state medical aid) | 2493/3152 | 79·10 | 77·2-81 |  |  | Ref· | <0·001*** |
|  | No (never, lost or ongoing application) | 296/510 | 58·04 | 53·1-63 | 0·4 | 0·3-0·4 | <0·001*** |  |
| **Self-reported health status** | Bad status overall | 597/761 | 78·42 | 74·8-82 |  |  | Ref· | 0·005** |
|  | Good status overall | 1356/1760 | 77·06 | 74·6-79·5 | 1·0 | 0·8-1·2 | 0·982 |  |
|  | Perfect condition | 808/1107 | 73·02 | 69·6-76·4 | 0·8 | 0·6-0·9 | 0·008** |  |
| **Chronic condition (self-reported)** | No | 1893/2561 | 73·92 | 71·6-76·2 |  |  | Ref· | <0·001*** |
|  | Yes | 886/1087 | 81·54 | 78·2-84·8 | 1·4 | 1·2-1·7 | <0·001*** |  |
| **Is followed by a regular GP** | Yes | 1984/2465 | 80·50 | 78·5-82·6 |  |  | Ref· | <0·001*** |
|  | No | 804/1193 | 67·35 | 64·1-70·6 | 0·5 | 0·4-0·5 | <0·001*** |  |
| **Date of last consultation (hospital/GP)** | After the start of vaccination campaign (>may 2021) | 2036/2595 | 78·45 | 76·2-80·7 |  |  | Ref· | <0·001*** |
|  | Between 2d lockdown and start of vaccination (dec 2020-may 2021) | 261/334 | 78·03 | 73·4-82·6 | 0·8 | 0·6-1 | 0·067· |  |
|  | Between 1st and 2d lockdown (mar-dec 2020) | 278/387 | 71·91 | 66·7-77·1 | 0·8 | 0·6-1 | 0·079· |  |
|  | Before the pandemic (<march 2020) | 131/203 | 64·71 | 56·7-72·7 | 0·4 | 0·3-0·5 | <0·001*** |  |
|  | Never consulted in France/Does not apply | 69/123 | 55·82 | 46·2-65·4 | 0·4 | 0·3-0·5 | <0·001*** |  |
| **Previous history/hospitalization of COVID-19** | No | 2276/2991 | 76·10 | 74-78·2 |  |  | Ref· | 0·001** |
|  | Yes, without hospitalization | 439/588 | 74·61 | 70·1-79·1 | 1·0 | 0·8-1·2 | 0·741 |  |
|  | Yes, with hospitalization | 86/96 | 89·92 | 82·5-97·4 | 3·8 | 1·8-8 | <0·001*** |  |
| **Previous history/hospitalization for COVID-19 among family or friends** | No | 1530/1988 | 76·95 | 74·5-79·4 |  |  | Ref· | 0·327 |
|  | Yes, without hospitalization | 708/959 | 73·85 | 70·3-77·4 | 1·0 | 0·8-1·2 | 0·605 |  |
|  | Yes, with hospitalization | 571/741 | 77·10 | 73·2-81 | 1·1 | 0·9-1·4 | 0·252 |  |
| **Vulnerable/at risk for COVID-19 people among relatives** | No | 1839/2420 | 76·00 | 73·7-78·3 |  |  | Ref· | 0·081· |
|  | Yes | 964/1260 | 76·49 | 73·2-79·8 | 0·9 | 0·7-1 | 0·081· |  |
| **Health literacy** | | | | | | | | |
| **Can read and understand drug notices** | With ease overall, alone | 1536/2037 | 75·39 | 72·8-77·9 |  |  | Ref· | 0·771 |
|  | With difficulty overall, alone | 1247/1611 | 77·42 | 74·9-79·9 | 1·0 | 0·9-1·2 | 0·771 |  |
| **Can understand oral medical instructions** | With ease overall, alone | 2196/2860 | 76·77 | 74·7-78·9 |  |  | Ref· | 0·001** |
|  | With difficulty overall, alone | 587/791 | 74·17 | 70·6-77·8 | 0·7 | 0·6-0·9 | 0·001** |  |
| **Can understand oral medical advice** | With ease overall, alone | 2031/2663 | 76·26 | 74·1-78·5 |  |  | Ref· | 0·011* |
|  | With difficulty overall, alone | 751/987 | 76·07 | 73-79·1 | 0·8 | 0·6-0·9 | 0·011* |  |
| **Can read and understand written medical instructions** | With ease overall, alone | 1628/2151 | 75·68 | 73·2-78·1 |  |  | Ref· | 0·92 |
|  | With difficulty overall, alone | 1155/1497 | 77·17 | 74·6-79·8 | 1·0 | 0·8-1·2 | 0·92 |  |
| **Can correctly fill out a medical form** | With ease overall, alone | 1218/1650 | 73·84 | 71·1-76·6 |  |  | Ref· | 0·178· |
|  | With difficulty overall, alone | 1564/2001 | 78·17 | 75·9-80·5 | 1·1 | 0·9-1·3 | 0·178· |  |
| **Discrimination** | | | | | | | | |
| **Experienced discrimination since Covid-19 outbreak** | No | 2100/2713 | 77·40 | 75·2-79·6 |  |  | Ref· | <0·001*** |
|  | Yes | 404/590 | 68·48 | 64·2-72·8 | 0·6 | 0·5-0·7 | <0·001*** |  |
|  | Does not know/Does not want to answer | 304/383 | 79·33 | 74-84·6 | 1·0 | 0·8-1·3 | 0·834 |  |
| **Is more afraid of being expelled from France since COVID-19 crisis** | No | 923/1234 | 74·79 | 71·7-77·8 |  |  | Ref· | 0·341 |
|  | Yes | 207/284 | 72·94 | 66·4-79·4 | 0·9 | 0·6-1·2 | 0·341 |  |
| **Has been denied vaccination** | No | 2773/3637 | 76·23 | 74·3-78·2 |  |  | Ref· | 0·237 |
|  | Yes | 10/14 | 69·74 | 45·1-94·4 | 0·5 | 0·2-1·5 | 0·237 |  |
| **has been denied healthcare** | No | 2646/3446 | 76·79 | 74·8-78·8 |  |  | Ref· | <0·001*** |
|  | Yes | 129/196 | 65·64 | 58·2-73·1 | 0·6 | 0·4-0·7 | <0·001*** |  |
| **Recruitment site information** | | | | | | | | |
| **Vaccine certificate is mandatory to access site** | No | 2689/3524 | 76·31 | 74·4-78·3 |  |  | Ref· | 0·267 |
|  | Yes | 39/50 | 78·16 | 66-90·3 | 1·6 | 0·7-3·4 | 0·267 |  |
| **Presence of social workers on site** | At least once per week | 2153/2726 | 78·98 | 76·7-81·2 |  |  | Ref· | <0·001*** |
|  | Less than once per week | 594/825 | 71·97 | 68·1-75·9 | 0·5 | 0·4-0·7 | <0·001*** |  |
|  | Unknown/Not applicable | 61/135 | 45·30 | 37-53·6 | 0·2 | 0·2-0·4 | <0·001*** |  |
| **Presence of GP on site** | At least once per week | 295/364 | 81·02 | 76·2-85·9 |  |  | Ref· | <0·001*** |
|  | Less than once per week | 2414/3140 | 76·88 | 74·8-79 | 0·8 | 0·6-1·2 | 0·255 |  |
|  | Unknown/Not applicable | 99/182 | 54·37 | 44·3-64·5 | 0·3 | 0·2-0·4 | <0·001*** |  |
| **Presence of health professionnals on site** | At least once per week | 815/1010 | 80·66 | 77·3-84·1 |  |  | Ref· | <0·001*** |
|  | Less than once per week | 1923/2525 | 76·17 | 73·8-78·5 | 0·8 | 0·7-1 | 0·063· |  |
|  | Unknown/Not applicable | 71/153 | 46·56 | 38·9-54·2 | 0·3 | 0·2-0·4 | <0·001*** |  |
| **Proximity of vaccination site targeting PEH (>15 or <15 min by foot)** | Close | 1111/1379 | 80·56 | 77·7-83·4 |  |  | Ref· | <0·001*** |
|  | Far | 1698/2309 | 73·55 | 71-76·1 | 0·6 | 0·5-0·8 | <0·001*** |  |
| **Proximity of a vaccination center (>15 or <15 min by foot)** | Close | 1377/1814 | 75·89 | 73·3-78·5 |  |  | Ref· | 0·951 |
|  | Far | 1432/1873 | 76·44 | 73·6-79·3 | 1·0 | 0·8-1·2 | 0·951 |  |
| **Proximity of a GP (>15 or <15 min by foot)** | Close | 2447/3223 | 75·92 | 73·8-78 |  |  | Ref· | 0·768 |
|  | Far | 362/464 | 77·93 | 72·9-83 | 1·0 | 0·8-1·4 | 0·768 |  |
| **Proximity of a drugstore (>15 or <15 min by foot)** | Close | 2515/3287 | 76·52 | 74·5-78·5 |  |  | Ref· | 0·199· |
|  | Far | 293/400 | 73·31 | 66·9-79·8 | 0·8 | 0·6-1·1 | 0·199· |  |
| **Proximity of a mobile clinic (targeting PEH)** | Close | 803/1008 | 79·70 | 75·9-83·5 |  |  | Ref· | 0·09· |
|  | Far | 2006/2680 | 74·85 | 72·6-77·1 | 0·8 | 0·7-1 | 0·09· |  |
| **Intervention on site - Awareness/sensitization** | No | 165/319 | 51·67 | 45·6-57·7 |  |  | Ref· | <0·001*** |
|  | Yes | 2644/3368 | 78·49 | 76·5-80·4 | 3·7 | 2·9-4·7 | <0·001*** |  |
| **Intervention on site - Personalized support/mobilization** | No | 1455/2009 | 72·41 | 69·8-75·1 |  |  | Ref· | <0·001*** |
|  | Yes | 1353/1677 | 80·68 | 77·9-83·5 | 1·6 | 1·3-2 | <0·001*** |  |
| **Intervention on site - Actual vaccination activity** | No | 1435/2022 | 70·96 | 68·1-73·9 |  |  | Ref· | <0·001*** |
|  | Yes | 1373/1664 | 82·50 | 80·2-84·8 | 2·4 | 2-2·8 | <0·001*** |  |

#### Appendix 10. Table S3. Description of Site characteristics (Site Level)

|  | | **All stratum** | | | **Asylum seeker accommodation in Ile-de-France (IDF)** | | | **Social accommodations for homeless in IDF** | | | **Workers' residences in IDF** | | | **Slums, informal camps and in the streets in IDF** | | | **Homeless people in Marseille** | | |  |
| --- | --- | --- | --- | --- | --- | --- | --- | --- | --- | --- | --- | --- | --- | --- | --- | --- | --- | --- | --- | --- |
| **Variable** | **Categories** | **N** | **%** | **IC95 %** | **N** | **%** | **IC95 %** | **N** | **%** | **IC95 %** | **N** | **%** | **IC95 %** | **N** | **%** | **IC95 %** | **N** | **%** | **IC95 %** | **p-value** |
| **Recruitment site information** | | | | | | | | | | | | | | | | | | | | |
| **Vaccine certificate is mandatory to access site** | No | 192 | 98·5 | 96·7-100 | 43 | 100·0 | 100-100 | 43 | 95·6 | 89·4-100 | 32 | 100·0 | 100-100 | 49 | 98·0 | 94·1-100 | 25 | 100·0 | 100-100 | 0·129· |
|  | Yes | 3 | 1·5 | 0-3·3 |  |  |  | 2 | 4·4 | 0-10·6 |  |  |  | 1 | 2·0 | 0-5·9 |  |  |  |  |
|  | *Missing* | *14* | *6·7* |  | *0* | *0·0* |  | *0* | *0·0* |  | *0* | *0·0* |  | *12* | *19·4* |  | *2* | *7·4* |  |  |
| **Presence of social workers on site** | At least once per week | 111 | 53·1 | 47·7-58·6 | 42 | 97·7 | 93·1-100 | 20 | 44·4 | 29·7-59·2 | 27 | 84·4 | 71·5-97·2 | 13 | 21·0 | 10·7-31·2 | 9 | 33·3 | 15·1-51·6 | p<0·001*** |
|  | Less than once per week | 59 | 28·2 | 22·7-33·7 | 1 | 2·3 | 0-6·9 | 24 | 53·3 | 38·5-68·2 | 5 | 15·6 | 2·8-28·5 | 13 | 21·0 | 10·7-31·2 | 16 | 59·3 | 40·3-78·3 |  |
|  | Unknown/Not applicable | 39 | 18·7 | 14·6-22·7 |  |  |  | 1 | 2·2 | 0-6·6 |  |  |  | 36 | 58·1 | 45·6-70·5 | 2 | 7·4 | 0-17·5 |  |
|  | *Missing* | *0* | *0·0* |  | *0* | *0·0* |  | *0* | *0·0* |  | *0* | *0·0* |  | *0* | *0·0* |  | *0* | *0·0* |  |  |
| **Presence of GP on site** | At least once per week | 21 | 10·0 | 5·9-14·2 | 4 | 9·3 | 0·5-18·1 | 5 | 11·1 | 1·8-20·5 | 3 | 9·4 | 0-19·7 | 4 | 6·5 | 0·2-12·7 | 5 | 18·5 | 3·5-33·5 | p<0·001*** |
|  | Less than once per week | 139 | 66·5 | 61·5-71·5 | 39 | 90·7 | 81·9-99·5 | 39 | 86·7 | 76·6-96·8 | 28 | 87·5 | 75·8-99·2 | 13 | 21·0 | 10·7-31·2 | 20 | 74·1 | 57·1-91 |  |
|  | Unknown/Not applicable | 49 | 23·4 | 19·6-27·3 |  |  |  | 1 | 2·2 | 0-6·6 | 1 | 3·1 | 0-9·3 | 45 | 72·6 | 61·3-83·8 | 2 | 7·4 | 0-17·5 |  |
|  | *Missing* | *0* | *0·0* |  | *0* | *0·0* |  | *0* | *0·0* |  | *0* | *0·0* |  | *0* | *0·0* |  | *0* | *0·0* |  |  |
| **Presence of health professionnals on site** | At least once per week | 53 | 25·4 | 19·4-31·3 | 15 | 34·9 | 20·4-49·4 | 11 | 24·4 | 11·7-37·2 | 6 | 18·8 | 4·9-32·6 | 12 | 19·4 | 9·4-29·3 | 9 | 33·3 | 15·1-51·6 | p<0·001*** |
|  | Less than once per week | 109 | 52·2 | 46·5-57·8 | 28 | 65·1 | 50·6-79·6 | 33 | 73·3 | 60·2-86·5 | 26 | 81·2 | 67·4-95·1 | 6 | 9·7 | 2·2-17·1 | 16 | 59·3 | 40·3-78·3 |  |
|  | Unknown/Not applicable | 47 | 22·5 | 18·7-26·3 |  |  |  | 1 | 2·2 | 0-6·6 |  |  |  | 44 | 71·0 | 59·5-82·4 | 2 | 7·4 | 0-17·5 |  |
|  | *Missing* | *0* | *0·0* |  | *0* | *0·0* |  | *0* | *0·0* |  | *0* | *0·0* |  | *0* | *0·0* |  | *0* | *0·0* |  |  |
| **Proximity of vaccination site targeting PEH (>15 or <15 min by foot)** | Close | 71 | 34·0 | 27·7-40·3 | 19 | 44·2 | 29·1-59·3 | 10 | 22·2 | 9·9-34·6 | 15 | 46·9 | 29·2-64·5 | 24 | 38·7 | 26·4-51 | 3 | 11·1 | 0-23·3 | 0·009** |
|  | Far | 138 | 66·0 | 59·7-72·3 | 24 | 55·8 | 40·7-70·9 | 35 | 77·8 | 65·4-90·1 | 17 | 53·1 | 35·5-70·8 | 38 | 61·3 | 49-73·6 | 24 | 88·9 | 76·7-100 |  |
|  | *Missing* | *0* | *0·0* |  | *0* | *0·0* |  | *0* | *0·0* |  | *0* | *0·0* |  | *0* | *0·0* |  | *0* | *0·0* |  |  |
| **Proximity of a vaccination center (>15 or <15 min by foot)** | Close | 95 | 45·5 | 38·9-52 | 15 | 34·9 | 20·4-49·4 | 30 | 66·7 | 52·7-80·7 | 18 | 56·2 | 38·7-73·8 | 17 | 27·4 | 16·2-38·7 | 15 | 55·6 | 36·3-74·8 | p<0·001*** |
|  | Far | 114 | 54·5 | 48-61·1 | 28 | 65·1 | 50·6-79·6 | 15 | 33·3 | 19·3-47·3 | 14 | 43·8 | 26·2-61·3 | 45 | 72·6 | 61·3-83·8 | 12 | 44·4 | 25·2-63·7 |  |
|  | *Missing* | *0* | *0·0* |  | *0* | *0·0* |  | *0* | *0·0* |  | *0* | *0·0* |  | *0* | *0·0* |  | *0* | *0·0* |  |  |
| **Proximity of a GP (>15 or <15 min by foot)** | Close | 183 | 87·6 | 83-92·1 | 36 | 83·7 | 72·5-95 | 40 | 88·9 | 79·5-98·2 | 30 | 93·8 | 85·2-100 | 52 | 83·9 | 74·6-93·2 | 25 | 92·6 | 82·5-100 | 0·542 |
|  | Far | 26 | 12·4 | 7·9-17 | 7 | 16·3 | 5-27·5 | 5 | 11·1 | 1·8-20·5 | 2 | 6·2 | 0-14·8 | 10 | 16·1 | 6·8-25·4 | 2 | 7·4 | 0-17·5 |  |
|  | *Missing* | *0* | *0·0* |  | *0* | *0·0* |  | *0* | *0·0* |  | *0* | *0·0* |  | *0* | *0·0* |  | *0* | *0·0* |  |  |
| **Proximity of a drugstore (>15 or <15 min by foot)** | Close | 187 | 89·5 | 85·3-93·6 | 36 | 83·7 | 72·5-95 | 41 | 91·1 | 82·7-99·6 | 31 | 96·9 | 90·7-100 | 53 | 85·5 | 76·6-94·4 | 26 | 96·3 | 89-100 | 0·222 |
|  | Far | 22 | 10·5 | 6·4-14·7 | 7 | 16·3 | 5-27·5 | 4 | 8·9 | 0·4-17·3 | 1 | 3·1 | 0-9·3 | 9 | 14·5 | 5·6-23·4 | 1 | 3·7 | 0-11 |  |
|  | *Missing* | *0* | *0·0* |  | *0* | *0·0* |  | *0* | *0·0* |  | *0* | *0·0* |  | *0* | *0·0* |  | *0* | *0·0* |  |  |
| **Proximity of a mobile clinic (targeting PEH)** | Close | 74 | 35·4 | 29·2-41·6 | 8 | 18·6 | 6·8-30·4 | 9 | 20·0 | 8·1-31·9 | 15 | 46·9 | 29·2-64·5 | 34 | 54·8 | 42·3-67·4 | 8 | 29·6 | 12-47·3 | p<0·001*** |
|  | Far | 135 | 64·6 | 58·4-70·8 | 35 | 81·4 | 69·6-93·2 | 36 | 80·0 | 68·1-91·9 | 17 | 53·1 | 35·5-70·8 | 28 | 45·2 | 32·6-57·7 | 19 | 70·4 | 52·7-88 |  |
|  | *Missing* | *0* | *0·0* |  | *0* | *0·0* |  | *0* | *0·0* |  | *0* | *0·0* |  | *0* | *0·0* |  | *0* | *0·0* |  |  |
| **Intervention on site - Awareness/sensitization** | No | 68 | 32·5 | 28-37·1 |  |  |  | 6 | 13·3 | 3·2-23·4 |  |  |  | 48 | 77·4 | 66·9-88 | 14 | 51·9 | 32·5-71·2 | p<0·001*** |
|  | Yes | 141 | 67·5 | 62·9-72 | 43 | 100·0 | 100-100 | 39 | 86·7 | 76·6-96·8 | 32 | 100·0 | 100-100 | 14 | 22·6 | 12-33·1 | 13 | 48·1 | 28·8-67·5 |  |
|  | *Missing* | *0* | *0·0* |  | *0* | *0·0* |  | *0* | *0·0* |  | *0* | *0·0* |  | *0* | *0·0* |  | *0* | *0·0* |  |  |
| **Intervention on site - Personalized support/mobilization** | No | 136 | 65·1 | 59-71·1 | 16 | 37·2 | 22·5-51·9 | 34 | 75·6 | 62·8-88·3 | 16 | 50·0 | 32·3-67·7 | 53 | 85·5 | 76·6-94·4 | 17 | 63·0 | 44·3-81·6 | p<0·001*** |
|  | Yes | 73 | 34·9 | 28·9-41 | 27 | 62·8 | 48·1-77·5 | 11 | 24·4 | 11·7-37·2 | 16 | 50·0 | 32·3-67·7 | 9 | 14·5 | 5·6-23·4 | 10 | 37·0 | 18·4-55·7 |  |
|  | *Missing* | *0* | *0·0* |  | *0* | *0·0* |  | *0* | *0·0* |  | *0* | *0·0* |  | *0* | *0·0* |  | *0* | *0·0* |  |  |
| **Intervention on site - Actual vaccination activity** | No | 142 | 67·9 | 62·4-73·5 | 16 | 37·2 | 22·5-51·9 | 38 | 84·4 | 73·7-95·2 | 12 | 37·5 | 20·4-54·6 | 56 | 90·3 | 82·9-97·8 | 20 | 74·1 | 57·1-91 | p<0·001*** |
|  | Yes | 67 | 32·1 | 26·5-37·6 | 27 | 62·8 | 48·1-77·5 | 7 | 15·6 | 4·8-26·3 | 20 | 62·5 | 45·4-79·6 | 6 | 9·7 | 2·2-17·1 | 7 | 25·9 | 9-42·9 |  |
|  | *Missing* | *0* | *0·0* |  | *0* | *0·0* |  | *0* | *0·0* |  | *0* | *0·0* |  | *0* | *0·0* |  | *0* | *0·0* |  |  |

#### Appendix 11. Table S4. Multivariate Negative Binomial Regression (Site-Level)

| VARIABLES | Categories | Adjusted Risk Ratio | 95%CI | p-value |
| --- | --- | --- | --- | --- |
| Total vaccinated | Nb |  | - | · |
| Stratum | Housed | ref |  |  |
|  | Accommodated | 0·911 | 0·876 - 0·948 | <0·001*** |
|  | Streets | 0·644 | 0·579 - 0·716 | <0·001*** |
| Presence of social workers on site | Never or <once a month | ref |  |  |
|  | At least once per week | 1·301 | 1·007 - 1·680 | 0·044** |
|  | Unknown/Not applicable | 1·298 | 1·008 - 1·671 | 0·043** |
| Presence of health professionals on site | Never or <once a month | ref |  |  |
|  | At least once per week | 0·879 | 0·692 - 1·116 | 0·289 |
|  | Unknown/Not applicable | 0·836 | 0·659 - 1·060 | 0·139 |
| Awareness Raising/sensitization | Yes vs No (Ref) | 1·101 | 1·007 - 1·204 | 0·035** |
| Personalized support/mobilization | Yes vs No (Ref) | 1·042 | 1·002 - 1·082 | 0·038** |
| Actual vaccination activity on site | Yes vs No (Ref) | 1·114 | 1·068 - 1·163 | <0·001*** |
| Proximity of site targeting PEH | Close (<=15 min) | ref |  |  |
|  | Far (>15min) | 0·960 | 0·924 - 0·998 | 0·040** |
| Proximity of closed drugstore | Close (<=15 min) | ref |  |  |
|  | Far (>15min) | 0·988 | 0·925 - 1·055 | 0·726 |
| Proximity of closest vaccination center | Close (<=15 min) | ref |  |  |
|  | Far (>15min) | 1·012 | 0·974 - 1·051 | 0·538 |
| Constant | Constant | 0·643 | 0·553 - 0·748 | <0·001*** |

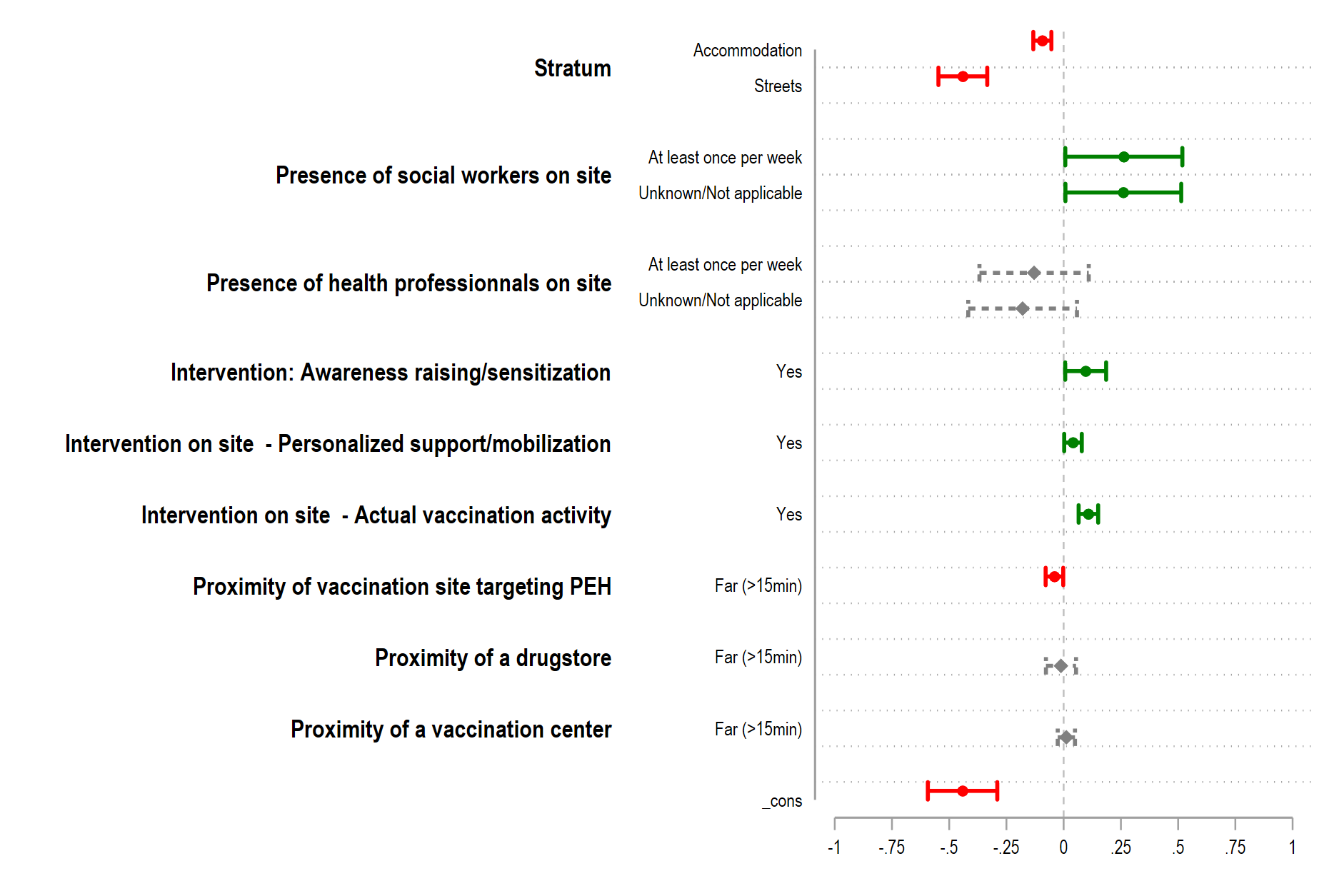

#### Appendix 12. Table S5. Stratified Analysis –

|  | Housing | | | Accomodation | | | Homeless (streets) | | |
| --- | --- | --- | --- | --- | --- | --- | --- | --- | --- |
| *Predictors* | *Odds Ratios* | *CI* | *p* | *Odds Ratios* | *CI* | *p* | *Odds Ratios* | *CI* | *p* |
| (Intercept) | 54·9 | 18·3 – 164·3 | **<0·001** | 7·1 | 3·6 – 14·1 | **<0·001** | 14·1 | 4·4 – 45·7 | **<0·001** |
| Gender: Male | *Reference* |  |  | *Reference* |  |  | *Reference* |  |  |
| Gender: Female | 2·1 | 0·6 – 7·1 | 0·232 | 0·8 | 0·7 – 1·1 | 0·194 | 0·8 | 0·4 – 1·6 | 0·539 |
| Age categories: Less than 35 years | *Reference* |  |  | *Reference* |  |  | *Reference* |  |  |
| Age categories: From 35 to 65 years | 1·2 | 0·7 – 2·1 | 0·487 | 1·5 | 1·2 – 1·9 | **<0·001** | 0·6 | 0·3 – 1·0 | 0·052 |
| Age categories: More than 65 years | 2·5 | 0·9 – 7·0 | 0·072 | 1·8 | 0·9 – 3·7 | 0·102 | 2·1 | 0·4 – 12·5 | 0·409 |
| Francophone: Yes | *Reference* |  |  | *Reference* |  |  | *Reference* |  |  |
| Francophone: No | 1·4 | 0·7 – 2·6 | 0·297 | 0·8 | 0·6 – 1·1 | 0·121 | 0·4 | 0·2 – 0·9 | **0·035** |
| Level of education: Never attended school / Illiterate | *Reference* |  |  | *Reference* |  |  | *Reference* |  |  |
| Level of education: Primary and/or literate | 1·2 | 0·6 – 2·7 | 0·579 | 0·8 | 0·5 – 1·1 | 0·192 | 0·9 | 0·4 – 1·9 | 0·713 |
| Level of education: Secondary | 0·6 | 0·3 – 1·2 | 0·149 | 0·8 | 0·5 – 1·2 | 0·354 | 0·7 | 0·3 – 1·5 | 0·340 |
| Level of education: Higher (university) | 0·7 | 0·3 – 1·5 | 0·327 | 1·0 | 0·6 – 1·7 | 0·966 | 1·7 | 0·6 – 4·8 | 0·316 |
| Attends food distributions on a regular basis : Autonomous / Does not receive or need support whatsoever: No | *Reference* |  |  | *Reference* |  |  | *Reference* |  |  |
| Attends food distributions on a regular basis : Autonomous / Does not receive or need support whatsoever: Yes | 1·5 | 0·9 – 2·5 | 0·125 | 1·6 | 1·2 – 2·3 | **0·005** | 0·5 | 0·2 – 0·9 | **0·026** |
| Perceived utility of Covid-19 vaccination : Yes | *Reference* |  |  | *Reference* |  |  | *Reference* |  |  |
| Perceived utility of Covid-19 vaccination : No | 0·1 | 0·1 – 0·2 | **<0·001** | 0·2 | 0·2 – 0·3 | **<0·001** | 0·1 | 0·1 – 0·2 | **<0·001** |
| Perceived utility of Covid-19 vaccination : No opinion | 0·3 | 0·2 – 0·7 | **0·004** | 0·3 | 0·2 – 0·5 | **<0·001** | 0·4 | 0·2 – 0·9 | **0·021** |
| Fear of Covid-19 vaccination : No | *Reference* |  |  | *Reference* |  |  | *Reference* |  |  |
| Fear of Covid-19 vaccination : Yes | 0·3 | 0·2 – 0·7 | **0·004** | 0·6 | 0·5 – 0·8 | **<0·001** |  |  |  |
| Needs/uses vaccine certificate in daily life: Yes | *Reference* |  |  | *Reference* |  |  | *Reference* |  |  |
| Needs/uses vaccine certificate in daily life: No | 0·3 | 0·2 – 0·6 | **<0·001** | 0·3 | 0·2 – 0·4 | **<0·001** | 0·3 | 0·2 – 0·5 | **<0·001** |
| Is followed by a regular GP: Yes | *Reference* |  |  | *Reference* |  |  | *Reference* |  |  |
| Is followed by a regular GP: No | 0·5 | 0·3 – 0·9 | **0·012** | 0·7 | 0·6 – 0·9 | **0·014** |  |  |  |
| Date of last consultation (hospital/GP): After the start of vaccination campaign (>may 2021) | *Reference* |  |  | *Reference* |  |  | *Reference* |  |  |
| Date of last consultation (hospital/GP): Between 2d lockdown and start of vaccination (dec 2020-may 2021) | 0·4 | 0·2 – 0·8 | **0·012** |  |  |  | 0·6 | 0·3 – 1·5 | 0·314 |
| Date of last consultation (hospital/GP): Between 1st and 2d lockdown (mar-dec 2020) | 0·6 | 0·3 – 1·2 | 0·120 |  |  |  | 0·6 | 0·3 – 1·3 | 0·167 |
| Date of last consultation (hospital/GP): Before the pandemic (<march 2020) | 0·4 | 0·2 – 1·0 | 0·052 |  |  |  | 0·3 | 0·1 – 0·6 | **0·002** |
| Date of last consultation (hospital/GP): Never consulted in France/Does not apply | 0·8 | 0·2 – 2·5 | 0·635 |  |  |  | 0·9 | 0·4 – 2·1 | 0·881 |
| Previous history/hospitalization for COVID-19 among family or friends: No | *Reference* |  |  | *Reference* |  |  | *Reference* |  |  |
| Previous history/hospitalization for COVID-19 among family or friends: Yes, without hospitalization | 0·4 | 0·2 – 0·8 | **0·011** |  |  |  |  |  |  |
| Previous history/hospitalization for COVID-19 among family or friends: Yes, with hospitalization | 0·7 | 0·4 – 1·2 | 0·187 |  |  |  |  |  |  |
| COVID-19 (Vaccine) information source - internet / social media: No | *Reference* |  |  | *Reference* |  |  | *Reference* |  |  |
| COVID-19 (Vaccine) information source - internet / social media: Yes | 0·4 | 0·3 – 0·7 | **0·002** | 0·6 | 0·5 – 0·8 | **<0·001** |  |  |  |
| COVID-19 (Vaccine) information source - television/radio: No | *Reference* |  |  | *Reference* |  |  | *Reference* |  |  |
| COVID-19 (Vaccine) information source - television/radio: Yes | 1·7 | 1·0 – 2·9 | 0·051 | 0·8 | 0·6 – 1·0 | 0·109 |  |  |  |
| Proximity of a drugstore (>15 or <15 min by foot): Close | *Reference* |  |  | *Reference* |  |  | *Reference* |  |  |
| Proximity of a drugstore (>15 or <15 min by foot): Far | 3·3 | 0·7 – 14·9 | 0·128 | 0·7 | 0·5 – 1·0 | **0·032** |  |  |  |
| Proximity of a mobile clinic (targeting PEH): Close | *Reference* |  |  | *Reference* |  |  | *Reference* |  |  |
| Proximity of a mobile clinic (targeting PEH): Far | 0·7 | 0·4 – 1·1 | 0·138 |  |  |  |  |  |  |
| Administrative status: French or European nationality (EU zone) | *Reference* |  |  | *Reference* |  |  | *Reference* |  |  |
| Administrative status: Asylum seeker / Ongoing application |  |  |  | 2·2 | 1·5 – 3·2 | **<0·001** |  |  |  |
| Administrative status: Residence permit / Refugee status |  |  |  | 1·4 | 0·9 – 2·2 | 0·092 |  |  |  |
| Administrative status: Undocumented |  |  |  | 1·5 | 1·1 – 2·2 | **0·021** |  |  |  |
| Meals provided by site managers: No | *Reference* |  |  | *Reference* |  |  | *Reference* |  |  |
| Meals provided by site managers: Yes |  |  |  | 1·7 | 1·2 – 2·4 | **0·002** |  |  |  |
| Attends food distributions on a regular basis : No | *Reference* |  |  | *Reference* |  |  | *Reference* |  |  |
| Attends food distributions on a regular basis : Yes |  |  |  | 1·5 | 1·1 – 2·0 | **0·004** |  |  |  |
| Opinion on vaccination (in general): Positive | *Reference* |  |  | *Reference* |  |  | *Reference* |  |  |
| Opinion on vaccination (in general): Negative |  |  |  | 0·5 | 0·4 – 0·7 | **<0·001** | 0·5 | 0·3 – 1·0 | 0·063 |
| Has medical coverage: Yes (national social security, state medical aid) | *Reference* |  |  | *Reference* |  |  | *Reference* |  |  |
| Has medical coverage: No (never, lost or ongoing application) |  |  |  | 0·6 | 0·4 – 0·8 | **<0·001** | 0·4 | 0·3 – 0·8 | **0·005** |
| Previous history/hospitalization of COVID-19: No | *Reference* |  |  | *Reference* |  |  | *Reference* |  |  |
| Previous history/hospitalization of COVID-19: Yes, without hospitalization |  |  |  | 0·8 | 0·6 – 1·0 | 0·066 |  |  |  |
| Previous history/hospitalization of COVID-19: Yes, with hospitalization |  |  |  | 2·4 | 1·0 – 5·9 | 0·062 |  |  |  |
| Can read and understand written medical instructions: With ease overall, alone | *Reference* |  |  | *Reference* |  |  | *Reference* |  |  |
| Can read and understand written medical instructions: With difficulty overall, alone |  |  |  | 0·6 | 0·4 – 1·0 | **0·030** |  |  |  |
| Can correctly fill out a medical form: With ease overall, alone | *Reference* |  |  | *Reference* |  |  | *Reference* |  |  |
| Can correctly fill out a medical form: With difficulty overall, alone |  |  |  | 1·7 | 1·1 – 2·4 | **0·008** |  |  |  |
| COVID-19 (Vaccine) information source - site manager: No | *Reference* |  |  | *Reference* |  |  | *Reference* |  |  |
| COVID-19 (Vaccine) information source - site manager: Yes |  |  |  | 2·1 | 1·1 – 3·9 | **0·019** |  |  |  |
| Intervention on site - Actual vaccination activity: No | *Reference* |  |  | *Reference* |  |  | *Reference* |  |  |
| Intervention on site - Actual vaccination activity: Yes |  |  |  | 1·5 | 1·1 – 2·0 | **0·004** |  |  |  |
| Household structure : Live by him/herself | *Reference* |  |  | *Reference* |  |  | *Reference* |  |  |
| Household structure : Live with his/her partner and children |  |  |  |  |  |  | 0·3 | 0·1 – 0·7 | **0·005** |
| Household structure : Live with his/her children, without other adults |  |  |  |  |  |  | 0·5 | 0·1 – 2·1 | 0·346 |
| Household structure : Live with his/her partner or related adults, without children |  |  |  |  |  |  | 1·0 | 0·5 – 2·1 | 0·907 |
| Household structure : Live with other non-related adults |  |  |  |  |  |  | 1·4 | 0·6 – 3·5 | 0·427 |
| Meals provided by relatives/family/friends: No | *Reference* |  |  | *Reference* |  |  | *Reference* |  |  |
| Meals provided by relatives/family/friends: Yes |  |  |  |  |  |  | 0·4 | 0·2 – 0·8 | **0·006** |
| Meals through food distributions: No | *Reference* |  |  | *Reference* |  |  | *Reference* |  |  |
| Meals through food distributions: Yes |  |  |  |  |  |  | 1·8 | 1·0 – 3·3 | **0·036** |
| Supported by site managers/social workers on site: No | *Reference* |  |  | *Reference* |  |  | *Reference* |  |  |
| Supported by site managers/social workers on site: Yes |  |  |  |  |  |  | 0·3 | 0·1 – 0·8 | **0·012** |
| Supported by health professionals: Attends food distributions on a regular basis : No | *Reference* |  |  | *Reference* |  |  | *Reference* |  |  |
| Supported by health professionals: Attends food distributions on a regular basis : Yes |  |  |  |  |  |  | 0·4 | 0·1 – 1·2 | 0·109 |
| Can understand oral medical instructions : With ease overall, alone | *Reference* |  |  | *Reference* |  |  | *Reference* |  |  |
| Can understand oral medical instructions : With difficulty overall, alone |  |  |  |  |  |  | 1·9 | 0·9 – 4·0 | 0·097 |
| COVID-19 (Vaccine) information source - health or social workers/professionnals: No | *Reference* |  |  | *Reference* |  |  | *Reference* |  |  |
| COVID-19 (Vaccine) information source - health or social workers/professionnals: Yes |  |  |  |  |  |  | 1·9 | 1·0 – 3·6 | **0·046** |
| Random Effects | | | | | | | | | |
| σ^2^ | 3·29 | | | 3·29 | | | 3·29 | | |
| τ_00_ | 1·45 _site_ | | | 0·08 _site_ | | | 0·23 _site_ | | |
| τ_11_ | 2·78 _site·vaccin_peur·newYes_ | | |  | | |  | | |
| ρ_01_ | -0·97 _site_ | | |  | | |  | | |
| ICC | 0·24 | | | 0·02 | | | 0·06 | | |
| N | 61 _site_ | | | 148 _site_ | | | 85 _site_ | | |
| Observations | 818 | | | 2219 | | | 479 | | |
| Marginal R^2^ / Conditional R^2^ | 0·397 / 0·544 | | | 0·352 / 0·368 | | | 0·535 / 0·565 | | |
| AIC | 547·055 | | | 1992·924 | | | 482·440 | | |
| AICc | 548·969 | | | 1993·831 | | | 486·316 | | |

#### Appendix 13. Figure S2. Flow between original strata and strata for analysis

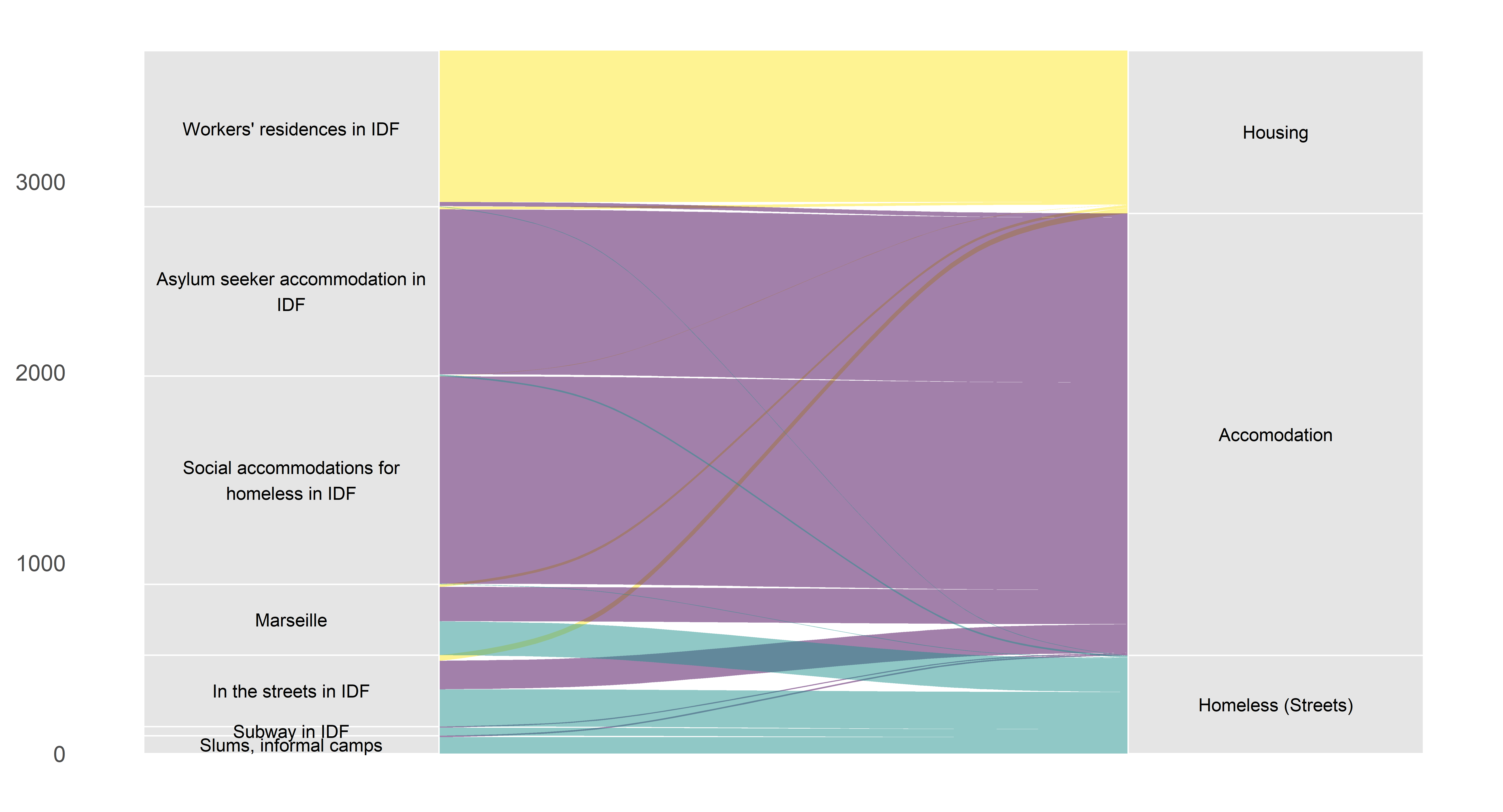
